# Comparative effectiveness of different exercises on bone mineral density in postmenopausal women: a systematic review and network meta-analysis of randomized controlled trials

**DOI:** 10.1101/2021.07.12.21260370

**Authors:** Yi Li, Xiaoyi Wang, Ruishi Zhang, Siyi Zhu, Liqiong Wang, Jingming Huang, Xinling Gan, Suhang Xie, Tao Wu, Chengqi He

## Abstract

**Objective:** To compare and rank different exercise interventions on bone mineral density (BMD) in postmenopausal women.

**Data Sources and Study Selection:** MEDLINE, EMBASE, CINAHL, AMED, the Cochrane Central Register of Controlled Trials (CENTRAL), Scopus and Web of Science were searched from database inception to January 2021 of randomized controlled studies investigating the effect of exercise more than six months on BMD in postmenopausal women.

**Data Extraction and Synthesis:** Data at baseline and post intervention (or the change from baseline) were extracted. A Bayesian random-effects network meta-analysis was performed.

**Main Outcomes and Measures:** The primary outcome was the change of BMD (at lumbar spine, femoral neck, and total hip) from baseline values. Effect size measures were mean differences with 95% credible intervals (CrIs).

**Results:** We identified 3324 citations and included 66 studies with a total number of 4336 participants. Associated with BMD at lumbar spine (LS) improve were found for multicomponent exercise, resistance training, mind body exercise, lower impact exercise, high impact exercise, and whole body vibration. With regard to femoral neck (FN), only multicomponent exercise, whole body vibration, and mind body exercise were effective. As for total hip (TH), only multicomponent exercise, resistance training, and flexibility exercise were found to be beneficial. Moreover, no matter the age of postmenopausal women, and the duration of intervention (range between 6 to 18 months), some certain kinds of exercise could be performed to improve BMD at LS and FN.

**Conclusions and Relevance:** This NMA confirms that exercise therapy has clear benefits on bone mineral density in postmenopausal women. It also shows that the magnitude of effect varies depending on the outcome of interest, the age of participants, and the duration of intervention. Clinicians might consult the ranking of the exercise intervention presented in this study, when designating an optimal, individualized exercise prescription to improve BMD.

## Introduction

In the early years after menopause, postmenopausal women experience dramatic decreases in bone mineral density (BMD), which may increase the likelihood of developing osteoporosis and fragility fractures.^1, 2^ As reported, approximately one-third of postmenopausal women have osteoporosis. ^3^ Every year, the number of osteoporotic fracture is estimated to be about nine million worldwide, 61% of which are women.^4^ Osteoporotic fracture is associated with substantial morbidity and mortality, imposing a significant negative impact not only on the patients themselves, but also on society in terms of cost of health care and fracture therapy.^5^

Different types of physical exercise, including aerobic exercise, impact exercise, strength training and balance training are used in clinical practice to maintain or increase BMD.^6, 7^ Several systematic reviews, including meta-analyses, have investigated the efficacy of different exercise interventions for BMD.^8-10^ However, review articles reported that the BMD effects of exercise are modest at best,^11, 12^ which may partially due to the large variety of exercise regimens and participant characteristics. Therefore, it is challenging to decide which is the most appropriate treatment and this may account for the variation in clinical practice.

Traditional pairwise meta analyses depend on qualified randomized controlled trials (RCTs) having similar treatment and control groups, and studies with different comparators were excluded consequently. Network meta-analysis (NMA), which synthesises evidence from direct and indirect comparisons of multiple treatments, makes it practical for simultaneous inferences regarding clinical efficacy of all available treatment groups. ^13, 14^ Moreover, it has the potential to enhance the power of the estimation as compared to traditional meta analyses. In the meantime, NMA could inform clinicians and patients by ranking of the effectiveness of different treatments.^15^

To our knowledge, no study has yet used NMA to compare the effects of different exercises on bone mineral density in postmenopausal women. The objective of this systematic review and hierarchical Bayesian network meta-analysis was to pool data from RCTs to compare the relative effects of all available exercise interventions on BMD in postmenopausal women, providing an up-to-date and comprehensive overview of evidence-based treatments.

## Methods

### Data Sources and Search Strategies

This systematic review was performed according to the Preferred Reporting Items for Systematic Reviews and Meta-Analyses (PRISMA) statement and the PRISMA network meta-analysis extension statement. ^16, 17^ The review protocol was registered in the International Platform of Registered Systematic Review and Meta-analysis Protocols (identifier: INPLASY202140107).

Two independent reviewers performed the search procedures. The following electronic databases were searched from inception to 17 January, 2021: MEDLINE, EMBASE, CINAHL, AMED, the Cochrane Central Register of Controlled Trials (CENTRAL), Scopus and Web of Science. We also searched using the World Health Organization International Clinical Trials Registry Platform search portal, such as ClinicalTrials.gov and ISRCTN, to identify further studies. No language restriction was applied. Furthermore, the bibliographies of selected articles and relevant review articles were examined for additional potentially relevant studies. A combination of relevant free text terms, synonyms and subject headings relating to postmenopausal women, intervention of interest and randomized controlled trial were included in the strategy. The detailed search strategies for MEDLINE are provided in Supplementary Appendix 1, and the search terms will be adjusted appropriately to the different syntax requirements of the other databases.

### Inclusion and Exclusion Criteria

Articles were eligible if they met the following inclusion criteria: (1) RCTs designed to compare any therapeutic exercise intervention with other forms of exercise, sham exercise or no exercise control group with usual activity; (2) Subjects were healthy or osteoporotic postmenopausal women; (3) Intervention lasted at least 6 months of duration; (4) The study provided original data or sufficient information about at least one of the following outcomes: BMD measured by dual energy X-ray absorptiometry (DEXA) or dual-photon absorptiometry (DPA) at lumbar spine (LS), total hip (TH), or/and femoral neck (FN) locations. We further excluded studies with (1) cross-over trials, non-RCTs or non-clinical trials; (2) mixed gender or mixed pre- and postmenopausal cohorts without separate subgroup analyses; (3) the addition of any medications (e.g. bisphosphonates, glucocorticoid or hormone replacement therapy) with a dedicate effect on bone metabolism, but participants who consumed calcium and vitamin D were eligible for inclusion; (4) identical exercise interventions between the intervention groups; (5) conference abstracts with no corresponding full article published in a peer-reviewed journal or no specific data provided even after contacting the author.

### Categorization of exercises

In order to determine the effects of different types of exercise, we categorized the exercise intervention into ten categories. The definition of these interventions is provided in table 1.

**Table 1.** Definitions of exercise interventions and control comparator

| Type | Definition |
| --- | --- |
| Flexibility exercise (FE) | Exercises aimed at improving flexibility and coordination of body. |
| Whole body vibration (WBV) | A whole-body vibration platform was used to apply training. |
| Wellness exercise (WE) | Low intensity exercises that focused on well-being. |
| Lower impact exercise (LIE) | Exercises impose a small amount impact through the bones, such as standing, walking, stair climbing, and gentle heel drops. |
| Moderate impact exercise (MIE) | Exercise impose a moderate force by pushing off and returning to the ground. Both feet generally leave the ground but with less height and force than with high impact activity, such as running, jumping, hopping and heel drops with sufficient force |
| High impact exercise (HIE) | Exercise in which a large force is created on returning to the ground, usually from a greater height, like exertional jumps. |
| Mind body exercise (MBE) | Exercises emphasize the interactions among mind, body, psychology and behavior, intending to influence physical function and promote health, such as Tai Chi, yoga, Pilates and eight-section brocade. |
| Water based exercise (WBE) | Exercises performed in an aquatic environment in which the buoyancy of water enables limbs to move efficiently with less effort, improving the coordination of movement |
| Resistance training (RT) | Exercises using different modes of training with a wide range of resistive loads like free weights, resistance bands, or weights machines |
| Multicomponent exercise (ME) | A combination of more than one of the above exercise interventions within a single intervention group |
| Control | Participants continue their routine daily life activities, and try to avoid any systematic exercise training programs throughout the study. |

### Outcome Measures and Risk of bias Assessment

Pairs of members of the review team used a pre-piloted extraction form to independently extract relevant data. A third arbitrator was involved in case any disagreements occurred. From each study, we extracted general manuscript information (author, year of publication, region), demographic and clinical characteristics of participants (number of participants, age, body mass index (BMI), years since menopause, additional supplement), intervention protocols (type of exercise, frequency, duration, main part of exercise, setting of intervention, session length, compliance and adverse events), quality assessment and outcomes.

BMD at LS, TH or/and FN locations were assessed by DXA or DPA immediately post intervention. The primary outcome was change of BMD from baseline values (end-point score minus baseline score). If the results were merely presented in figures, values were extracted using a plot digitizer program. ^18^ Quality assessments were performed with the PEDro scale.^19^

### Data Synthesis and Statistical Analysis

The mean difference of the change score was used to estimate the effect size (ES) (Supplementary Appendix 2). Pooling of effect estimates across studies were conducted using random-effects models, due to the variation of the results of previous systematic reviews. ^8^

We performed a meta-analysis using STATA V16·0 (StataCorp, Texas, USA) for all available direct comparisons. Statistical heterogeneity was assessed by the Q test and estimated I2 value. Heterogeneity was considered moderate or high when I2 values were above 25% and 50%, respectively.^20^

We also developed network plots using Stata version 16 to visualize the relative amount of relative evidence and comparative relationships of different exercise interventions. Moreover, in order to check for publication bias which may be caused by small studies, a network funnel plot was generated when ten or more trials were available for one comparison.^21^

Network analysis was conducted in GeMTC 0·14·3 (Generate Mixed Treatment Comparisons). We checked for inconsistency between all direct and indirect evidence to compare different exercise interventions with regard to BMD at LS, FN, and TH, and calculated mean differences along with corresponding 95% credible interval (CrI) with a Bayesian Markov chain Monte Carlo method.

GeMTC was implemented as follows: the initial values scaling is 2·5, a long burn-in period (20000 iterations) and follow-up period (50000 iterations) with a thinning interval of 10 was allowed for convergence. The number of chains was set to four. Once the comparative efficacy of the interventions were identified, they were ranked to determine which one was superior. We used surface under cumulative ranking (SUCRA) rather than the probability of being the best (PrBest) intervention because SUCRA is regarded as the more precise method for calculating ranking probabilities.^13, 22^ The values of SUCRA equals one or zero means an exercise intervention ranks first or last, respectively. SUCRA was performed using OpenBUGS (Version 3·2·3).

We performed a subgroup analysis to establish the effectiveness relative to the intervention duration (≤8, 9–18, and >18 months, taking the remodeling cycle between cancellous and cortical bone into account^23^) and the age (<60 and≥60). The sensitivity analysis was conducted by deleting studies of low quality.

### Certainty of evidence

Confidence in Network Meta-Analysis (CINeMA) was used to assess the certainty of evidence from the NMA.24 Two independent researchers graded the evidence on the basis of within-study bias, reporting bias, indirectness, imprecision, heterogeneity and incoherence. We rated the evidence from the NMA as a whole to manifest the strength of the NMA recommendations.

## Results

A flowchart of the search process is shown in Supplementary Appendix 3. We identified 3324 records from the initial title and abstract screening, and a total of 66 studies with a total number of 4336 participants fulfilled the inclusion criteria and were finally included in this network meta-analysis. A detailed list of the included and excluded studies (with reasons) is provided in Supplementary Appendix 4.

The baseline characteristics of participants and exercise protocol of all the included studies are presented in Table 2 and 3. Sample size ranged from 15 to 246 participants (mean age range, 50·5–79·6 years). Most RCTs were from USA (n=15), followed by Australia (n=8) and Canada (n=7). The mean menopausal year ranged from at least 0·5 to 30 years, and the mean body mass index (BMI) varied from 19·7 to 28·8 kg/m2. The exercise intervention were delivered at home, center, or both. Compliance with the exercise ranged from 39% to 96%.

**Table 2.**
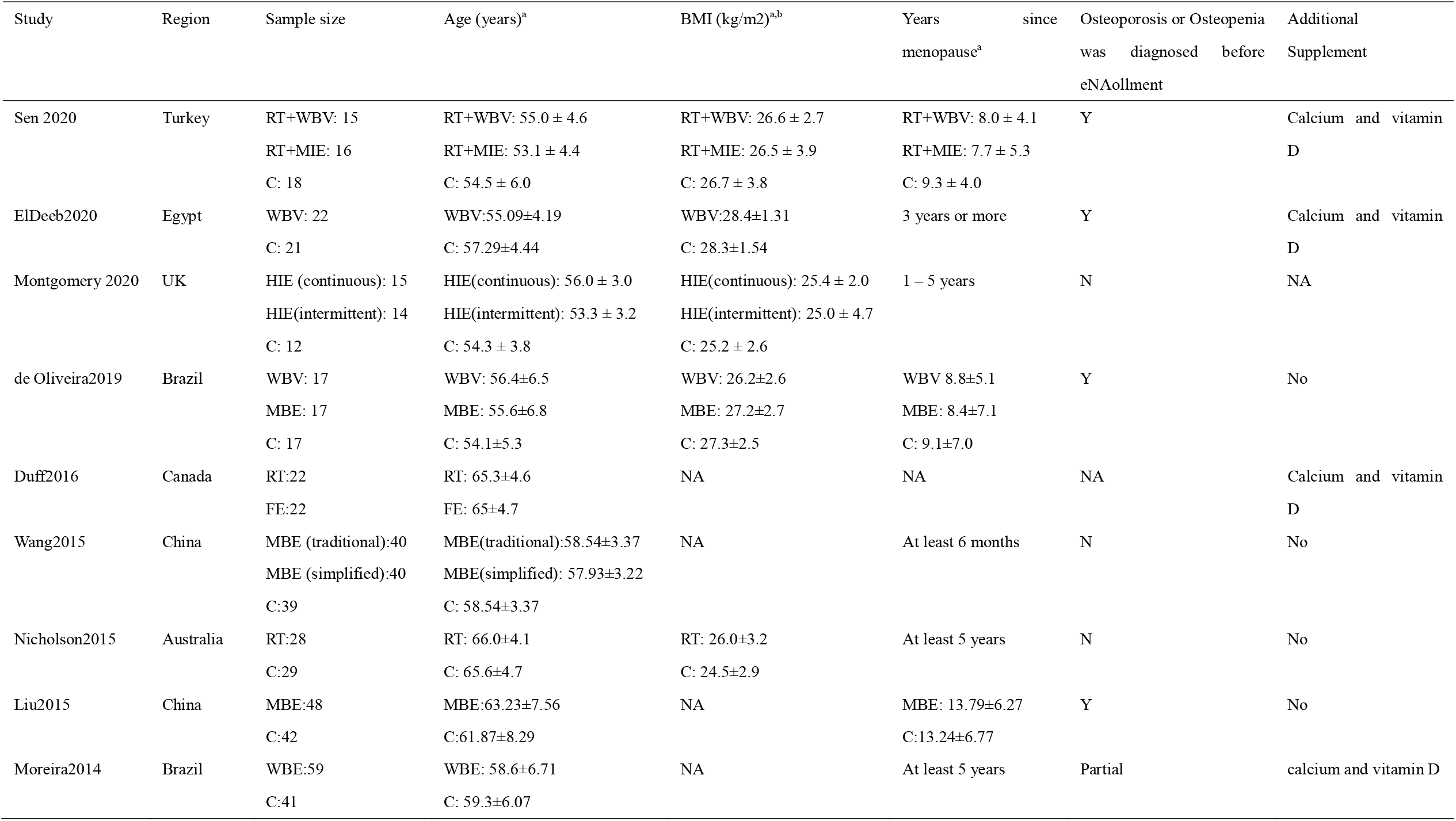
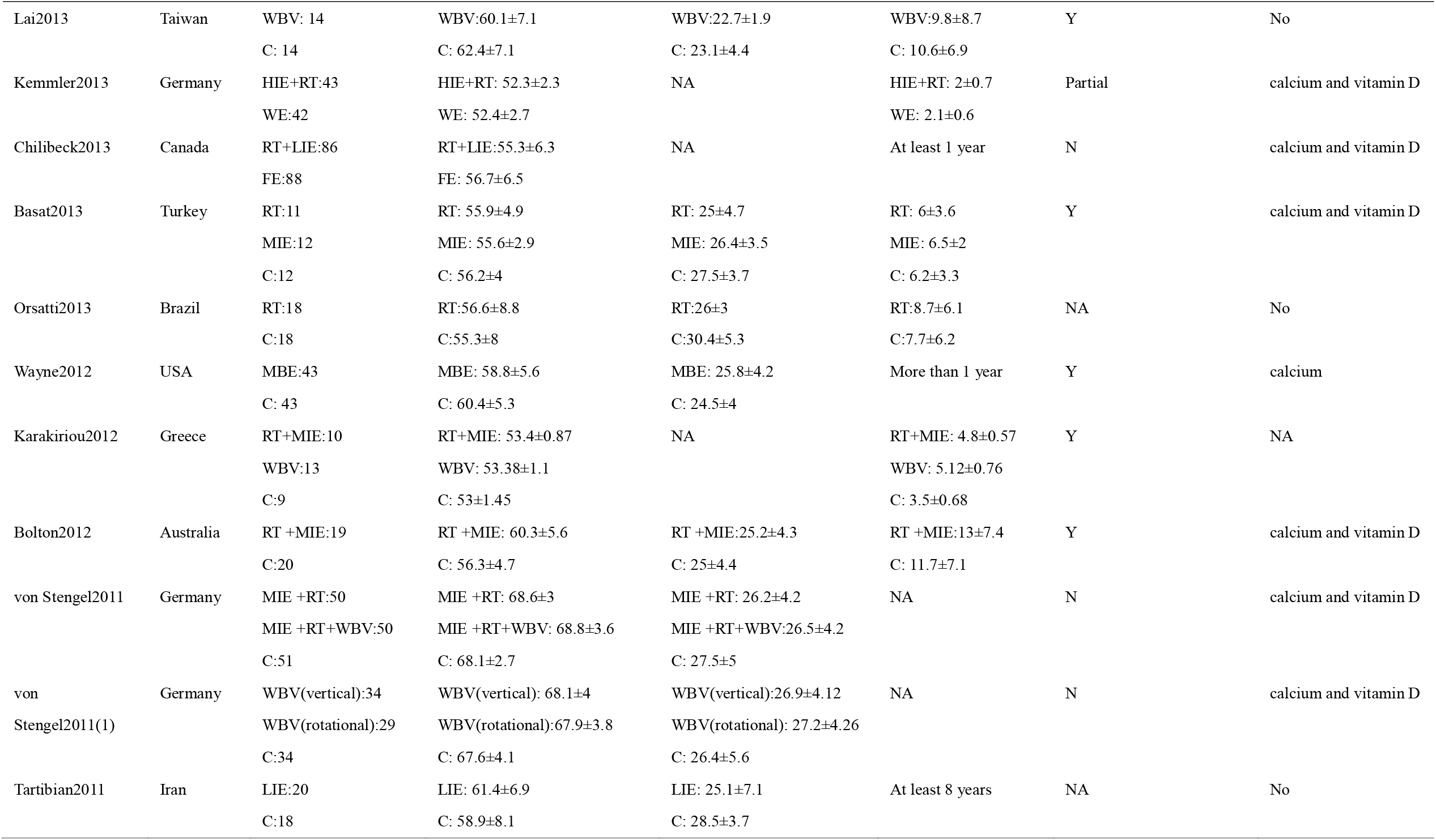
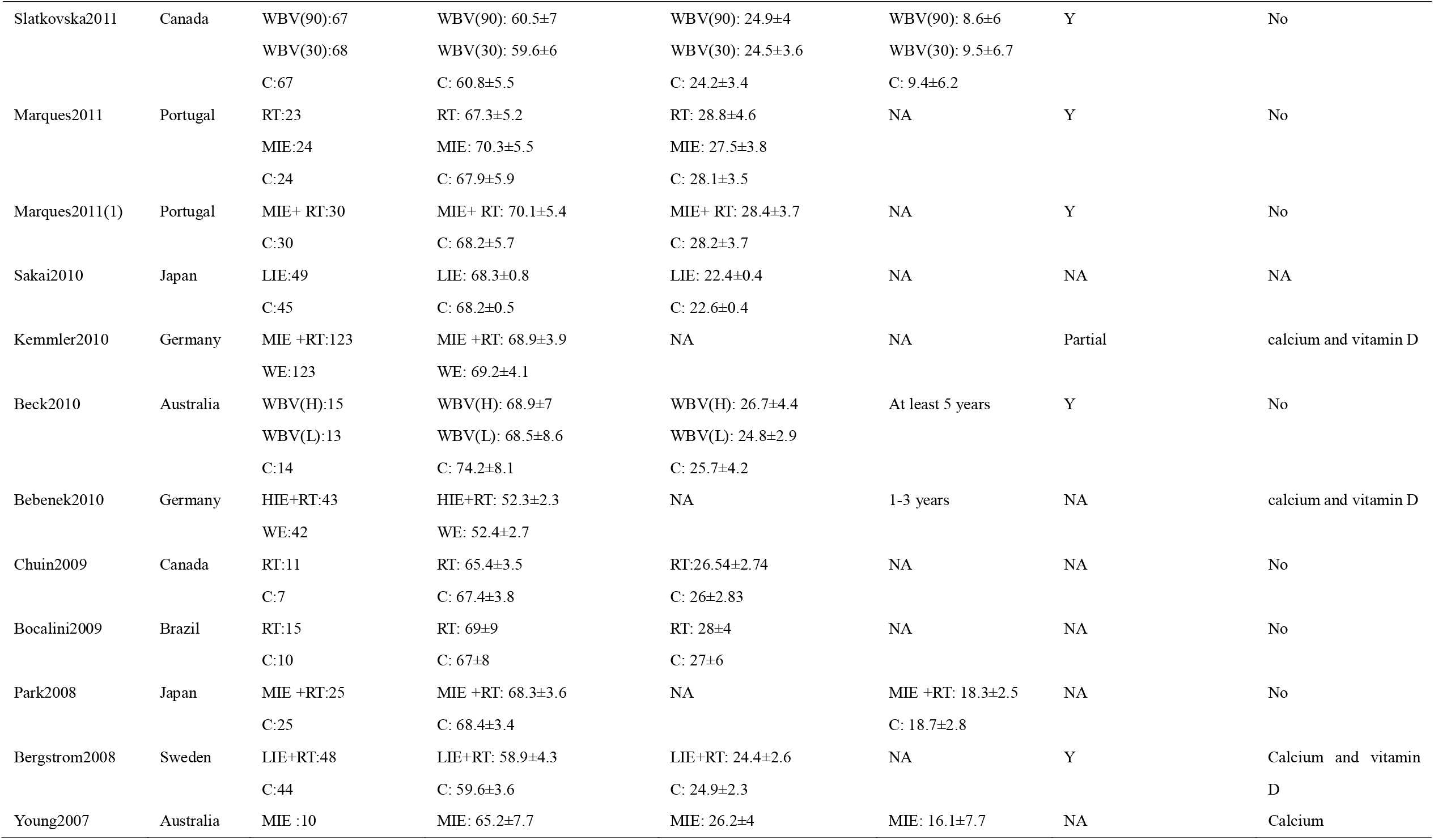
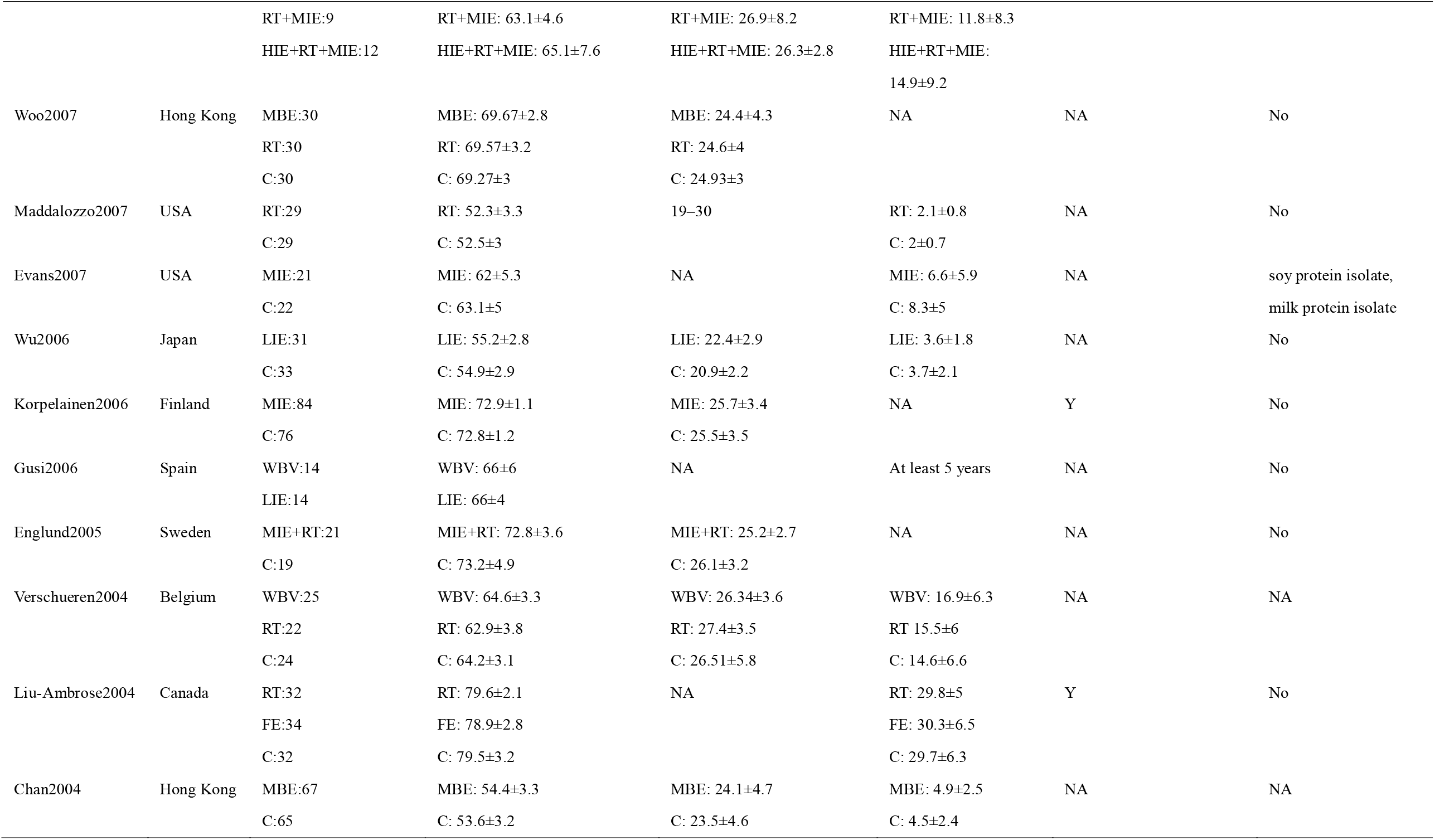
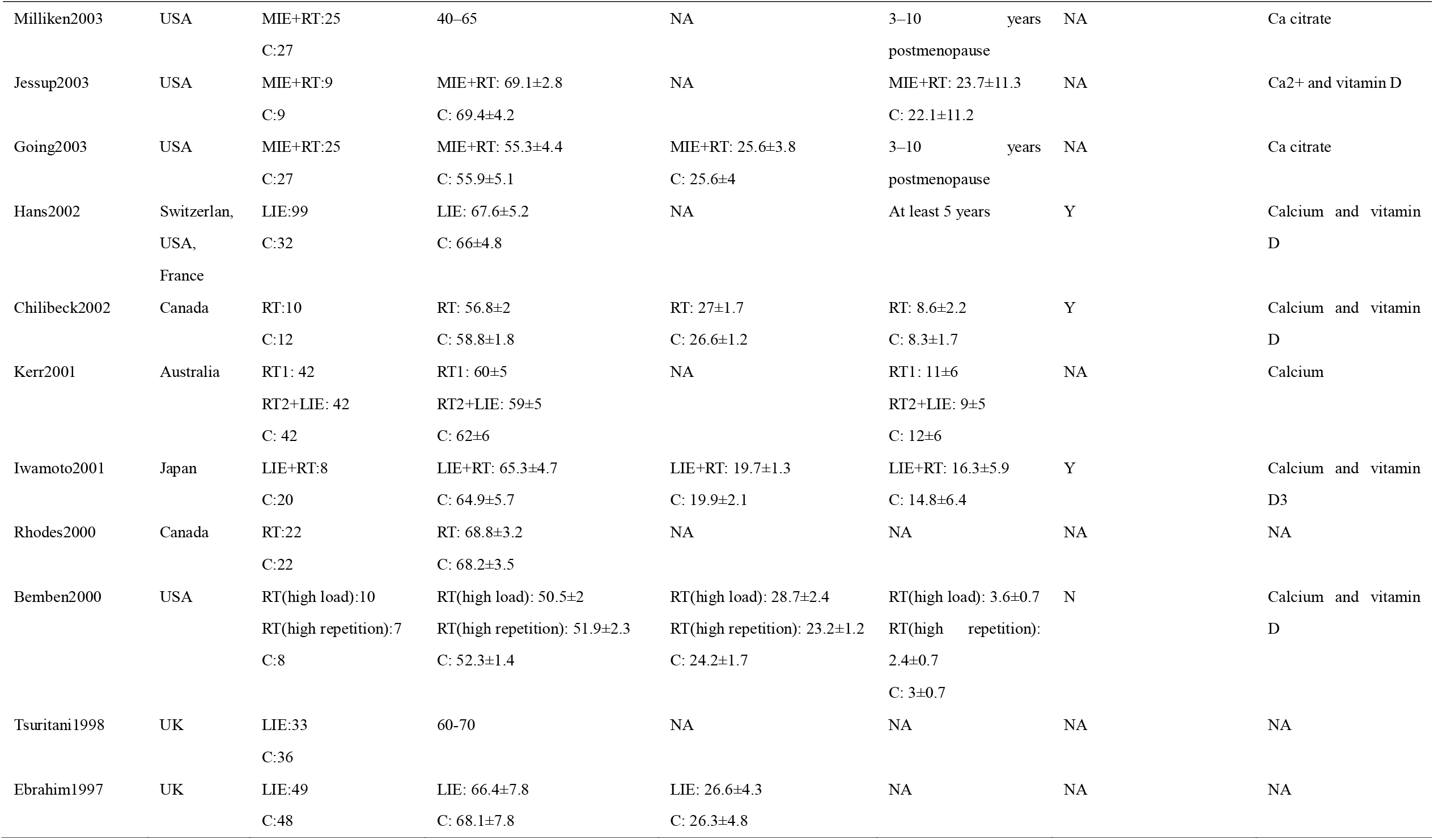
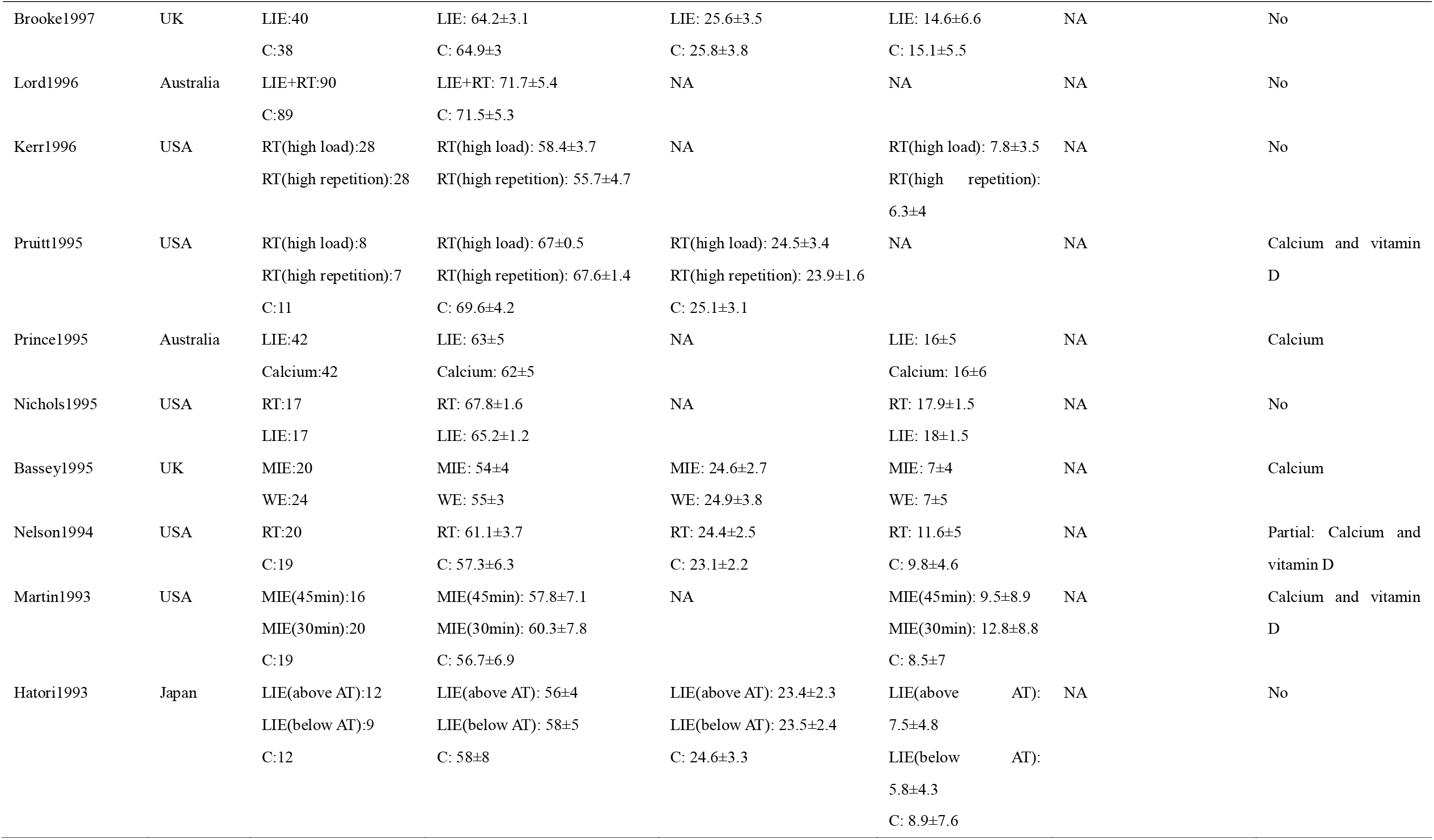
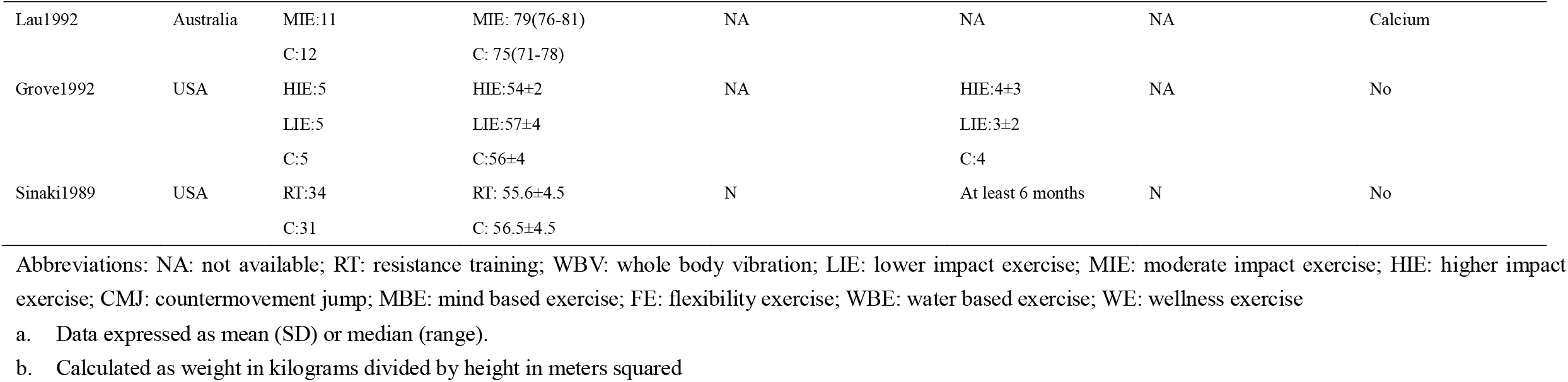
Characteristics of included studies

**Table 3.**
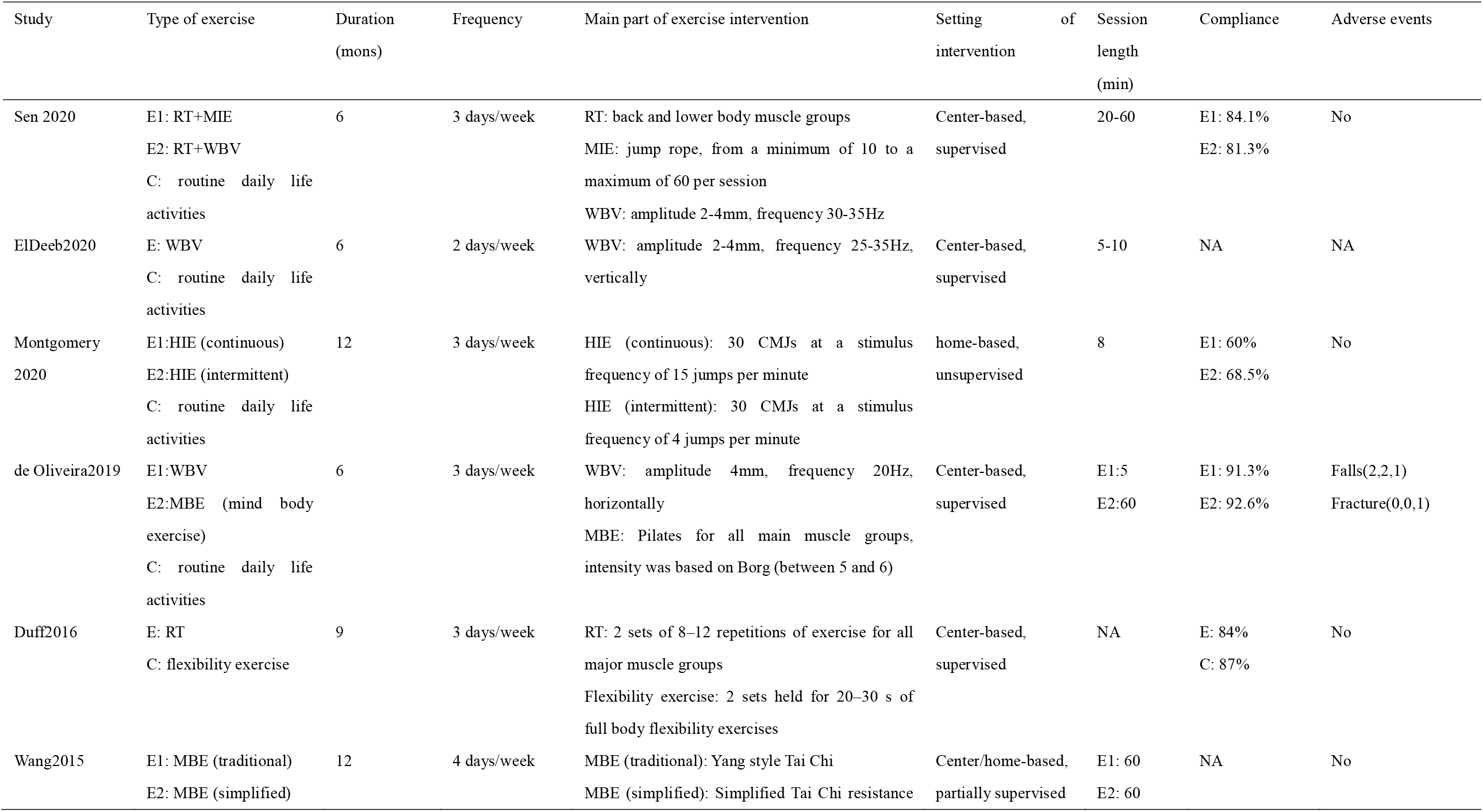
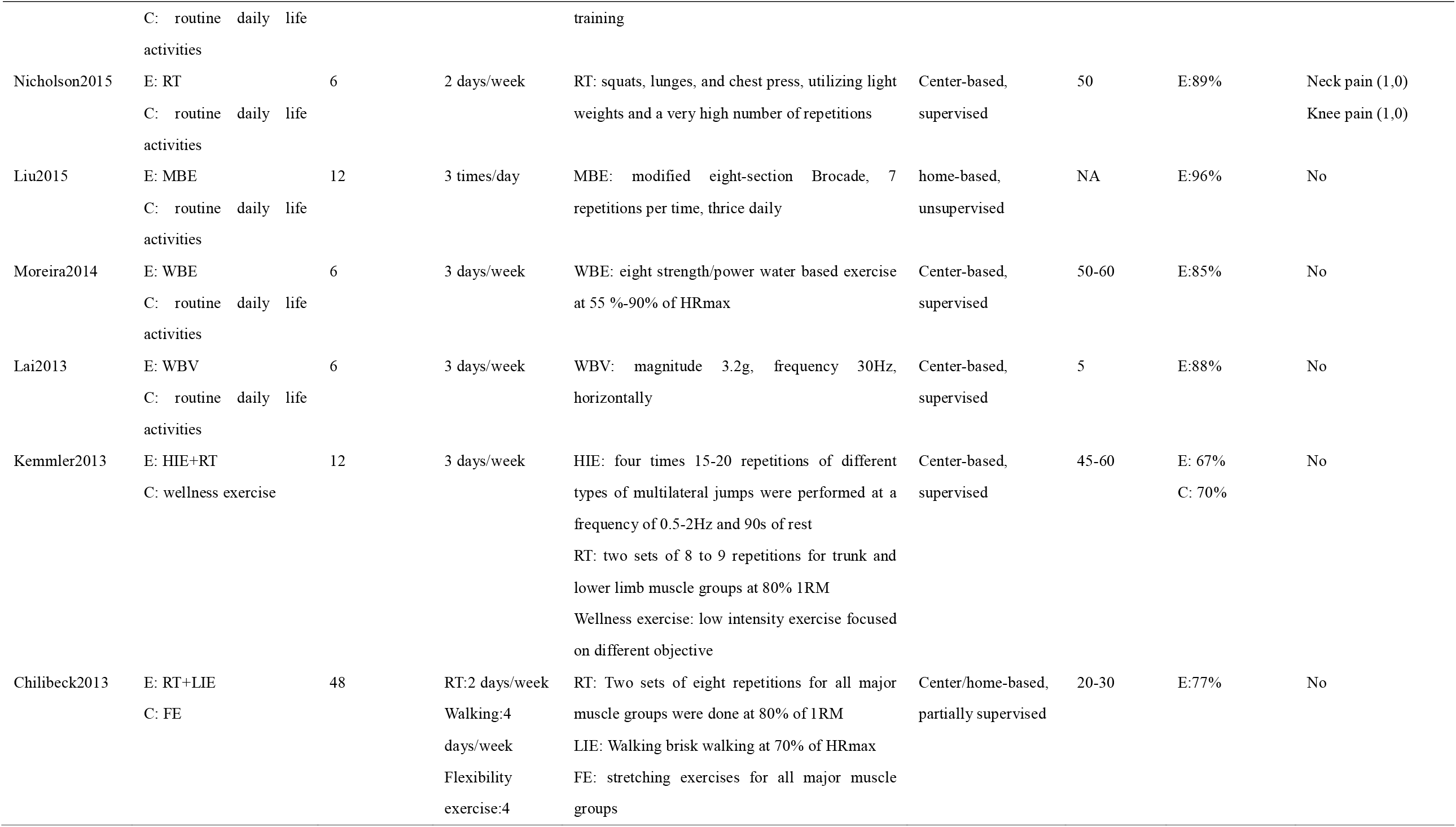
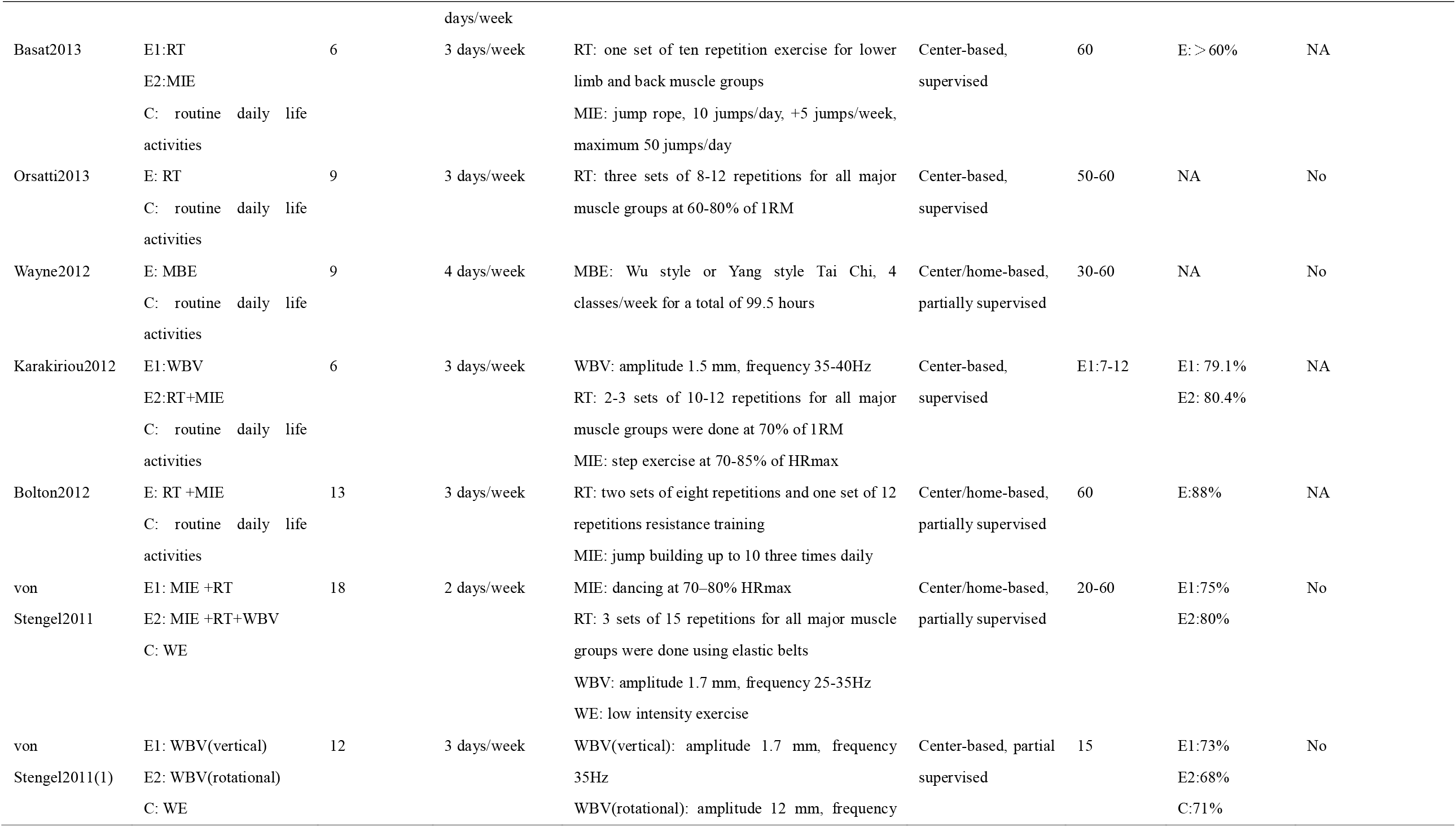
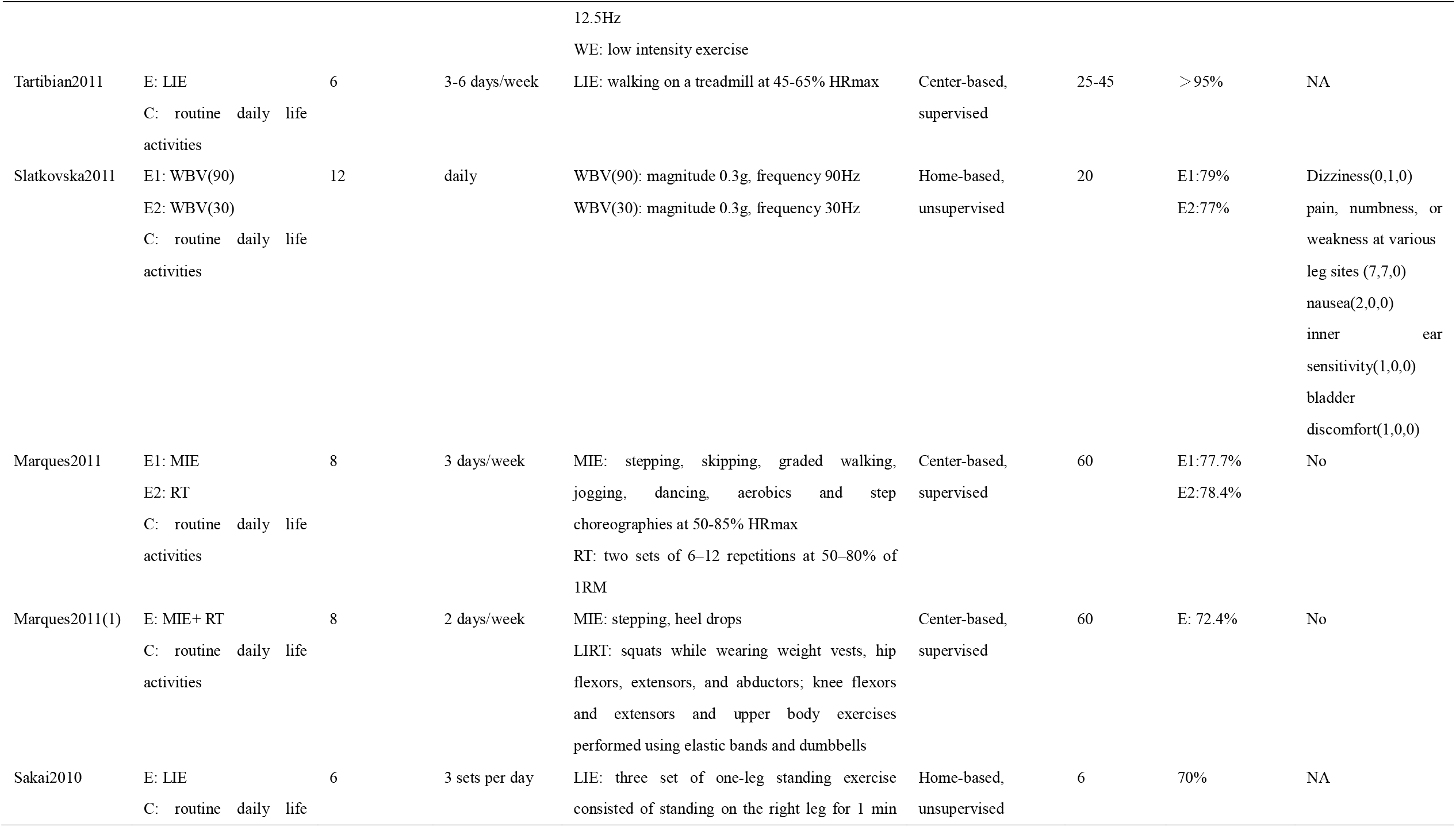
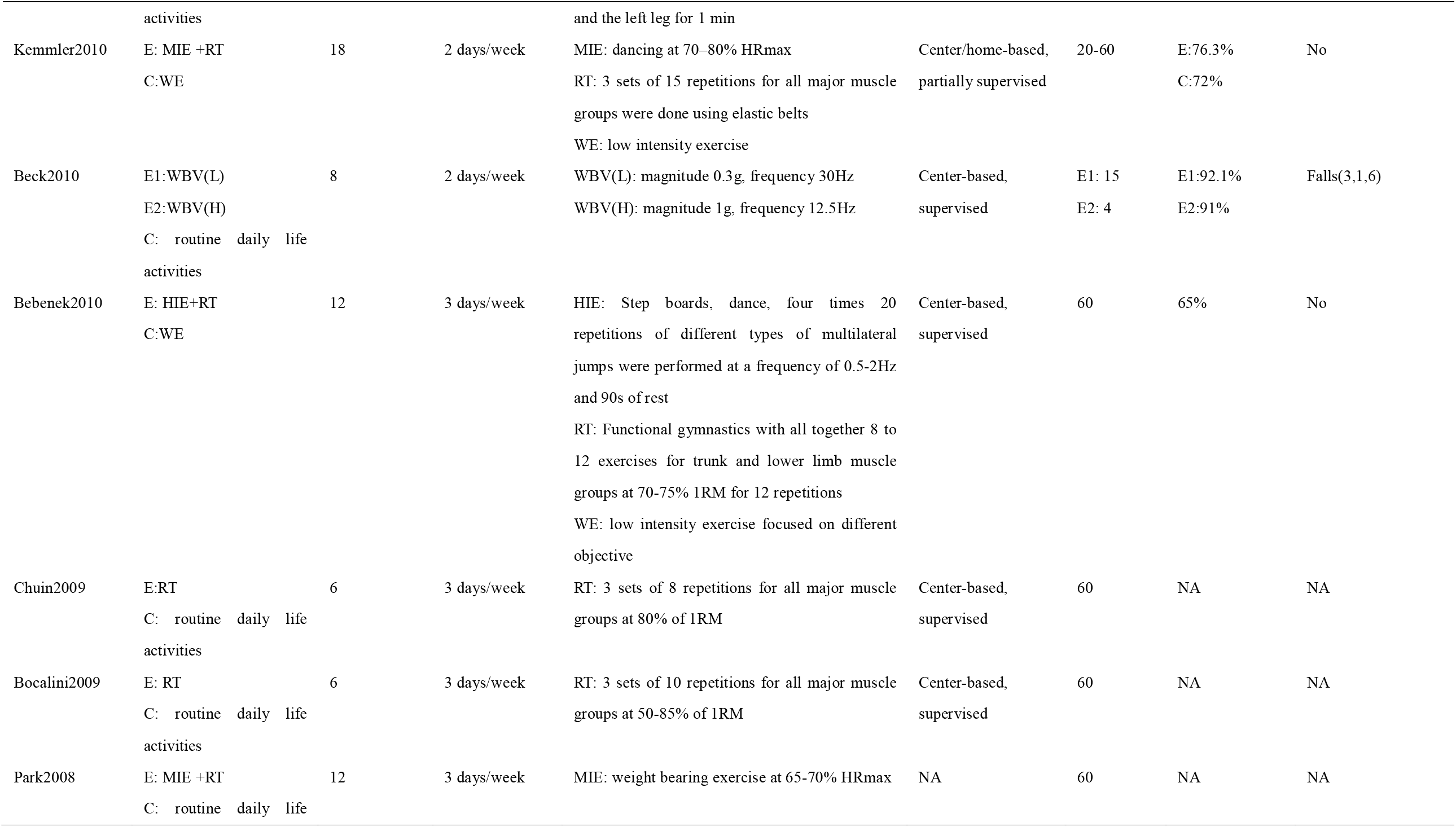
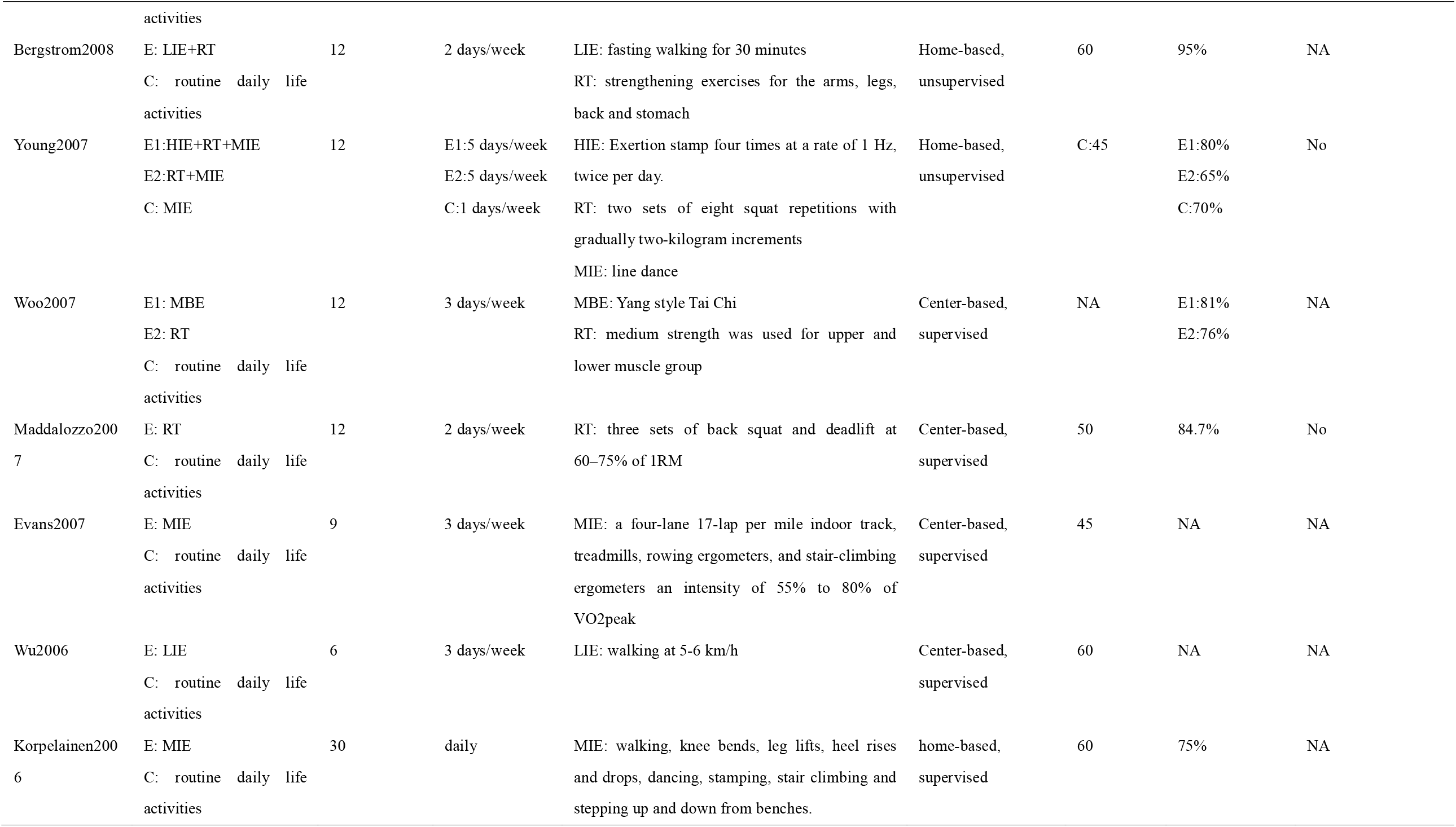
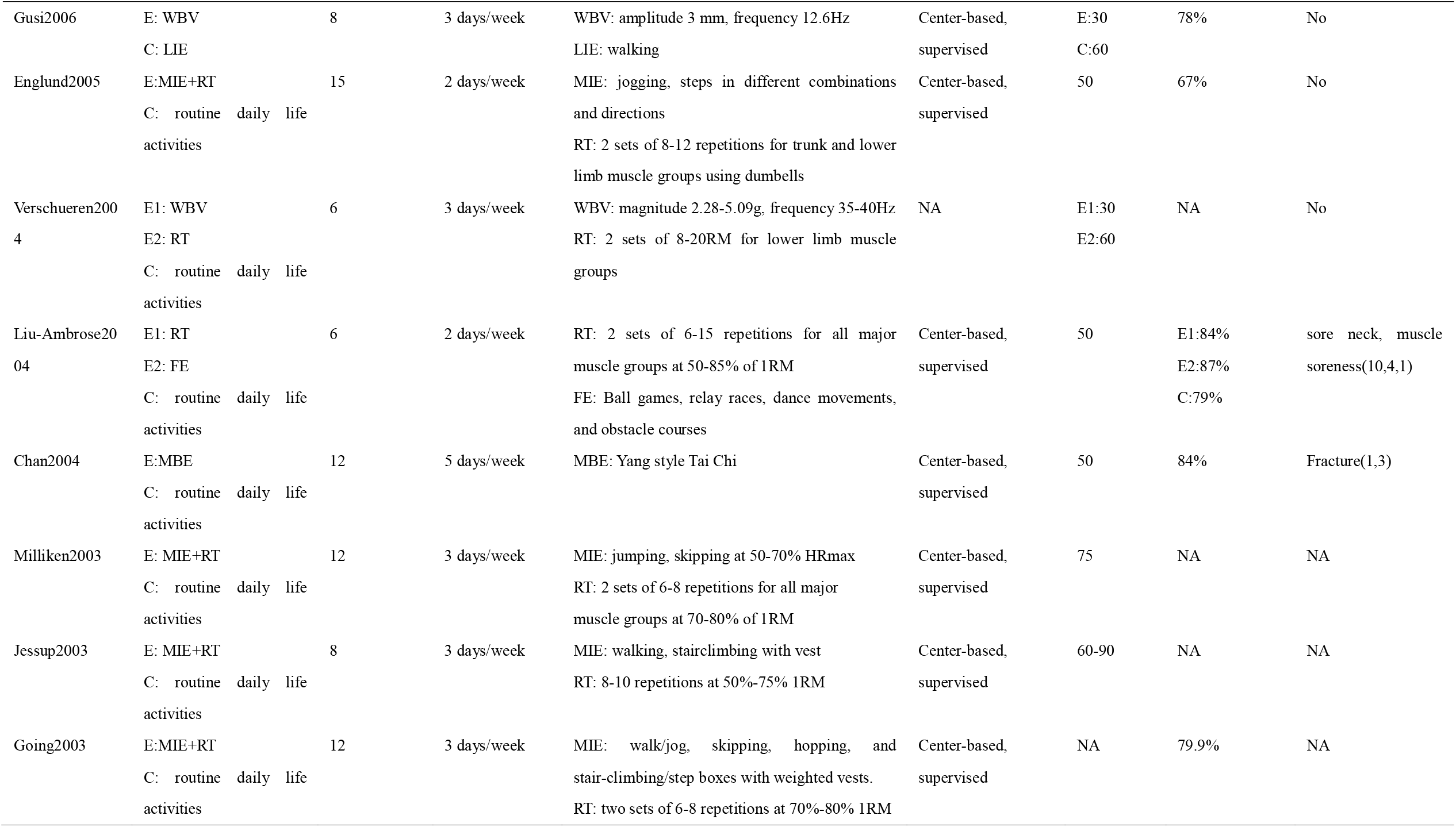
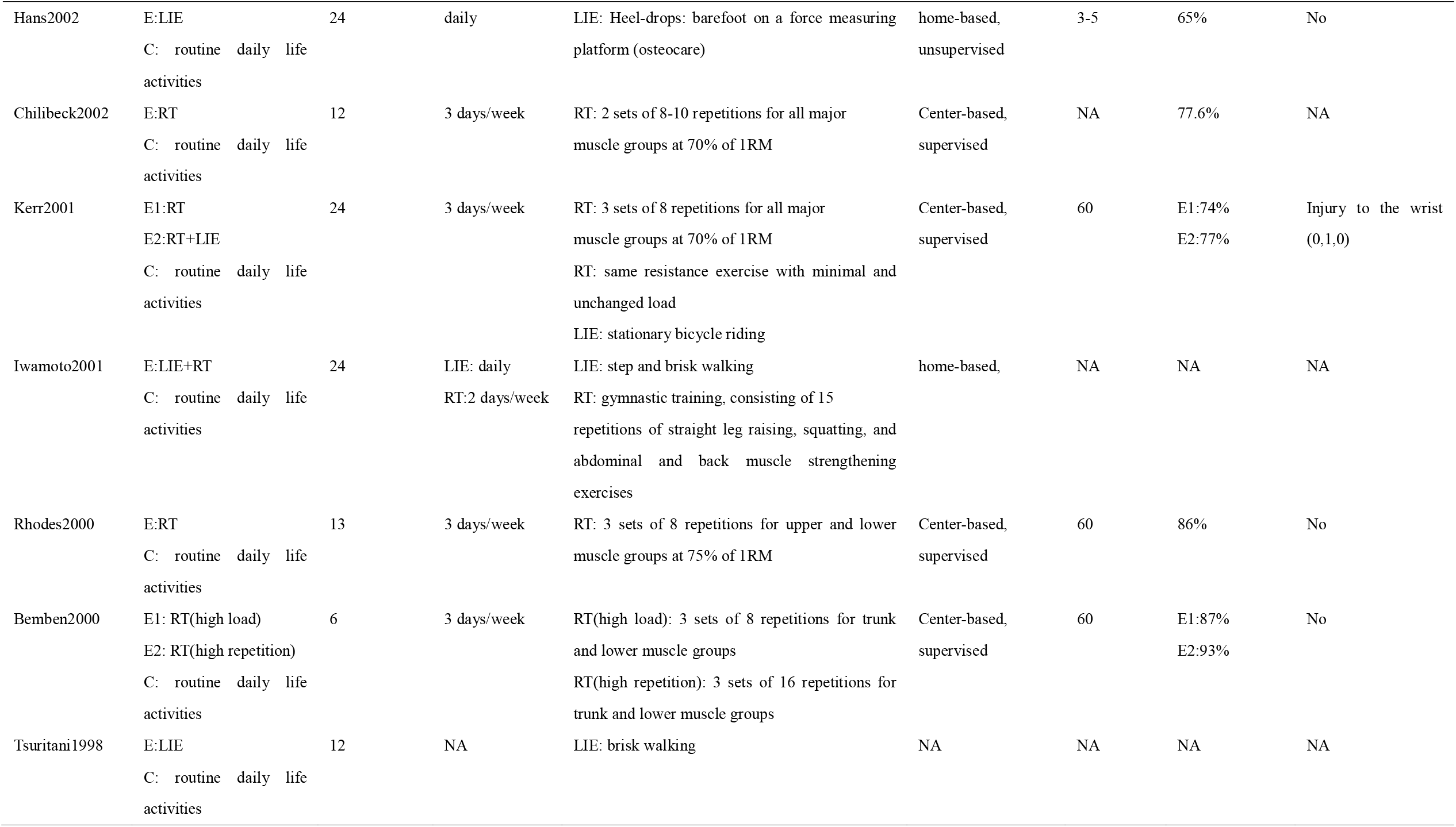
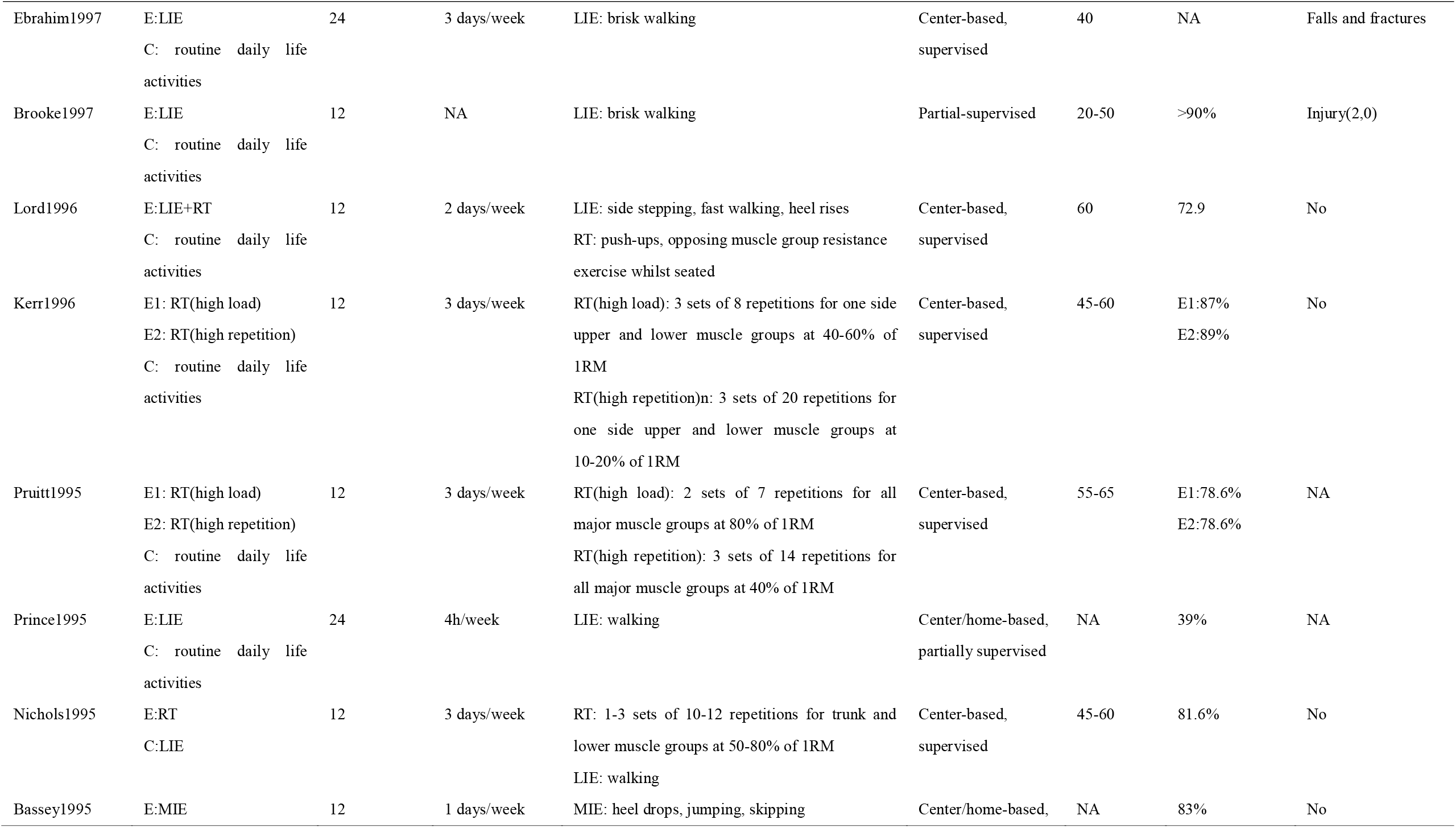
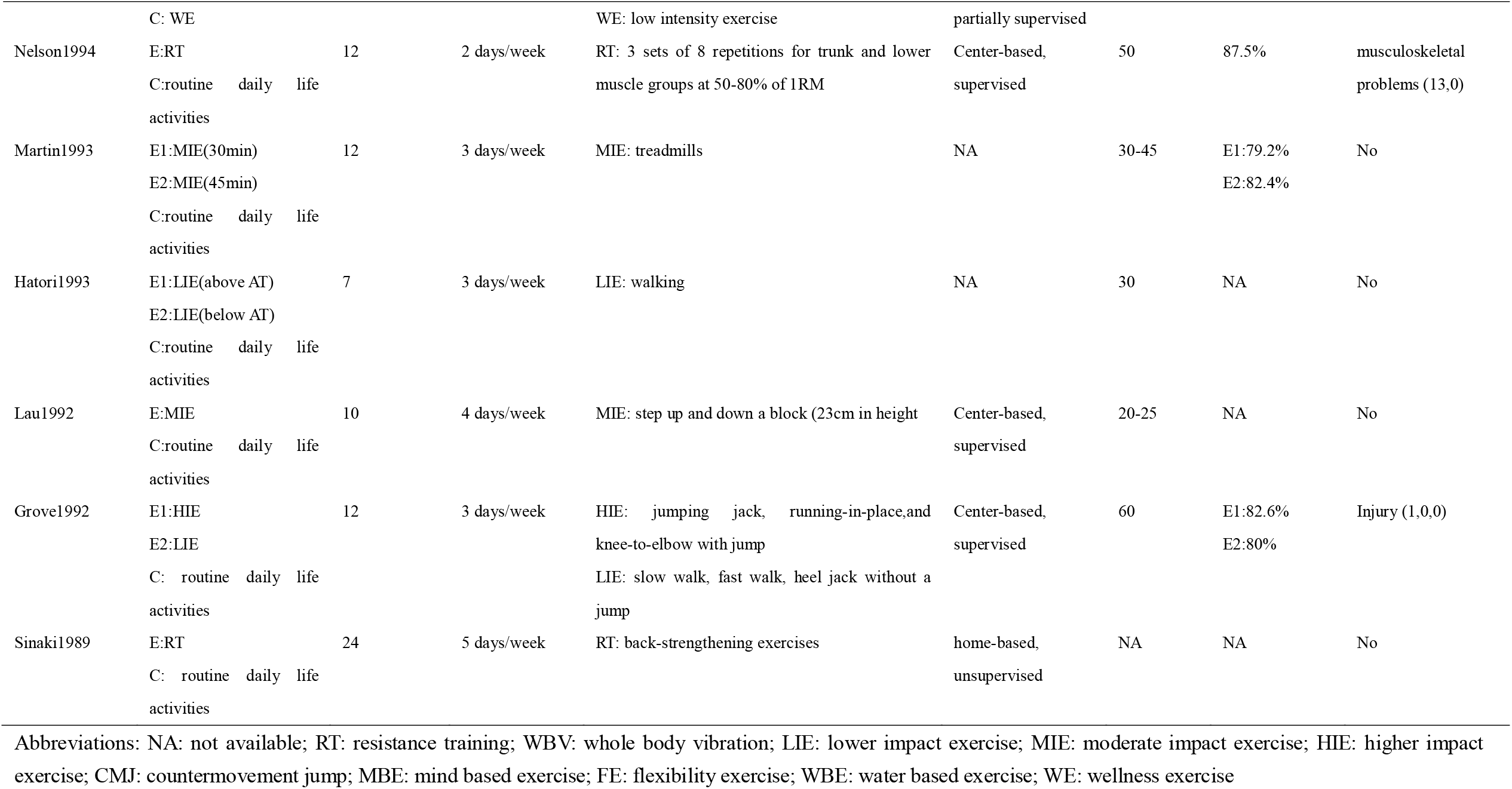
Exercise prescription characteristics of included studies

The PEDro scores of the included studies ranged from 3 to 8, with a mean score of 5·4. 11 (17%) studies can be considered as high, 42 (64%) were considered to be of moderate, and 13 (19%) of low quality. A detailed evaluation of the methodological quality is provided in Supplementary Appendix 5.

All models converged well. The parameter PSRF of all outcomes are close to 1, indicating a favorable convergence. No significant differences in comparison groups of all outcomes were found by inconsistency between direct and indirect estimates from the node splitting analysis (Supplementary Appendix 6). Supplementary Appendix 7 depicts the network diagram of eligible comparisons for BMD at LS (fifty eight studies with 3665 participants), FN (fifty studies with 3376 participants), and TH (twenty studies with 1664 participants), respectively.

Associated with BMD at LS improve were found for the multicomponent exercise 0·01 g/cm2, (95% CrI 0·01 to 0·02), resistance training 0·01 g/cm2, (95% CrI 0·00 to 0·02), mind body exercise 0·01 g/cm2, (95% CrI 0·00 to 0·02), lower impact exercise 0·01 g/cm2, (95% CrI 0·00 to 0·02), high impact exercise 0·03 g/cm2, (95% CrI 0·00 to 0·07), and whole body vibration 0·01 g/cm2, (95% CrI 0·0 to 0·02). After 12 studies of low quality were excluded, 25-36 there were no longer an association of high and lower impact exercise with improved BMD when compared with control (table 4).

**Table 4.** Relative effect estimates for the contrasts between the different interventions and control arms on BMD at LS

|  |  |  |  |  |  |  |  |  |  |  |
| --- | --- | --- | --- | --- | --- | --- | --- | --- | --- | --- |
| Control | **-0.02**<br>(-0.03,-0.01) | **-0.02**<br>(-0.02,-0.01) | 0.00<br>(-0.01,0.01) | -0.01<br>(-0.03,0.00) | **-0.01**<br>(-0.02,-0.00) | -0.04<br>(-0.07,0.00) | -0.01<br>(-0.03,0.00) | -0.01<br>(-0.02,0.00) | **-0.01**<br>(-0.02,-0.00) | -0.01<br>(-0.05,0.03) |
| **-0.01**<br>(-0.02,-0.00) | RT | 0.00<br>(-0.01,0.01) | **0.02**<br>(0.00,0.03) | 0.00<br>(-0.02,0.02) | 0.01<br>(-0.01,0.02) | -0.02<br>(-0.05,0.02) | 0.00<br>(-0.01,0.02) | 0.01<br>(-0.01,0.02) | 0.01<br>(-0.01,0.02) | 0.00<br>(-0.04,0.04) |
| **-0.01**<br>(-0.02,-0.01) | -0.00<br>(-0.01,0.01) | ME | **0.02**<br>(0.01,0.03) | 0.00<br>(-0.02,0.02) | 0.00<br>(-0.00,0.01) | -0.02<br>(-0.05,0.02) | 0.00<br>(-0.01,0.02) | 0.01<br>(-0.01,0.02) | 0.01<br>(-0.01,0.02) | 0.00<br>(-0.04,0.04) |
| 0.00<br>(-0.01,0.01) | **0.01**<br>(0.00,0.02) | **0.02**<br>(0.01,0.02) | WE | -0.02<br>(-0.03,0.00) | **-0.01**<br>(-0.02,-0.00) | -0.04<br>(-0.07,0.00) | -0.01<br>(-0.03,0.01) | -0.01<br>(-0.03,0.00) | -0.01<br>(-0.02,0.00) | -0.02<br>(-0.06,0.03) |
| -0.01<br>(-0.03,0.00) | -0.00<br>(-0.02,0.01) | 0.00<br>(-0.01,0.01) | -0.01<br>(-0.03,0.00) | FE | 0.01<br>(-0.01,0.02) | -0.02<br>(-0.06,0.02) | 0.00<br>(-0.02,0.03) | 0.01<br>(-0.01,0.03) | 0.00<br>(-0.01,0.02) | 0.00<br>(-0.04,0.05) |
| **-0.01**<br>(-0.02,-0.00) | 0.00<br>(-0.01,0.01) | 0.00<br>(-0.00,0.01) | **-0.01**<br>(-0.02,-0.00) | 0.00<br>(-0.01,0.02) | WBV | -0.03<br>(-0.06,0.01) | -0.00<br>(-0.02,0.02) | 0.00<br>(-0.01,0.01) | -0.00<br>(-0.01,0.01) | -0.00<br>(-0.05,0.04) |
| **-0.03**<br>(-0.07,-0.00) | -0.03<br>(-0.06,0.01) | -0.02<br>(-0.06,0.01) | **-0.04**<br>(-0.07,-0.00) | -0.02<br>(-0.06,0.02) | -0.03<br>(-0.06,0.01) | HIE | 0.02<br>(-0.02,0.06) | 0.03<br>(-0.01,0.06) | 0.03<br>(-0.01,0.06) | 0.02<br>(-0.04,0.08) |
| -0.01<br>(-0.02,0.01) | 0.00<br>(-0.01,0.02) | 0.01<br>(-0.00,0.02) | -0.01<br>(-0.02,0.01) | 0.01<br>(-0.01,0.03) | 0.00<br>(-0.01,0.02) | 0.03<br>(-0.01,0.07) | MIE | 0.00<br>(-0.02,0.02) | 0.00<br>(-0.02,0.02) | -0.00<br>(-0.05,0.04) |
| **-0.01**<br>(-0.02,-0.00) | -0.00<br>(-0.01,0.01) | 0.00<br>(-0.01,0.02) | -0.01<br>(-0.02,0.00) | 0.00<br>(-0.01,0.02) | -0.00<br>(-0.01,0.01) | 0.03<br>(-0.01,0.06) | -0.00<br>(-0.02,0.01) | LIE | -0.00<br>(-0.02,0.02) | -0.01<br>(-0.05,0.04) |
| **-0.01**<br>(-0.02,-0.00) | 0.00<br>(-0.01,0.01) | 0.00<br>(-0.01,0.01) | -0.01<br>(-0.02,0.00) | 0.00<br>(-0.01,0.02) | 0.00<br>(-0.01,0.01) | 0.03<br>(-0.01,0.06) | -0.00<br>(-0.02,0.01) | 0.00<br>(-0.01,0.01) | MBE | -0.00<br>(-0.04,0.04) |
| -0.02<br>(-0.06,0.03) | -0.00<br>(-0.05,0.04) | -0.00<br>(-0.04,0.04) | -0.02<br>(-0.06,0.03) | -0.00<br>(-0.05,0.04) | -0.01<br>(-0.05,0.04) | 0.02<br>(-0.03,0.07) | -0.01<br>(-0.05,0.04) | -0.00<br>(-0.05,0.04) | -0.01<br>(-0.05,0.04) | WBE |

Summary estimates from the network meta-analysis are shown in lower left triangle, and summary estimates after excluding low quality studies in upper right triangle. Each cell shows a mean difference (MD), with a 95 % CI in parentheses . For any cell, a negative MD favours the lower-right intervention, and a positive SMD favours the upper-left intervention. Significant results in bold text.

Among the high quality trials, high impact exercise had the highest probability of being the best exercise intervention (SUCRA value of 0·88 compared with 0·72 for resistance training and 0·66 for multicomponent exercise). However, there were only three trials for this intervention and the results were limited by wide 95% CrIs. When all trials were included, high impact exercise was also the best exercise intervention (0 · 91) followed by multicomponent exercise (0·71) and flexibility exercise (0·61) (Supplementary Appendix 8).

Only multicomponent exercise (0·03 g/cm2, 95% CrI 0·01 to 0·04), whole body vibration (0·02 g/cm2, 95% CrI 0·00 to 0·03), and mind body exercise (0·02 g/cm2, 95% CrI 0·00 to 0·03) were effective for improving BMD at FN. Moreover, multicomponent exercise yielded significant benefits relative to resistance training, and moderate impact exercise. After nine studies of low quality were excluded, ^25, 26, 28-30, 32-34, 37^, the effect size of multicomponent exercise versus lower impact exercise turned to increase from 0·02 g/cm2 to 0·03 g/cm2, with 95% CrI of 0·01 to 0·05, while the other findings kept consistent with the prior data (table 5).

**Table 5.** Relative effect estimates for the contrasts between the different interventions and control arms on BMD at FN

|  |  |  |  |  |  |  |  |  |  |  |
| --- | --- | --- | --- | --- | --- | --- | --- | --- | --- | --- |
| Control | -0.01<br>(-0.02,0.00) | **-0.03**<br>**(-0.05,-0.02)** | -0.02<br>(-0.04,0.00) | -0.01<br>(-0.04,0.01) | **-0.02**<br>**(-0.03,-0.00)** | -0.00<br>(-0.04,0.04) | 0.00<br>(-0.02,0.02) | -0.01<br>(-0.02,0.01) | **-0.02**<br>**(-0.03,-0.00)** | -0.02<br>(-0.07,0.02) |
| -0.01<br>(-0.02,0.00) | RT | **-0.02**<br>**(-0.04,-0.00)** | -0.01<br>(-0.03,0.02) | -0.00<br>(-0.03,0.02) | -0.01<br>(-0.03,0.01) | 0.01<br>(-0.04,0.05) | 0.01<br>(-0.01,0.04) | 0.00<br>(-0.02,0.02) | -0.01<br>(-0.03,0.01) | -0.01<br>(-0.06,0.03) |
| **-0.03**<br>**(-0.04,-0.01)** | **-0.02**<br>**(-0.03,-0.00)** | ME | 0.01<br>(-0.01,0.04) | 0.02<br>(-0.00,0.04) | 0.01<br>(-0.00,0.03) | 0.03<br>(-0.01,0.07) | **0.04**<br>**(0.01,0.06)** | **0.03**<br>**(0.01,0.05)** | 0.02<br>(-0.01,0.04) | 0.01<br>(-0.03,0.05) |
| -0.01<br>(-0.03,0.01) | -0.00<br>(-0.02,0.02) | 0.02<br>(-0.00,0.04) | WE | 0.00<br>(-0.02,0.04) | -0.00<br>(-0.02,0.02) | 0.02<br>(-0.03,0.06) | 0.02<br>(-0.01,0.05) | 0.01<br>(-0.01,0.04) | 0.00<br>(-0.03,0.03) | 0.01<br>(-0.05,0.04) |
| -0.01<br>(-0.03,0.01) | -0.00<br>(-0.02,0.02) | 0.02<br>(-0.01,0.04) | -0.00<br>(-0.03,0.03) | FE | -0.00<br>(-0.03,0.02) | 0.01<br>(-0.03,0.06) | 0.02<br>(-0.01,0.05) | 0.01<br>(-0.02,0.03) | -0.00<br>(-0.03,0.02) | -0.01<br>(-0.06,0.04) |
| **-0.02**<br>**(-0.03,-0.00)** | -0.01<br>(-0.03,0.01) | 0.01<br>(-0.01,0.03) | -0.01<br>(-0.03,0.01) | -0.01<br>(-0.03,0.02) | WBV | 0.02<br>(-0.03, 0.06) | 0.02<br>(-0.00, 0.05) | 0.01<br>(-0.01, 0.03) | 0.00<br>(-0.02, 0.02) | -0.01<br>(-0.05, 0.04) |
| -0.00<br>(-0.04,0.04) | 0.00<br>(-0.04,0.05) | 0.02<br>(-0.02,0.06) | 0.01<br>(-0.04,0.05) | 0.01<br>(-0.04,0.05) | 0.01<br>(-0.03,0.06) | HIE | 0.01<br>(-0.04, 0.05) | -0.01<br>(-0.05, 0.04) | -0.02<br>(-0.06, 0.03) | -0.02<br>(-0.08, 0.04) |
| 0.00<br>(-0.01,0.02) | 0.01<br>(-0.01,0.03) | **0.03**<br>**(0.01,0.05)** | 0.01<br>(-0.01,0.03) | 0.01<br>(-0.01,0.04) | 0.21<br>(-0.00,0.04) | 0.01<br>(-0.04,0.05) | MIE | -0.01<br>(-0.04, 0.01) | -0.02<br>(-0.05, 0.01) | -0.03<br>(-0.07, 0.02) |
| -0.01<br>(-0.02,0.01) | -0.00<br>(-0.02,0.02) | 0.02<br>(-0.00,0.04) | 0.00<br>(-0.02,0.02) | 0.00<br>(-0.02,0.03) | 0.01<br>(-0.01,0.03) | -0.01<br>(-0.05,0.04) | -0.01<br>(-0.03,0.01) | LIE | -0.01<br>(-0.03, 0.01) | -0.02<br>(-0.06, 0.03) |
| **-0.02**<br>**(-0.03,-0.00)** | -0.01<br>(-0.03,0.01) | 0.01<br>(-0.01,0.03) | -0.01<br>(-0.03,0.02) | -0.01<br>(-0.03,0.02) | -0.00<br>(-0.02,0.02) | -0.01<br>(-0.06,0.03) | -0.02<br>(-0.04,0.00) | -0.01<br>(-0.03,0.01) | MBE | -0.01<br>(-0.05, 0.04) |
| -0.02<br>(-0.06,0.02) | -0.02<br>(-0.06,0.03) | 0.00<br>(-0.04,0.04) | -0.01<br>(-0.06,0.03) | -0.01<br>(-0.06,0.03) | -0.01<br>(-0.05,0.04) | -0.02<br>(-0.08,0.04) | -0.03<br>(-0.07,0.02) | -0.02<br>(-0.06,0.03) | -0.01<br>(-0.05,0.04) | WBE |

Summary estimates from the network meta-analysis are shown in lower left triangle, and summary estimates after excluding low quality studies in upper right triangle. Each cell shows a mean difference (MD), with a 95 % CI in parentheses . For any cell, a negative MD favours the lower-right intervention, and a positive SMD favours the upper-left intervention. Significant results in bold text.

Among the high quality trials, multicomponent exercise had the highest probability of being the best exercise intervention (0·94 compared with 0·68 for water based exercise and 0·61 for whole body vibration). When all trials were included, the SUCRA and rankogram plots followed a similar pattern (Supplementary Appendix 9).

Only multicomponent exercise (0·01 g/cm2, 95% CrI 0·00 to 0.01), resistance training (0·01 g/cm2, 95% CrI 0·00 to 0·02), and flexibility exercise (0·01 g/cm2, 95% CrI 0·00 to 0·02) were effective for improving BMD at TH. Moreover, resistance training, and flexibility exercise yielded significant benefits versus whole body vibration. After two studies of low quality were excluded, ^27, 34^ multicomponent exercise was associated with improved BMD at TH compared with whole body vibration, while the other findings kept consistent with the prior data (Supplementary Appendix 11).

Among the high quality trials, resistance training had the highest probability of being the best exercise intervention (0·85 compared with 0·8 for flexibility exercise and 0·72 for multicomponent exercise). When all trials were included, the SUCRA and rankogram plots followed a similar pattern (Supplementary Appendix 10).

For participants with age no less than 60, whole body vibration, lower impact exercise, resistance training and multicomponent exercise were beneficial for improving BMD at LS (Supplementary Appendix 12.1). With regard to BMD at FN, multicomponent exercise was superior to control, moderate impact exercise and resistance training (Supplementary Appendix 12.2). No exercise was found effective for BMD at TH in participants with age no less than 60 (Supplementary Appendix 12.3).

For participants younger than 60 years old, mind body exercise, resistance training, multicomponent exercise, and high impact exercise were beneficial for improving BMD at LS. With regard to BMD at FN, mind body exercise, multicomponent exercise, and flexibility exercise were superior to control (Supplementary Appendix 14.2). No exercise was found effective for BMD at TH in participants younger than 60 years old (Supplementary Appendix 14.3). Supplementary Appendix 13 and 15 showed the bayesian ranking profiles of comparable treatments in different populations.

For exercise intervention with duration no more than eight months, whole body vibration, mind based exercise, resistance training, and multicomponent exercise were beneficial for improving BMD at LS (Supplementary Appendix 16.1). With regard to BMD at FN, multicomponent exercise was superior to control, lower and moderate impact exercise, resistance training, as well as flexibility exercise (Supplementary Appendix 16.2). No exercise was found effective for BMD at TH when the intervention duration was no more than eight months (Supplementary Appendix 16.3).

For exercise intervention with duration between 9 to 18 months, resistance training, high impact exercise, flexibility exercise and multicomponent exercise were beneficial for improving BMD at LS (Supplementary Appendix 18.1). With regard to BMD at FN, mind body exercise was superior to control, whereas resistance training produced significant improvement for BMD at TH (Supplementary Appendix 18.3). Supplementary Appendix 17 and 19 showed the bayesian ranking profiles of comparable treatments in different intervention duration.

No subgroup analyses could be performed for the BMD at LS, FN, and TH with duration of intervention more than 18 months due to insufficient evidence.

Supplementary Appendix 20 summarizes the certainty of evidence for the three networks (details on the CINeMA assessment of each pairwise comparisons are provided in appendix 21.1-3). The comparison-adjusted funnel plots were visually inspected. No apparent publication bias was found for BMD at LS and FN, except for BMD at TH (Supplementary Appendix 22).

## Discussion

In this NMA, we comprehensively display the comparative effect of all available exercise interventions on bone mineral density in postmenopausal women. According to the CINeMA criteria, the quality of evidence was low to very low, suggesting that findings should be interpreted with caution. The results indicate that: (1) depending on outcome of interest, multicomponent exercise (for BMD at LS,FN), and resistance training (for BMD at TH) were the most effective interventions; (2) Multicomponent exercise improved all the outcomes to a varying degree; (3) No matter the age of postmenopausal women, and the duration of intervention (range between 6 to 18 months), some certain kinds of exercise could be performed to improve BMD at LS and FN.

Lack of physical activity is related to decreased bone mass, whereas exercise involving bone loading could promote the increase of bone mass. Therefore, exercise has been proposed as a promising method to manage osteoporosis. Yet even exercise is recommended by multiple international guidelines,^38-40^ the magnitude of benefit of exercise intervention has been considered as moderate at best,^11, 12, 41^ which may be partially due to the large variety of exercise regimens and participant characteristics. Our results align with previous systematic review and meta-analyses where protocols including different intensity of impact exercise, as well as protocols including impact exercise with resistance training, were effective at preventing bone density loss at LS and FN.^42, 43^ Also, our results is consistent with the results of Kemmler,^44^ where resistance training tends to have larger effect size than impact exercise and multicomponent exercise. According to previous evidence, high intensity resistance training typically has beneficial effects only on the spine.^45^ And impact exercise, such as skipping, hopping, constantly attenuated before transmission to the spine, resulting in an osteogenic stimulus merely for the hips.^46^ Hence, single component training intervention generally produces a site-specific effect on bone. It also has been observed that adaptive skeletal response necessitates dynamic rather than static mechanical stimulation, and exercise intensity, stimulation frequency, as well as mechanical loading pattern are all crucial elements for enhancing skeletal response.^47^ Accordingly, multicomponent exercise integrating all the crucial elements may be the optimum choice for improving postmenopausal bone health. This may be the reason why multicomponent exercise could improve all the outcomes to a varying degree.

The results of subgroup analysis based on the age of participants are different from those of Rahimi et al.^48^ The possible reason for these divergent results may be caused by the number of included studies, and different inclusion criteria, such as controlled trial, language, additional supplement of calcium and vitamin D. Apart from the beneficial effects of mechanical stimulus on bone itself, older women were still sensitive to exercise maybe partially due to an increase of calcium absorption caused by exercise.^49^

Typically, prolonged intervention duration should result in greater bone benefit, at least when strain was periodically progressed. According to the view that exercise-induced changes of BMD were primarily caused by remodeling, and taking into account the length of a remodeling cycle in adults, interventions less than 8 months may be insufficient to determine the full amount of new mineral bone.^23^ However, the subgroup analysis demonstrated considerably higher effects of several exercises on BMD at LS and FN among studies with short compared with longer duration. Moreover, participants seemed to be more sensitive to multicomponent exercise at LS and FN. We attributed this suspicious finding to the complex interaction of exercise parameters.

To the best of our knowledge, this is the first network systematic review to compare and rank different exercise interventions on BMD in postmenopausal women. We conducted a thorough search of multiple databases and sources to identify eligible studies without language restriction. All the process during literature search, data extraction and methodological quality assessment were performed by a pair of independent reviewers to minimize potential bias.

Nevertheless, several noteworthy limitations in this network meta-analysis should be addressed. A significant limitation is that we were not entirely relied on author description of exercise and control groups for classification. Exercise intervention and group are rarely standardized and vary substantially among studies. Therefore, based on description of the article and the definition of exercise according to our criteria, we categorized the exercise intervention. Besides, although strict inclusion criteria was set to make the interventions were reasonably similar among studies, the included treatments still contained various exercise interventions. It suggests that further disassemble of the specific physical elements that constitute an effective combined intervention should be investigated.

Second, despite our attempt to contact the related authors, some outcome data remain unavailable. Therefore, a few of the SDs of the absolute change in BMD was calculated according to specific formula, which may affected the accuracy of the data. Additionally, substantial publication bias was found in studies investigating the effect of exercise in BMD at TH. Given the nature that authors tend to report positive results, the actual effect of exercise on BMD could be somewhat lower than the values reported here.

Third, the primary outcome of our study is the change of BMD, however, it only accounts for about 60% of variation in bone strength. Other surrogate parameter like microarchitecture were not included. Moreover, for the analysis of results, only the effect sizes at the end of the intervention were extracted rather than those at further follow-up times, which can be criticized. However, we believe that just as medication, exercise intervention are most beneficial during the duration of implementation. Therefore, the effect sized post intervention were considered the most relevant.

Last, PEDro scale is used to evaluate methodological quality, it may underestimate risk of bias of included studies when compared with Cochrane Risk of Bias 2 tool. ^50^ Furthermore, since the overall very low graded evidence, the relative treatment estimates may alter as a more high-quality evidence obtained.

## Data Availability

All data, models, and code generated or used during the study appear in the submitted article.

## Contributors

HCQ obtained funding for the study and LY, ZSY and HCQ designed it. WXY and ZYS screened the literature search, acquired reports of relevant trials, selected included studies. WLQ and WT extracted data. XSH,GXL and HJM did all statistical analyses. HCQ, ZSY and LY analysed and interpreted the data. LY and HCQ drafted the report. All the authors critically reviewed the report and approved the final submitted version.

