## supplementary material for "Comparative effectiveness of different exercises on bone mineral density in postmenopausal women: a systematic review and network meta-analysis of randomized controlled trials"

### Appendix 1: MEDLINE search strategy

1 osteoporosis/

2 osteoporo$.mp.

3 osteopenia.mp.

4 bone density/

5 bone densit$.mp.

6 bone loss$.mp.

7 bone mass$.mp.

8 bone mineral densit$.mp.

9 bone mineral content$.mp.

10 bone age.mp.

11 bone defect$.mp.

12 bone deminerali?ation.mp.

13 bone mineral$.mp.

14 bone strength.mp.

15 decalcifi$.mp.

16 deminerali?ed bone.mp.

17 or/1-16

18 exp Menopause/

19 (menopaus$ or postmenopaus$ or post-menopaus$ or (post adj menopause$)).mp.

20 18 or 19

21 17 and 20

22 exp exercise/

23 exp exercise therapy/

24 exerci$.mp.

25 exp sports/

26 sport$.mp.

27 physical fitness/

28 physical fitness.mp.

29 physical activit$.mp.

30 vibration/tu

31 vibration therap$.mp.

32 22 OR 23 OR 24 OR 25 OR 26 OR 27 OR 28 OR 29 OR 30 OR 31

33 randomized controlled trial.pt.

34 controlled clinical trial.pt.

35 randomi?ed.ab.

36 placebo.ab.

37 clinical trials as topic.sh.

38 randomly.ab.

39 trial.ti.

40 33 OR 34 OR 35 OR 36 OR 37 OR 38 OR 39

41 21 AND 32 AND 40

### Appendix 2: Statistical methods in detail

**Calculation of change score**

If the studies presented a confidence interval (CI) or standard errors (SE), they were converted to standard deviation (SD) by using the following formula: SD=$SE\times\sqrt{N}$. ^3 2 1 1^ When the SDs of absolute changes from baseline were not available from individual trials, we imputed them as described in detail in the Cochrane Handbook: ${SD}_{change}=\sqrt{{{SD}^{2}}_{pre}+{{SD}^{2}}_{post}-(2\times R\times{SD}_{pre}\times{SD}_{post})}$. ^3 2 1 1^ R refers to correlation coefficient which was calculated using the mean of the correlations available for some included studies. Therefore, the value was estimated of r=0·82 and r=0·76 in exercise and control groups at LS, respectively. It also resulted in a within-group correlation of r=0·85 for the exercise group and r=0·96 among control groups at FN. Finally, at TH, r=0·98 and r=0·99 were considered for intervention and control groups, respectively. Using the imputed correlation coefficient values, we thereafter derived unavailable ${SD}_{change}$ from calculated ${SD}_{change}$.

**Assessment for convergence**

We evaluated convergence according to the Brooks-Gelman-Rubin method. This method calculates the possible scale reduction factor (PSRF) by comparing within- and between-chain variation. Convergence is deemed favorable if PSRF is close to one, and the consistency of the homogeneity model was considered reliable for further analysis. With regard to the inconsistency model, if there is no inconsistency, the random effects variance and inconsistency variance should be roughly equal. If the latter one is bigger, that indicates a problem.^4^ The node splitting method was used to further examine the inconsistency between the direct and indirect evidence (with high p values indicating better consistency).^5^

### Appendix 3: Flowchart of literature search and study selection.

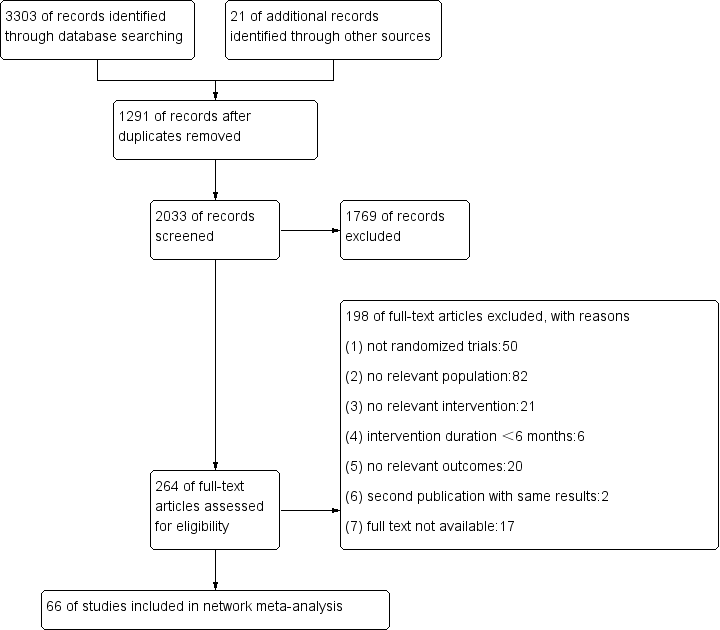

### Appendix 4: Details of included and excluded studies

**2.1 list of include studies**

6.	Wang H, Yu B, Chen W, Lu Y, Yu D. Simplified Tai Chi resistance training versus traditional Tai Chi in slowing bone loss in postmenopausal women. Evidence-based Complementary and Alternative Medicine. 2015;**2015 (no pagination)**.

**2.2 list of excluded studies**

**Not randomized trials (n=50) :**

**No relevant population (n=82):**

63.	Bailey, C.A. and K. Brooke-Wavell, Daily exercise is most effective for increasing hip bone mineral density: A randomized high-impact, unilateral intervention. Bone, 2009. 44: p. S100-S101.

70.	Effect of eight weeks aerobic training on serum parathormone, estrogen and alkaline phosphatase concentration in obese women with premature menopause. Iranian journal of obstetrics, gynecology and infertility. 20(7): p. 8-17.

**No relevant intervention (n=21):**

**Intervention duration＜6 months (n=6)：**

**No relevant outcome (n=20):**

**Second publication with same results (n=2):**

1.	Watson, S.L., et al., Heavy resistance training is safe and improves bone, function, and stature in postmenopausal women with low to very low bone mass: novel early findings from the LIFTMOR trial. Osteoporosis International, 2015. 26(12): p. 2889-94.

2.	Bergstrom, I., et al., Physical training increases osteoprotegerin in postmenopausal women. Journal of Bone & Mineral Metabolism, 2012. 30(2): p. 202-7.

**Full text not available (n=17):**

1.	Effects of exercise on bone mineral density of the lumbar spine in postmenopausal women. Annales de readaptation ET de medecine physique.38(3):117-24.

2.	Lumbar bone loss and compliance of postmenopausal, psoas-trained women: a 3-year follow-up study. Annales de readaptation ET de medecine physique.41(4):203-8.

### Appendix 5: PEDro assessment quality results of included studies

| **Appendix 3 PEDro assessment of the quality of the results in the included studies** | | | | | | | | | | | | | |
| --- | --- | --- | --- | --- | --- | --- | --- | --- | --- | --- | --- | --- | --- |
| Study | Eligibility criteria | Random  allocation | Concealed  allocation | Similar  groups  at baseline | Blinding  subjects | Blinding  therapists | Blinding  assessors | Outcome obtained in more than 85% of the subjects | Intention-to-treat analysis | Between-group  statistical comparison | Point estimates and measures of variability | Total | Quality |
| Sen 2020 | YES | 1 | 0 | 1 | 0 | 0 | 0 | 1 | 1 | 1 | 1 | 6 | M |
| Montgomery 2020 | YES | 1 | 0 | 1 | 0 | 0 | 1 | 0 | 1 | 1 | 1 | 6 | M |
| ElDeeb 2020 | YES | 1 | 1 | 1 | 0 | 0 | 0 | 1 | 0 | 1 | 1 | 6 | M |
| de Oliveira 2019 | YES | 1 | 1 | 1 | 0 | 0 | 0 | 1 | 1 | 1 | 1 | 7 | H |
| Duff2016 | YES | 1 | 1 | 1 | 1 | 0 | 1 | 0 | 1 | 1 | 1 | 8 | H |
| Wang2015 | YES | 1 | 0 | 1 | 0 | 0 | 0 | 1 | 0 | 1 | 1 | 5 | M |
| Nicholson2015 | YES | 1 | 0 | 0 | 0 | 0 | 1 | 1 | 1 | 1 | 1 | 6 | M |
| Liu2015 | YES | 1 | 0 | 1 | 0 | 0 | 0 | 1 | 0 | 1 | 1 | 5 | M |
| Moreira2014 | YES | 1 | 0 | 1 | 0 | 0 | 0 | 1 | 0 | 1 | 1 | 5 | M |
| Lai 2013 | YES | 1 | 0 | 1 | 0 | 0 | 0 | 1 | 0 | 1 | 1 | 5 | M |
| Kemmler2013 | YES | 1 | 0 | 1 | 0 | 0 | 1 | 0 | 1 | 1 | 1 | 6 | M |
| Chilibeck 2013 | YES | 1 | 1 | 1 | 0 | 0 | 1 | 1 | 1 | 1 | 1 | 8 | H |
| Basat 2013 | YES | 1 | 0 | 1 | 0 | 0 | 0 | 1 | 0 | 1 | 1 | 5 | M |
| Orsatti 2013 | YES | 1 | 0 | 0 | 0 | 0 | 0 | 1 | 0 | 1 | 1 | 4 | L |
| Wayne2012 | YES | 1 | 0 | 1 | 0 | 0 | 1 | 1 | 1 | 1 | 1 | 7 | H |
| Karakiriou2012 | YES | 1 | 0 | 1 | 0 | 0 | 1 | 1 | 0 | 1 | 1 | 6 | M |
| Bolton2012 | YES | 1 | 1 | 1 | 0 | 0 | 1 | 1 | 0 | 1 | 1 | 7 | H |
| von Stengel 2011 | YES | 1 | 1 | 1 | 0 | 0 | 0 | 1 | 1 | 1 | 1 | 7 | H |
| von Stengel 2011(1) | YES | 1 | 0 | 1 | 1 | 0 | 1 | 1 | 1 | 1 | 1 | 8 | H |
| Tartibian2011 | No | 1 | 0 | 1 | 0 | 0 | 0 | 1 | 0 | 1 | 1 | 5 | M |
| Slatkovska2011 | YES | 1 | 1 | 1 | 0 | 0 | 1 | 1 | 1 | 1 | 1 | 8 | H |
| Marques 2011 | YES | 1 | 1 | 1 | 0 | 0 | 1 | 0 | 1 | 1 | 1 | 7 | H |
| Marques 2011(1) | YES | 1 | 1 | 1 | 0 | 0 | 0 | 0 | 1 | 1 | 1 | 6 | M |
| Sakai2010 | No | 1 | 0 | 1 | 0 | 0 | 0 | 1 | 0 | 1 | 1 | 5 | M |
| Kemmler2010 | YES | 1 | 0 | 1 | 0 | 0 | 1 | 1 | 1 | 1 | 1 | 7 | H |
| Beck2010 | YES | 1 | 0 | 0 | 0 | 0 | 1 | 1 | 1 | 1 | 1 | 6 | M |
| Bebenek 2010 | YES | 1 | 0 | 1 | 0 | 0 | 1 | 0 | 1 | 1 | 1 | 6 | M |
| Chuin2009 | YES | 1 | 0 | 1 | 0 | 0 | 0 | 1 | 0 | 1 | 1 | 5 | M |
| Bocalini2009 | YES | 1 | 0 | 1 | 0 | 0 | 0 | 0 | 0 | 1 | 1 | 4 | L |
| Park2008 | YES | 1 | 0 | 1 | 0 | 0 | 0 | 1 | 0 | 1 | 1 | 5 | M |
| Bergstrom 2008 | YES | 1 | 0 | 1 | 0 | 0 | 0 | 1 | 1 | 1 | 1 | 6 | M |
| Young2007 | YES | 1 | 0 | 1 | 0 | 0 | 0 | 0 | 0 | 1 | 1 | 4 | L |
| Woo2007 | YES | 1 | 0 | 1 | 0 | 0 | 1 | 1 | 0 | 1 | 1 | 6 | M |
| Maddalozzo 2007 | YES | 1 | 0 | 1 | 0 | 0 | 0 | 1 | 0 | 1 | 1 | 5 | M |
| Evans2007 | YES | 1 | 0 | 1 | 0 | 0 | 0 | 0 | 1 | 1 | 1 | 5 | M |
| Wu2006 | YES | 1 | 0 | 1 | 0 | 0 | 0 | 1 | 0 | 1 | 1 | 5 | M |
| Korpelainen2006 | YES | 1 | 0 | 1 | 0 | 0 | 1 | 1 | 1 | 1 | 1 | 7 | H |
| Gusi2006 | YES | 1 | 0 | 1 | 0 | 0 | 0 | 0 | 1 | 1 | 1 | 5 | M |
| Englund2005 | YES | 1 | 0 | 1 | 0 | 0 | 0 | 1 | 0 | 1 | 1 | 5 | M |
| Verschueren2004 | YES | 1 | 0 | 1 | 0 | 0 | 1 | 1 | 0 | 1 | 1 | 6 | M |
| Liu-Ambrose2004 | YES | 1 | 0 | 1 | 0 | 0 | 1 | 1 | 0 | 1 | 1 | 6 | M |
| Chan2004 | YES | 1 | 0 | 1 | 0 | 0 | 0 | 1 | 0 | 1 | 1 | 5 | M |
| Milliken 2003 | YES | 1 | 0 | 0 | 0 | 0 | 0 | 1 | 0 | 1 | 1 | 4 | L |
| Jessup 2003 | YES | 1 | 0 | 0 | 0 | 0 | 1 | 1 | 0 | 1 | 1 | 5 | M |
| Going2003 | YES | 1 | 0 | 1 | 0 | 0 | 0 | 0 | 1 | 1 | 1 | 5 | M |
| Hans2002 | YES | 1 | 1 | 1 | 0 | 0 | 0 | 1 | 0 | 1 | 1 | 6 | M |
| Chilibeck2002 | YES | 1 | 0 | 1 | 0 | 0 | 0 | 1 | 0 | 1 | 1 | 5 | M |
| Kerry2001 | YES | 1 | 0 | 1 | 0 | 0 | 0 | 0 | 0 | 1 | 1 | 4 | L |
| Iwamoto2001 | No | 1 | 0 | 1 | 0 | 0 | 0 | 1 | 0 | 1 | 1 | 5 | M |
| Rhodes2000 | YES | 1 | 0 | 1 | 0 | 0 | 0 | 1 | 0 | 1 | 1 | 5 | M |
| Bemben 2000 | YES | 1 | 0 | 1 | 0 | 0 | 0 | 1 | 0 | 1 | 1 | 5 | M |
| Tsuritani1998 | No | 1 | 0 | 0 | 0 | 0 | 0 | 0 | 0 | 1 | 1 | 3 | L |
| Ebrahim 1997 | YES | 1 | 1 | 1 | 0 | 0 | 1 | 0 | 0 | 1 | 1 | 6 | M |
| Brooke1997 | YES | 1 | 0 | 1 | 0 | 0 | 1 | 1 | 0 | 1 | 1 | 6 | M |
| Lord1996 | YES | 1 | 0 | 1 | 0 | 0 | 0 | 0 | 0 | 1 | 1 | 4 | L |
| Kerr1996 | YES | 1 | 0 | 1 | 0 | 0 | 0 | 0 | 0 | 1 | 1 | 4 | L |
| Pruitt1995 | YES | 1 | 0 | 1 | 0 | 0 | 0 | 0 | 0 | 1 | 1 | 4 | L |
| Prince 1995 | YES | 1 | 1 | 1 | 0 | 0 | 0 | 0 | 0 | 1 | 1 | 5 | M |
| Nichols1995 | YES | 1 | 0 | 1 | 0 | 0 | 0 | 0 | 0 | 1 | 1 | 4 | L |
| Bassey1995 | YES | 1 | 0 | 0 | 0 | 0 | 1 | 0 | 0 | 1 | 1 | 4 | L |
| Nelson1994 | YES | 1 | 0 | 0 | 0 | 0 | 0 | 1 | 1 | 1 | 1 | 5 | M |
| Hatori1993 | YES | 1 | 0 | 1 | 0 | 0 | 1 | 1 | 0 | 1 | 1 | 6 | M |
| Martin1993 | YES | 1 | 0 | 1 | 0 | 0 | 0 | 0 | 0 | 1 | 1 | 4 | L |
| Lau1992 | YES | 1 | 0 | 1 | 0 | 0 | 0 | 0 | 0 | 1 | 1 | 4 | L |
| Grove1992 | YES | 1 | 0 | 1 | 0 | 0 | 0 | 1 | 0 | 1 | 1 | 5 | M |
| Sinaki1989 | YES | 1 | 0 | 1 | 0 | 0 | 0 | 1 | 0 | 1 | 1 | 5 | M |

The scale contains 11 items. Item one reflects external validity and is not included in the total PEDro score. The other ten items evaluate the internal validity of a clinical trial.^1^ One point was given for each criterion that was satisfied. Therefore, a score of 0-10 was allocated to each study (≥ 7 = high, 5–6 = moderate, and < 5 = low).^2^

### Appendix 6: Side-split analyses from network meta-analysis of studies examining the efficacy of exercise intervention in postmenopausal women.

|  |  | Beta coefficients ( standard error) | | |  |
| --- | --- | --- | --- | --- | --- |
| Comparison |  | Direct comparison | Indirect comparison | Overall | P-value |
| BMD at LS |  |  |  |  |  |
| control | RT | 0.01(0.00,0.02) | -0.01(-0.04,0.02) | 0.01(0.00,0.02) | 0.2 |
| control | ME | 0.01(0.01,0.02) | 0.02(0.01,0.03) | 0.01(0.01,0.02) | 0.25 |
| control | WBV | 0.01(0.00,0.02) | 0.01(-0.00,0.03 | 0.01(0.00,0.02) | 0.85 |
| control | MIE | 0.01(-0.00,0.02) | -0.01(-0.04,0.02) | 0.01(-0.010.02) | 0.31 |
| control | LIE | 0.01(-0.00,0.02) | 0.01(-0.02,0.04) | 0.01(0.02,0.02) | 0.81 |
| RT | FE | 0.01(-0.01,0.04) | -0.00(-0.02,0.01) | 0.00(-0.01,0.02) | 0.26 |
| RT | WBV | -0.01(-0.06,0.03) | -0.00(-0.01,0.01) | -0.00(-0.01,0.01) | 0.66 |
| RT | MIE | -0.00(-0.02,0.02) | -0.01(-0.03,0.01) | -0.00(-0.02,0.01) | 0.5 |
| RT | LIE | 0.02(-0.05,0.1) | -0.00(-0.01,0.01) | 0.00(-0.01,0.01) | 0.6 |
| RT | MBE | -0.01(-0.03,0.00) | 0.00(-0.01,0.02) | -0.00(-0.01,0.01) | 0.07 |
| ME | WE | -0.02(-0.03,-0.01) | -0.01(-0.03,0.00) | -0.02(-0.02,-0.01) | 0.5 |
| ME | FE | -0.01(-0.02,0.01) | 0.01(-0.02,0.04) | -0.00(-0.02,0.01) | 0.25 |
| ME | WBV | -0.00(-0.04,0.03) | -0.00(-0.01,0.00) | -0.00(-0.01,0.00) | 0.93 |
| ME | MIE | 0.01(-0.05,0.08) | -0.01(-0.02,0.00) | -0.01(-0.02,0.01) | 0.66 |
| WE | WBV | 0.01(-0.00,0.02) | 0.01(-0.00,0.02) | 0.01(0.00,0.02) | 0.92 |
| WE | MIE | -0.02(-0.05,0.02) | 0.01(-0.00,0.0) | 0.01(0.00,0.02) | 0.18 |
| WBV | LIE | 0.00(-0.03,0.03) | 0.00(-0.01,0.01) | 0.00(-0.01,0.01) | 0.97 |
| WBV | MBE | 0.00(-0.02,0.02) | -0.00(-0.02,0.01) | -0.00(-0.01,0.01) | 0.62 |
| HIE | LIE | -0.01(-0.05,0.03) | -0.05(-0.09,0.00) | -0.02(-0.06,0.01) | 0.19 |
| BMD at FN |  |  |  |  |  |
| control | RT | 0.01(-0.00,0.02) | 0.02(-0.02,0.05) | 0.01(-0.00,0.02) | 0.56 |
| control | ME | 0.03(0.02,0.04) | 0.01(-0.02,0.03) | 0.03(0.01,0.04) | 0.13 |
| control | FE | 0.01(-0.03,0.04) | 0.01(-0.02,0.04) | 0.01(-0.01,0.03) | 0.8 |
| control | WBV | 0.01(-0.01,0.03) | 0.02(-0.00,0.05) | 0.02(0.00,0.03) | 0.43 |
| control | MIE | -0.01(-0.03,0.01) | 0.02(-0.02,0.07) | -0.00(-0.02,0.01) | 0.17 |
| control | LIE | 0.01(-0.00,0.03) | -0.00(-0.04,0.03) | 0.01(-0.01,0.02) | 0.39 |
| RT | FE | -0.01(-0.03,0.02) | 0.03(-0.01,0.07) | 0.00(-0.02,0.02) | 0.16 |
| RT | MIE | 0.01(-0.03,0.04) | -0.02(-0.04,0.01) | -0.01(-0.03,0.01) | 0.26 |
| ME | WE | -0.01(-0.04,0.01) | -0.02(-0.05,0.00) | -0.02(-0.04,0.00) | 0.53 |
| ME | FE | 0.00(-0.03,0.04) | -0.03(-0.06,-0.00) | -0.02(-0.04,0.01) | 0.14 |
| WE | WBV | 0.00(-0.02,0.03) | 0.01(-0.02,0.04) | 0.01(-0.01,0.03) | 0.64 |
| WE | MIE | 0.01(-0.03,0.04) | -0.02(-0.05,0.00) | -0.01(-0.03,0.01) | 0.19 |
| WBV | LIE | -0.04(-0.08,0.00) | 0.00(-0.02,0.02) | -0.01(-0.03,0.01) | 0.08 |
| WBV | MBE | -0.00(-0.05,0.04) | 0.00(-0.02,0.03) | 0.00(-0.02,0.02) | 0.86 |
| BMD at TH |  |  |  |  |  |
| control | FE | 0.01(-0.01,0.02) | 0.02(0.00,0.03) | 0.01(0.00,0.02) | 0.27 |
| control | RT | 0.01(0.00,0.02) | 0.00(-0.02,0.03) | 0.01(0.00,0.02) | 0.6 |
| Control | ME | 0.01(0.00,0.02) | 0.00(-0.02,0.02) | 0.01(0.00,0.01) | 0.58 |
| FE | RT | -0.01(-0.02,0.01) | 0.00(-0.01,0.02) | -0.00(-0.01,0.01) | 0.25 |
| FE | ME | -0.01(-0.02,0.01) | -0.00(-0.02,0.01) | -0.01(-0.02,0.01) | 0.65 |
| WBV | MBE | 0.01(-0.01,0.03) | 0.01(-0.00,0.03) | 0.01(-0.00,0.02) | 0.75 |
| MBE | RT | 0.00(-0.02,0.02) | 0.00(-0.01,0.01) | 0.00(-0.01,0.01) | 0.96 |
| RT | ME | -0.01(-0.03,0.00) | -0.00(-0.01,0.01) | -0.00(-0.01,0.01) | 0.16 |

### Appendix 7: Network graph of included studies for bone mineral density

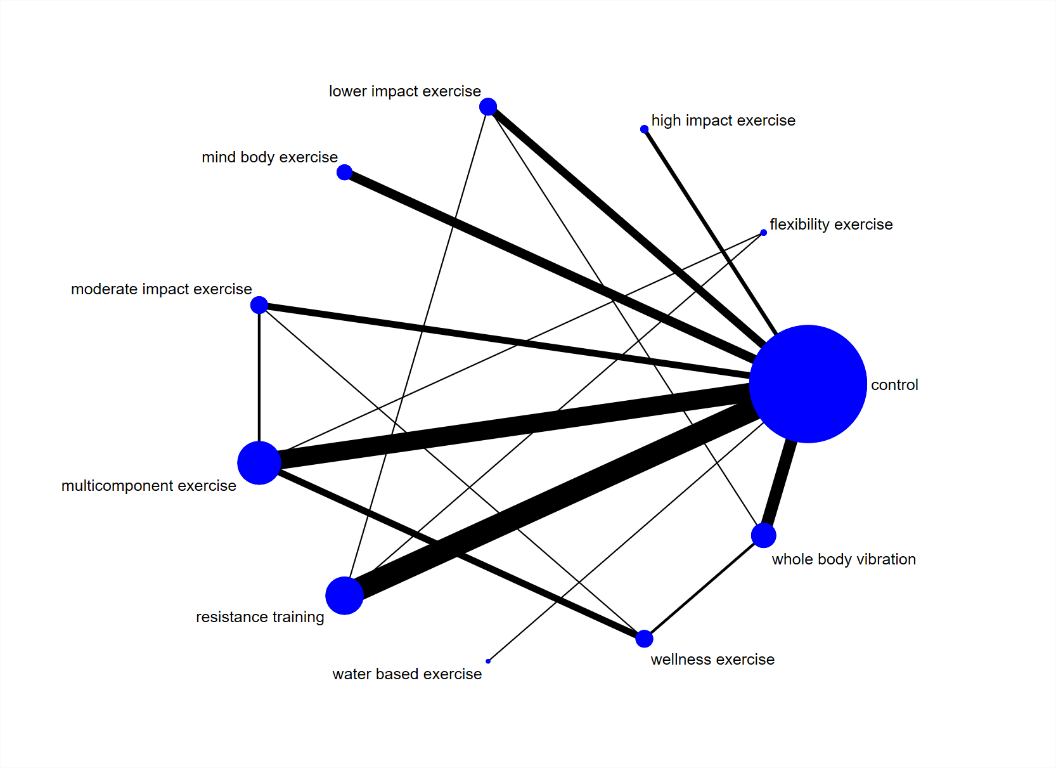
A.

B.

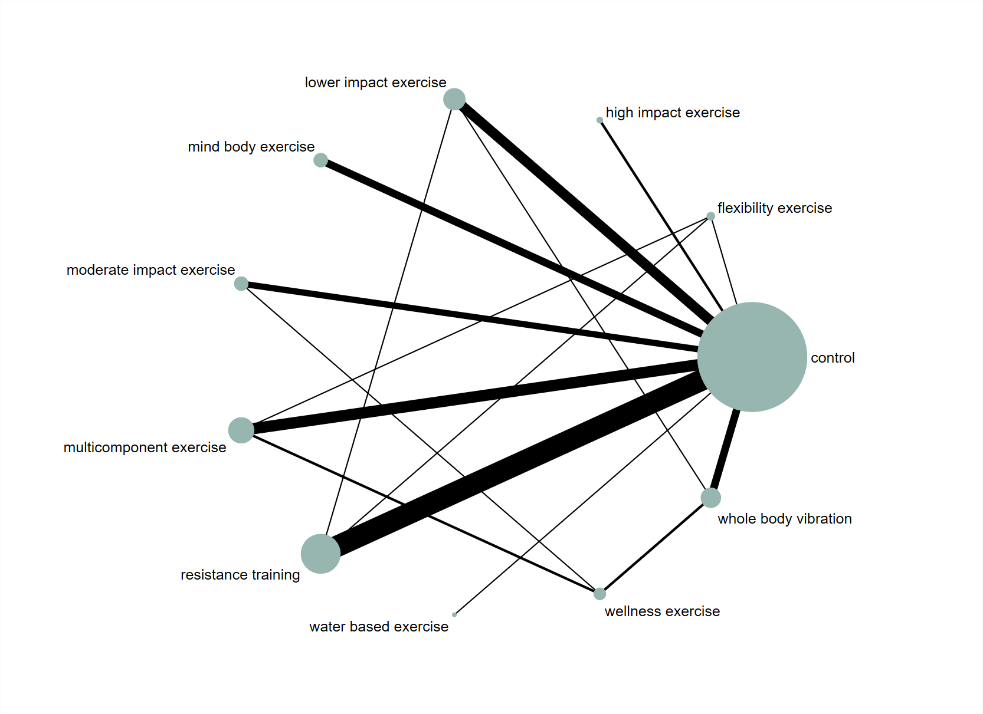

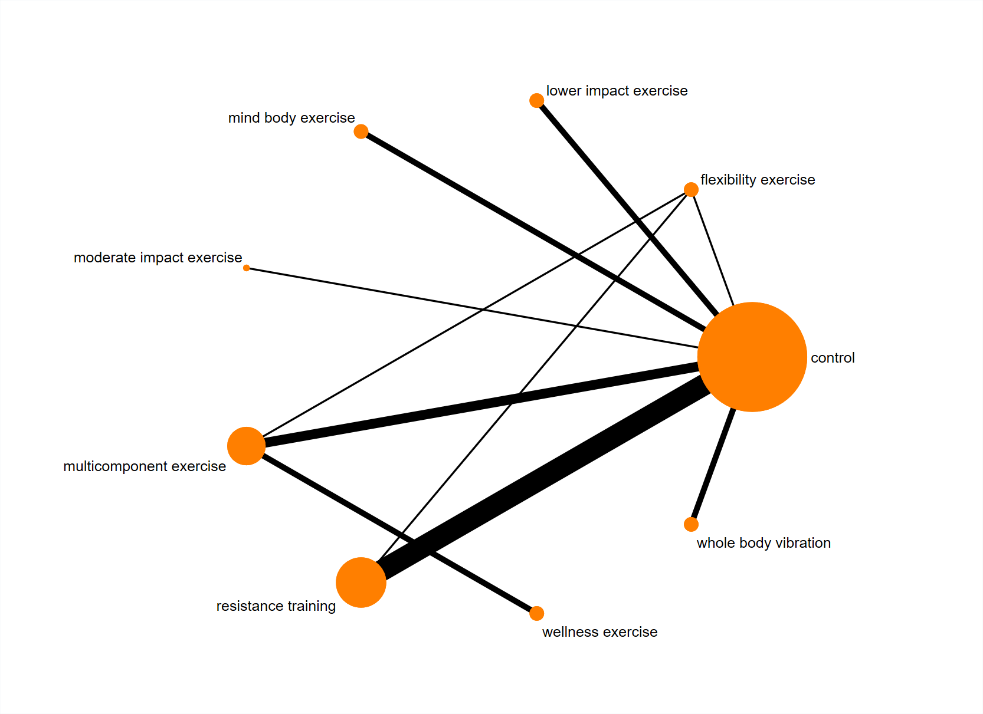

C.

Network graph of included studies for bone mineral density. (A) lumbar spine evidence; (B) femoral neck evidence; (C) total hip evidence.

For each outcome, a network diagram was developed in which nodes represented different types of exercise intervention while lines connecting the nodes represented the direct head-to-head comparisons between interventions. In network plots, the size of nodes and the thickness of the edge lines are proportionate to the number of participants and the number of trials, respectively.

RT: resistance training; WBV: whole body vibration; LIE: lower impact exercise; MIE: moderate impact exercise; HIE: higher impact exercise; CMJ: countermovement jump; MBE: mind based exercise; FE: flexibility exercise; WBE: water based exercise; WE: wellness exercise

### Appendix 8: Cumulative ranking plots for BMD at LS

A.

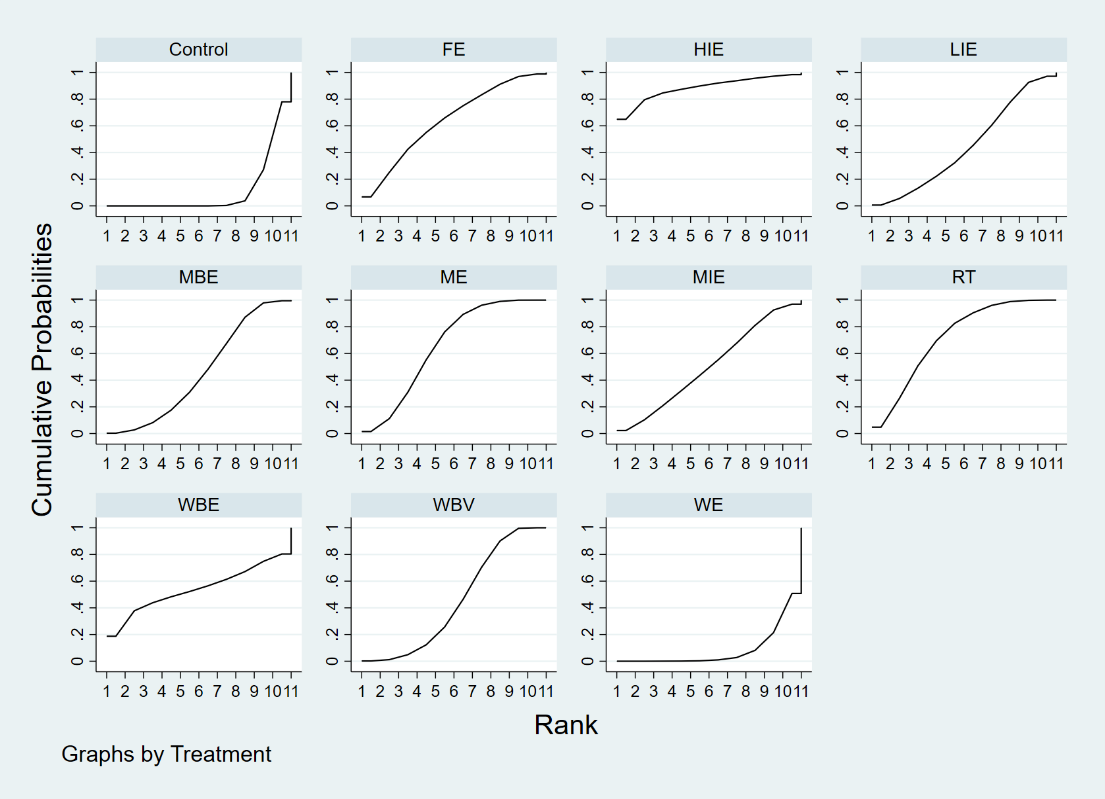

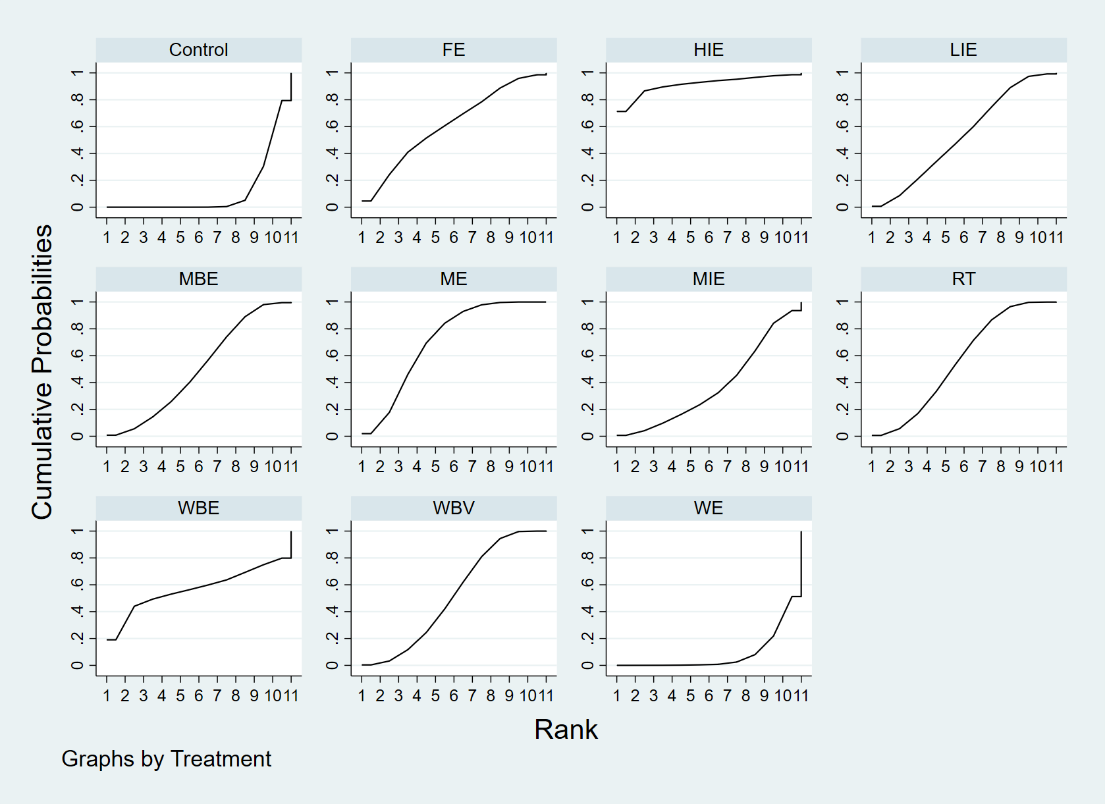

Cumulative ranking plots to show comparative effectiveness of treatments from a BMD outcome at LS network meta-analysis, for each of (A) all trials included, (B) low quality studies excluded.

B.

### Appendix 9: Cumulative ranking plots for BMD at FN

A.

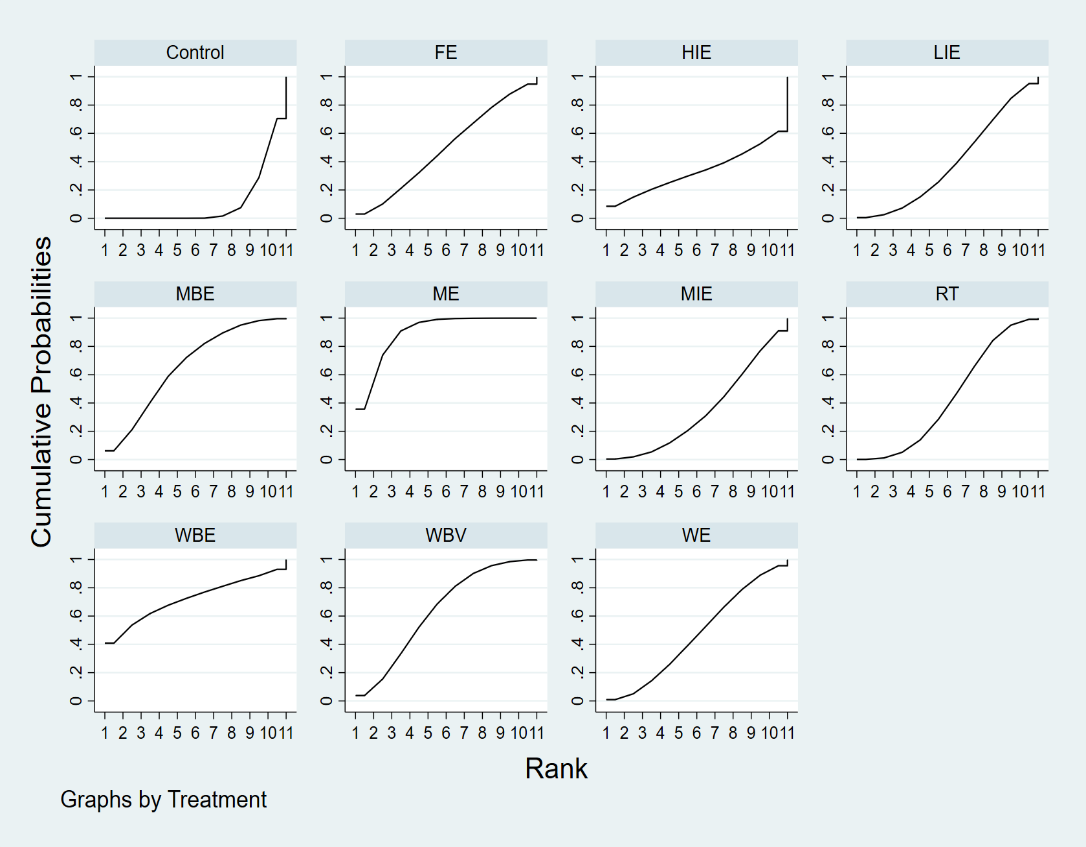

B.

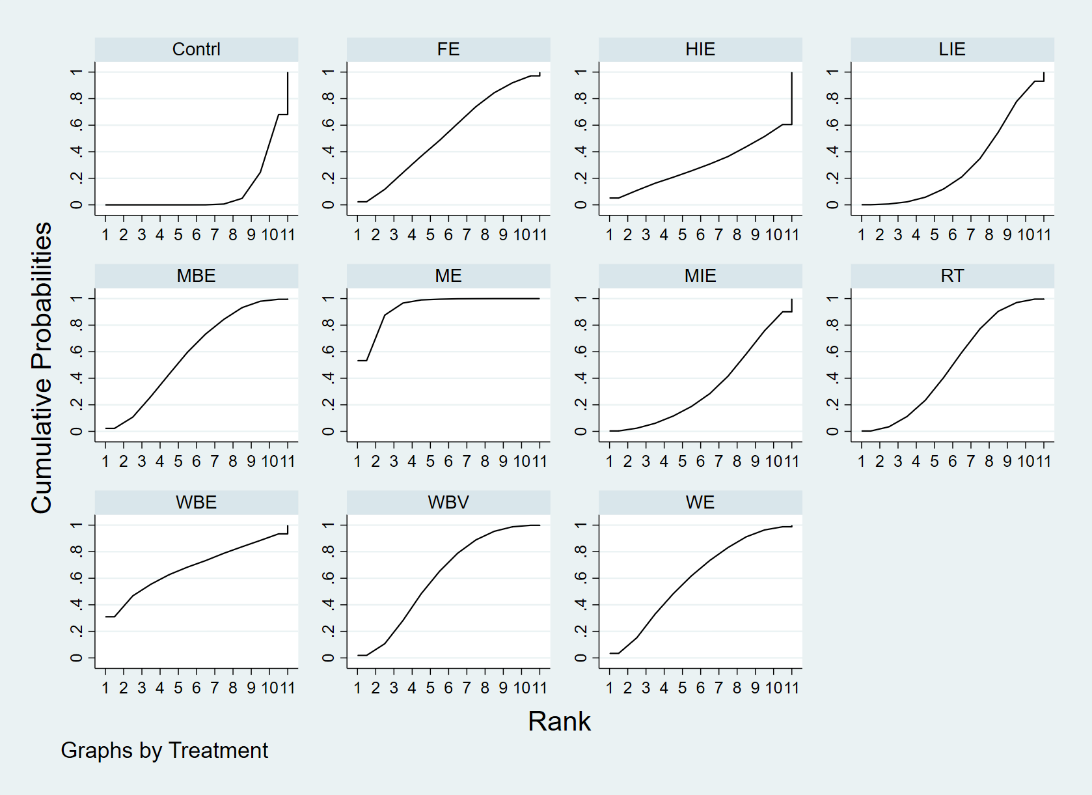

Cumulative ranking plots to show comparative effectiveness of treatments from a BMD outcome at FN network meta-analysis, for each of (A) all trials included, (B) low quality studies excluded.

### Appendix 10: Cumulative ranking plots for BMD at TH

A.

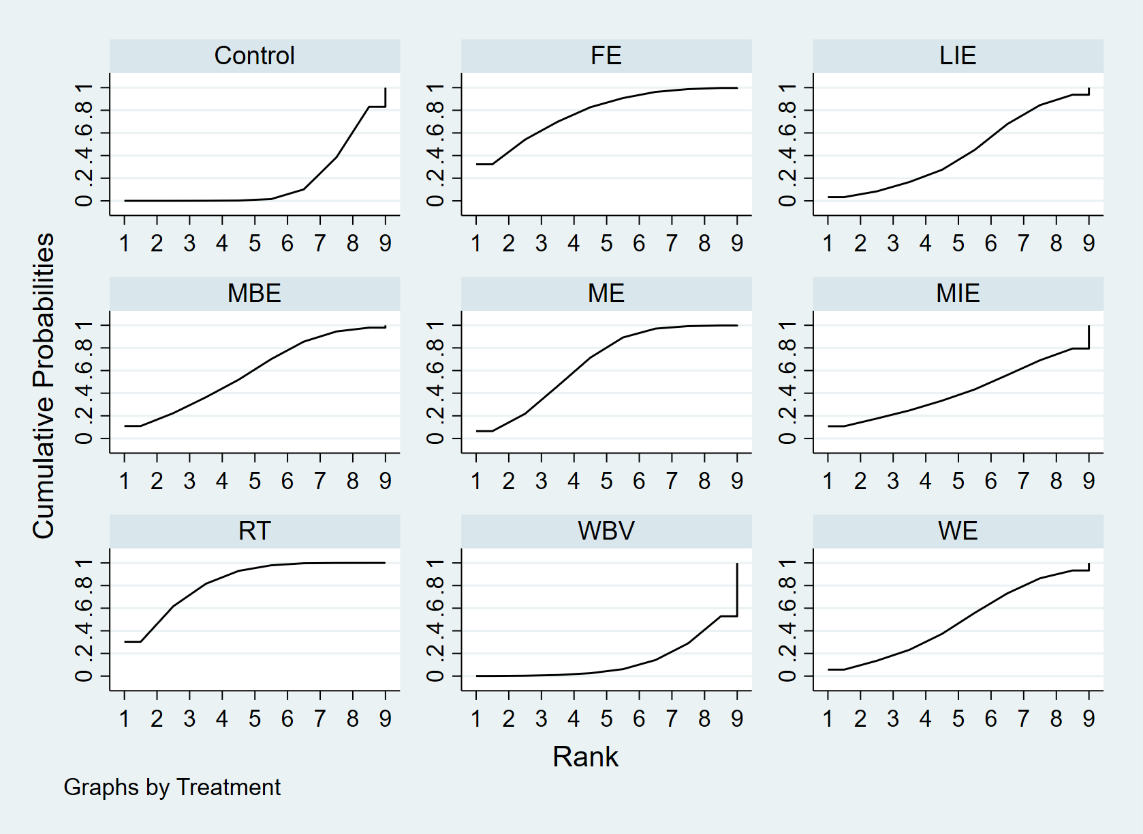

B.

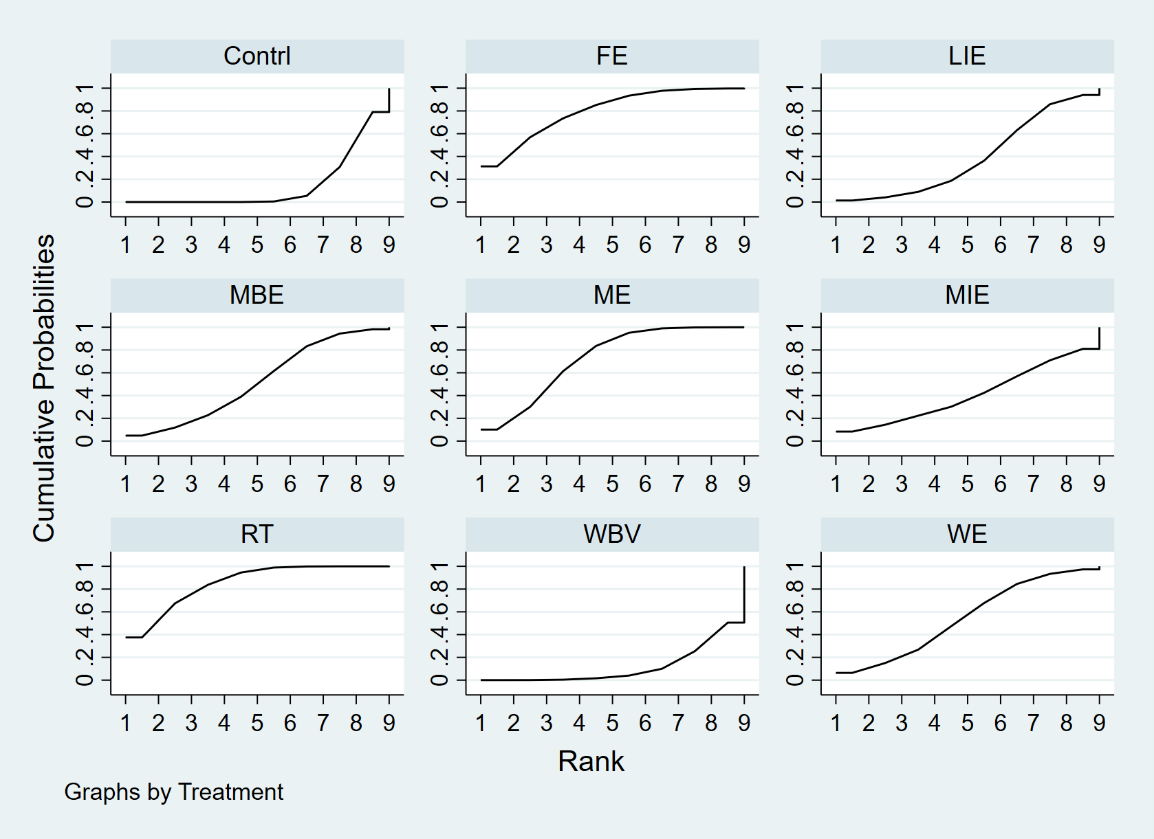

Cumulative ranking plots to show comparative effectiveness of treatments from a BMD outcome at TH network meta-analysis, for each of (A) all trials included, (B) low quality studies excluded.

### Appendix 11: Relative effect estimates for the contrasts between the different interventions and control arms on BMD at TH

| Control | -0.01  (-0.02, 0.00) | **-0.01**  **(-0.02, -0.00)** | 0.00  (-0.01, 0.01) | -0.00  (-0.02, 0.01) | -0.00  (-0.01, 0.00) | -0.01  (-0.02, 0.00) | **-0.01**  **(-0.02, -0.01)** | **-0.01**  **(-0.02, -0.00)** |
| --- | --- | --- | --- | --- | --- | --- | --- | --- |
| -0.00  (-0.02,0.01) | WE | -0.01  (-0.02, 0.01) | 0.01  (-0.01, 0.02) | 0.00  (-0.02, 0.02) | 0.00  (-0.01, 0.02) | -0.00  (-0.02, 0.01) | -0.01  (-0.02, 0.01) | -0.00  (-0.01, 0.01) |
| **-0.01**  **(-0.02,-0.00)** | -0.01  (-0.02,0.01) | FE | **0.02**  **(0.00, 0.03)** | 0.01  (-0.01, 0.03) | 0.01  (-0.00, 0.02) | 0.01  (-0.01, 0.02) | 0.00  (-0.01, 0.01) | 0.00  (-0.01, 0.01) |
| 0.00  (-0.01,0.01) | 0.01  (-0.01,0.02) | **0.01**  **(0.00,0.03)** | WBV | -0.01  (-0.02, 0.01) | -0.01  (-0.02, 0.01) | -0.01  (-0.02, 0.00) | **-0.01**  **(-0.02, -0.00)** | **-0.01**  **(-0.02, -0.00)** |
| -0.00  (-0.02,0.01) | 0.00  (-0.02,0.02) | 0.01  (-0.01,0.03) | -0.01  (-0.02,0.01) | MIE | -0.00  (-0.02, 0.02) | -0.00  (-0.02, 0.01) | -0.01  (-0.02, 0.01) | -0.01  (-0.02, 0.01) |
| -0.00  (-0.01,0.00) | 0.00  (-0.02,0.02) | 0.01  (-0.01,0.02) | -0.01  (-0.02,0.01) | -0.00  (-0.02,0.02) | LIE | -0.00  (-0.02, 0.01) | -0.01  (-0.02, 0.00) | -0.01  (-0.02, 0.01) |
| -0.01  (-0.02,0.00) | -0.00  (-0.02,0.01) | 0.01  (-0.01,0.02) | -0.01  (-0.02,0.00) | -0.00  (-0.02,0.01) | -0.00  (-0.02,0.01) | MBE | -0.00  (-0.01, 0.01) | -0.00  (-0.01, 0.01) |
| **-0.01**  **(-0.02,-0.00)** | -0.01  (-0.02,0.01) | 0.00  (-0.01,0.01) | **-0.01**  **(-0.02,-0.00)** | -0.01  (-0.02,0.01) | -0.01  (-0.02,0.01) | -0.00  (-0.01,0.01) | RT | 0.00  (-0.01, 0.01) |
| **-0.01**  **(-0.01,-0.00)** | -0.00  (-0.01,0.01) | 0.01  (-0.01,0.02) | -0.01  (-0.02,0.00) | -0.00  (-0.02,0.01) | -0.00  (-0.01,0.01) | -0.00  (-0.01,0.01) | 0.01  (-0.01,0.01) | ME |
| Summary estimates from the network meta-analysis are shown in lower left triangle, and summary estimates after excluding low quality studies in upper right triangle. Each cell shows a mean difference (MD), with a 95 % CI in parentheses . For any cell, a negative MD favours the lower-right intervention, and a positive SMD favours the upper-left intervention. Significant results in bold text. | | | | | | | | |

### Appendix 12.1: Relative effect estimates for the contrasts between the different interventions and control arms on BMD at LS in participants with age no less than 60

| Control |  |  |  |  |  |  |  |  |
| --- | --- | --- | --- | --- | --- | --- | --- | --- |
| -0.00  (-0.01, 0.01) | WE |  |  |  |  |  |  |  |
| -0.02  (-0.04, 0.00) | -0.02  (-0.04, 0.01) | FE |  |  |  |  |  |  |
| **-0.01**  **(-0.02, -0.00)** | **-0.01**  **(-0.02, -0.00)** | 0.01  (-0.02, 0.03) | WBV |  |  |  |  |  |
| -0.01  (-0.02, 0.01) | -0.00  (-0.02, 0.01) | 0.02  (-0.01, 0.04) | 0.01  (-0.01, 0.03) | MIE |  |  |  |  |
| **-0.01**  **(-0.02, -0.00)** | -0.01  (-0.02, 0.01) | 0.01  (-0.01, 0.04) | 0.00  (-0.01, 0.02) | -0.00  (-0.02, 0.02) | LIE |  |  |  |
| 0.00  (-0.01, 0.02) | 0.00  (-0.02, 0.03) | 0.02  (-0.00, 0.05) | 0.02  (-0.00, 0.04) | 0.01  (-0.01, 0.03) | 0.01  (-0.01, 0.03) | MBE |  |  |
| **-0.01**  **(-0.01, -0.00)** | -0.01  (-0.02, 0.01) | 0.01  (-0.01, 0.03) | 0.01  (-0.01, 0.02) | -0.00  (-0.02, 0.02) | 0.00  (-0.01, 0.01) | -0.01  (-0.03, 0.01) | RT |  |
| **-0.02**  **(-0.03, -0.01)** | **-0.01**  **(-0.02, -0.00)** | 0.01  (-0.02, 0.03) | -0.00  (-0.01, 0.01) | -0.01  (-0.03, 0.01) | -0.01  (-0.02, 0.00) | -0.02  (-0.04, 0.00) | -0.01  (-0.02, 0.00) | ME |

| Control |  |  |  |  |  |  |  |  |
| --- | --- | --- | --- | --- | --- | --- | --- | --- |
| -0.03  (-0.07,0.01) | WE |  |  |  |  |  |  |  |
| 0.00  (-0.03,0.03) | 0.03  (-0.02,0.08) | FE |  |  |  |  |  |  |
| -0.03  (-0.07,0.00) | -0.01  (-0.04,0.02) | -0.03  (-0.09,0.01) | WBV |  |  |  |  |  |
| -0.00  (-0.02,0.02) | 0.02  (-0.02,0.07) | -0.00  (-0.04,0.04) | 0.03  (-0.01,0.07) | MIE |  |  |  |  |
| -0.01  (-0.03,0.01) | 0.02  (-0.02,0.06) | -0.01  (-0.05,0.03) | 0.03  (-0.01,0.06) | -0.01  (-0.04,0.02) | LIE |  |  |  |
| -0.01  (-0.06,0.03) | 0.01  (-0.04,0.07) | -0.01  (-0.07,0.04) | 0.02  (-0.04,0.08) | -0.01  (-0.06,0.04) | -0.00  (-0.05,0.05) | MBE |  |  |
| -0.01  (-0.02,0.01) | 0.02  (-0.02,0.06) | -0.01  (-0.04,0.02) | -0.03  (-0.01,0.07) | -0.01  (-0.04,0.02) | 0.00  (-0.02,0.03) | -0.01  (-0.04,0.05) | RT |  |
| **-0.03**  **(-0.06,-0.01)** | -0.01  (-0.04,0.03) | -0.03  (-0.07,0.01) | 0.00  (-0.04,0.04) | **-0.03**  **(-0.07,-0.00)** | -0.03  (-0.05,0.00) | -0.02  (-0.07,0.03) | **-0.03**  **(-0.06,-0.00)** | ME |

### Appendix 12.2: Relative effect estimates for the contrasts between the different interventions and control arms on BMD at FN in participants with age no less than 60

### Appendix 12.3: Relative effect estimates for the contrasts between the different interventions and control arms on BMD at TH in participants with age no less than 60

| Control |  |  |  |  |  |  |  |  |
| --- | --- | --- | --- | --- | --- | --- | --- | --- |
| -0.01  (-0.04,0.02) | WE |  |  |  |  |  |  |  |
| -0.01  (-0.02,0.01) | -0.00  (-0.03,0.04) | FE |  |  |  |  |  |  |
| -0.00  (-0.03,0.02) | -0.01  (-0.03,0.05) | -0.00  (-0.02,0.03) | HIE |  |  |  |  |  |
| -0.01  (-0.02,0.01) | -0.00  (-0.03,0.04) | -0.00  (-0.03,0.02) | -0.00  (-0.03,0.03) | MIE |  |  |  |  |
| -0.02  (-0.04,0.01) | -0.01  (-0.05,0.03) | -0.01  (-0.04,0.02) | -0.01  (-0.05,0.02) | -0.01  (-0.04,0.02) | LIE |  |  |  |
| -0.02  (-0.04,0.01) | -0.01  (-0.05,0.03) | -0.01  (-0.04,0.02) | -0.01  (-0.05,0.02) | -0.01  (-0.04,0.02) | -0.00  (-0.04,0.04) | MBE |  |  |
| -0.01  (-0.02,0.00) | -0.00  (-0.03,0.03) | -0.00  (-0.02,0.01) | -0.01  (-0.03,0.02) | -0.00  (-0.02,0.02) | 0.01  (-0.02,0.04) | 0.01  (-0.02,0.04) | WBE |  |
| -0.01  (-0.03,0.01) | -0.00  (-0.02,0.02) | -0.00  (-0.04,0.03) | -0.01  (-0.04,0.03) | -0.00  (-0.03,0.03) | -0.01  (-0.03,0.04) | 0.01  (-0.03,0.04) | -0.00  (-0.03,0.03) | RT |

### Appendix 13: Cumulative ranking plots to show comparative effectiveness of treatments from a BMD outcome at (A) LS, (B) FN, and (C) FN network meta-analysis in participants with age no less than 60

A.

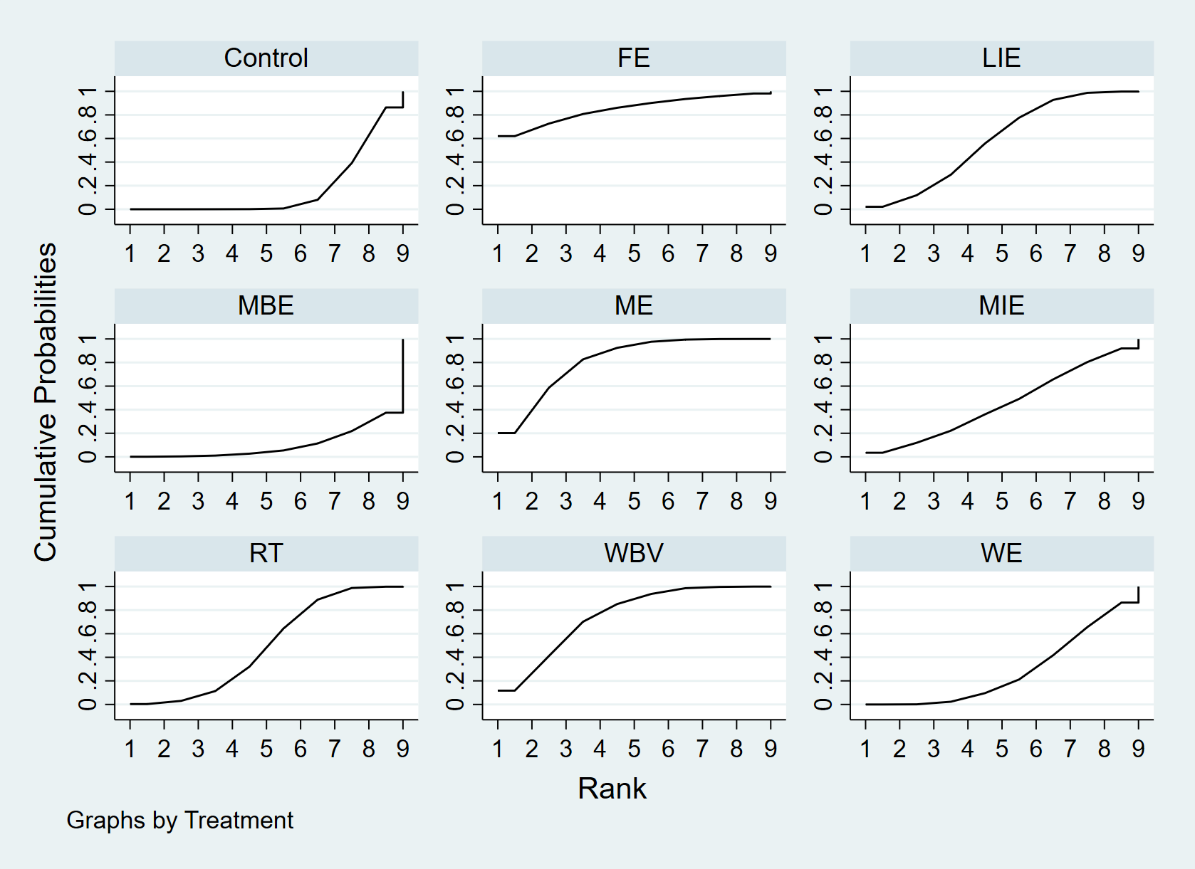

B.

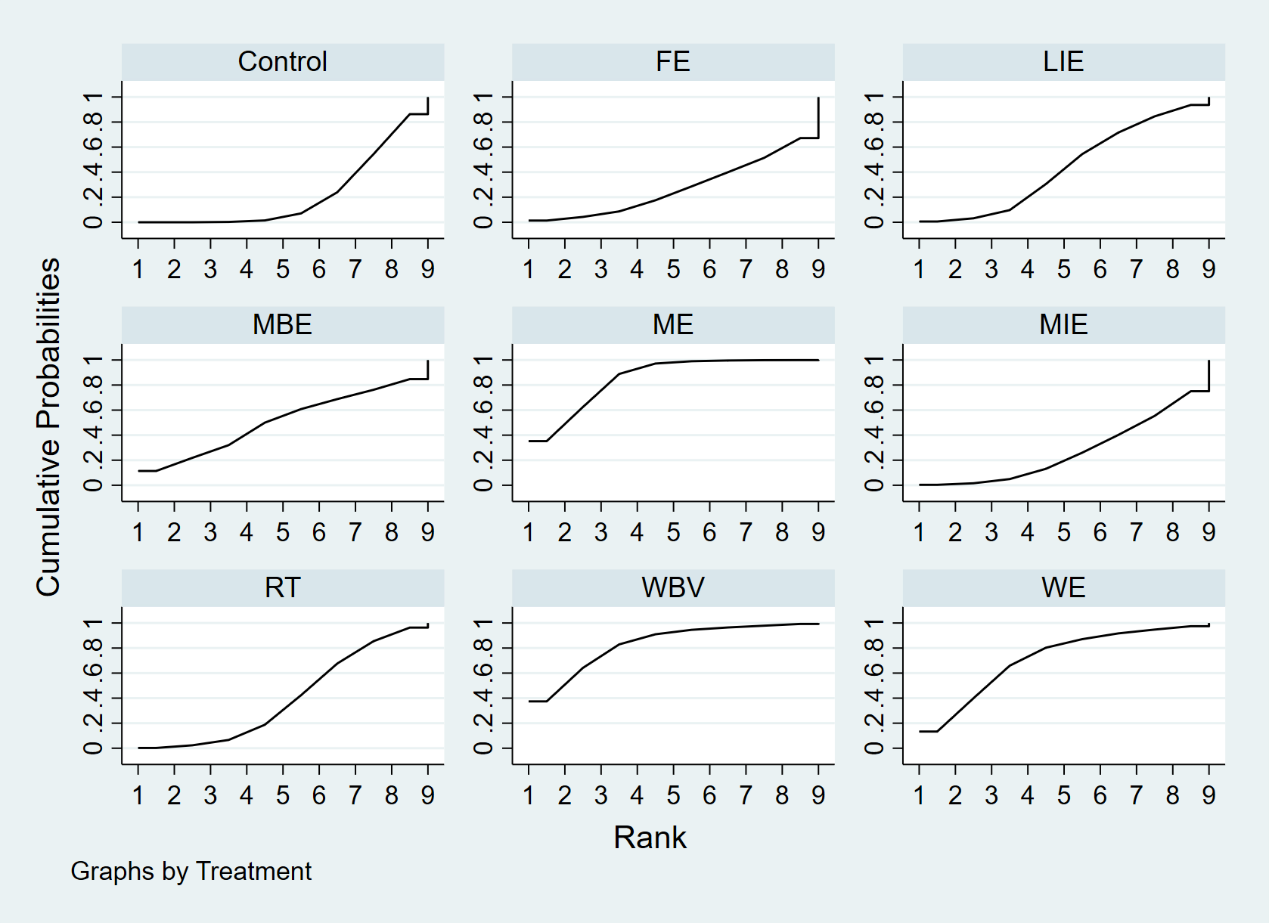

C.

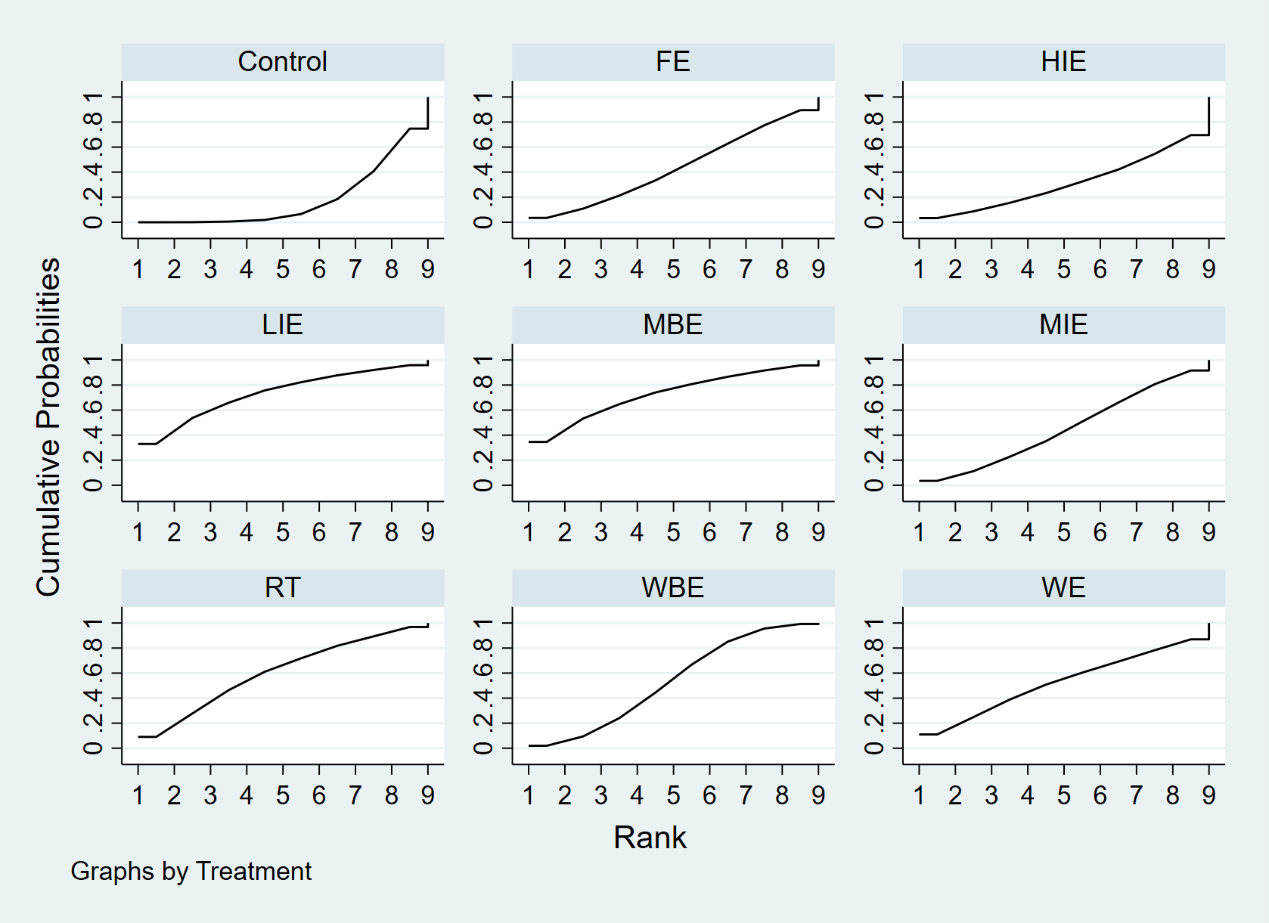

### Appendix 14.1: Relative effect estimates for the contrasts between the different interventions and control arms on BMD at LS in participants younger than 60 years old

| Control | **-0.01**  **(-0.03, -0.00)** | **-0.01**  **(-0.03, -0.00)** | 0.00  (-0.02, 0.02) | -0.01  (-0.03, 0.02) | -0.01  (-0.02, 0.00) | **-0.04**  **(-0.08, -0.00)** | -0.01  (-0.03, 0.01) | -0.02  (-0.05, 0.01) | **-0.01**  **(-0.03, -0.00)** | -0.01  (-0.06, 0.03) |
| --- | --- | --- | --- | --- | --- | --- | --- | --- | --- | --- |
|  | RT | 0.00  (-0.02, 0.02) | 0.02  (-0.01, 0.04) | 0.01  (-0.02, 0.03) | 0.01  (-0.01, 0.02) | -0.03  (-0.07, 0.02) | 0.01  (-0.02, 0.03) | -0.01  (-0.04, 0.03) | 0.00  (-0.02, 0.02) | 0.00  (-0.04, 0.05) |
|  |  | ME | **0.02**  **(0.00, 0.03)** | 0.01  (-0.02, 0.03) | 0.00  (-0.01, 0.02) | -0.03  (-0.07, 0.01) | 0.01  (-0.02, 0.03) | -0.01  (-0.04, 0.03) | 0.00  (-0.02, 0.02) | -0.00  (-0.04, 0.05) |
|  |  |  | WE | -0.01  (-0.04, 0.02) | -0.01  (-0.03, 0.01) | -0.04  (-0.09, 0.00) | -0.01  (-0.03, 0.02) | -0.02  (-0.06, 0.01) | -0.02  (-0.04, 0.01) | -0.02  (-0.06, 0.03) |
|  |  |  |  | FE | -0.00  (-0.03, 0.02) | -0.03  (-0.08, 0.01) | -0.00  (-0.03, 0.03) | -0.01  (-0.05, 0.03) | -0.01  (-0.03, 0.02) | -0.01  (-0.06, 0.04) |
|  |  |  |  |  | WBV | -0.03  (-0.08, 0.01) | 0.00  (-0.02, 0.03) | -0.01  (-0.05, 0.02) | -0.00  (-0.02, 0.01) | -0.00  (-0.05, 0.04) |
|  |  |  |  |  |  | HIE | 0.03  (-0.01, 0.08) | 0.02  (-0.02, 0.06) | 0.03  (-0.01, 0.07) | 0.03  (-0.03, 0.09) |
|  |  |  |  |  |  |  | MIE | -0.01  (-0.05, 0.03) | -0.01  (-0.03, 0.02) | -0.01  (-0.05, 0.04) |
|  |  |  |  |  |  |  |  | LIE | 0.01  (-0.03, 0.04) | 0.01  (-0.05, 0.06) |
|  |  |  |  |  |  |  |  |  | MBE | -0.00  (-0.05, 0.05) |
|  |  |  |  |  |  |  |  |  |  | WBE |

### Appendix 14.2: Relative effect estimates for the contrasts between the different interventions and control arms on BMD at FN in participants younger than 60 years old

| Control | -0.01  (-0.03, 0.00) | **-0.03**  **(-0.05, -0.01)** | -0.02  (-0.05, 0.01) | **-0.03**  **(-0.07, -0.00)** | -0.01  (-0.03, 0.00) | -0.00  (-0.04, 0.03) | -0.02  (-0.05, 0.01) | -0.01  (-0.04, 0.01) | **-0.02**  **(-0.03, -0.00)** | -0.02  (-0.06, 0.01) |
| --- | --- | --- | --- | --- | --- | --- | --- | --- | --- | --- |
|  | RT | -0.02  (-0.04, 0.01) | -0.01  (-0.04, 0.03) | -0.02  (-0.06, 0.02) | 0.00  (-0.02, 0.02) | 0.01  (-0.03, 0.05) | -0.01  (-0.04, 0.02) | -0.00  (-0.03, 0.03) | -0.00  (-0.03, 0.02) | -0.01  (-0.05, 0.03) |
|  |  | ME | 0.01  (-0.01, 0.04) | -0.00  (-0.03, 0.02) | 0.02  (-0.00, 0.04) | 0.03  (-0.01, 0.07) | 0.01  (-0.02, 0.04) | 0.01  (-0.02, 0.05) | 0.01  (-0.01, 0.03) | 0.00  (-0.03, 0.04) |
|  |  |  | WE | -0.02  (-0.05, 0.02) | 0.01  (-0.02, 0.04) | 0.01  (-0.03, 0.06) | -0.00  (-0.03, 0.02) | 0.00  (-0.04, 0.04) | 0.00  (-0.03, 0.03) | -0.01  (-0.05, 0.04) |
|  |  |  |  | FE | 0.02  (-0.01, 0.06) | 0.03  (-0.02, 0.08) | 0.01  (-0.03, 0.05) | 0.02  (-0.02, 0.06) | 0.02  (-0.02, 0.05) | 0.01  (-0.04, 0.06) |
|  |  |  |  |  | WBV | 0.01  (-0.03, 0.05) | -0.01  (-0.04, 0.02) | -0.00  (-0.04, 0.03) | -0.01  (-0.03, 0.01) | -0.01  (-0.05, 0.03) |
|  |  |  |  |  |  | HIE | -0.02  (-0.07, 0.03) | -0.01  (-0.06, 0.03) | -0.01  (-0.06, 0.03) | -0.02  (-0.07, 0.03) |
|  |  |  |  |  |  |  | MIE | 0.01  (-0.03, 0.05) | 0.00  (-0.03, 0.04) | -0.00  (-0.05, 0.04) |
|  |  |  |  |  |  |  |  | LIE | -0.00  (-0.03, 0.03) | -0.01  (-0.05, 0.03) |
|  |  |  |  |  |  |  |  |  | MBE | -0.01  (-0.04, 0.03) |
|  |  |  |  |  |  |  |  |  |  | WBE |

### Appendix 14.3: Relative effect estimates for the contrasts between the different interventions and control arms on BMD at TH in participants younger than 60 years old

| Control | -0.01  (-0.03,0.02) | -0.02  (-0.04,0.00) | 00.0  (-0.01,0.01) | -0.00  (-0.02,0.02) | -0.00  (-0.02,0.01) | -0.02  (-0.04,0.00) | -0.01  (-0.02,0.00) | -0.01  (-0.02,0.00) |
| --- | --- | --- | --- | --- | --- | --- | --- | --- |
|  | WE | -0.01  (-0.04,0.01) | 0.01  (-0.02,0.03) | 0.00  (-0.03,0.03) | 0.00  (-0.02,0.03) | -0.01  (-0.04,0.02) | -0.01  (-0.03,0.02) | -0.00  (-0.02,0.01) |
|  |  | FE | 0.02  (-0.01,0.04) | 0.02  (-0.01,0.05) | 0.01  (-0.01,0.04) | -0.00  (-0.03,0.03) | 0.01  (-0.02,0.03) | 0.01  (-0.01,0.03) |
|  |  |  | WBV | -0.00  (-0.03,0.02) | -0.01  (-0.02,0.01) | -0.02  (-0.04,0.00) | -0.01  (-0.04,0.00) | -0.01  (-0.03,0.00) |
|  |  |  |  | MIE | -0.00  (-0.03,0.02) | -0.02  (-0.05,0.01) | -0.01  (-0.03,0.01) | -0.01  (-0.03,0.01) |
|  |  |  |  |  | LIE | -0.01  (-0.04,0.01) | -0.01  (-0.03,0.01) | -0.01  (-0.02,0.01) |
|  |  |  |  |  |  | MBE | 0.01  (-0.02,0.03) | 0.01  (-0.01,0.03) |
|  |  |  |  |  |  |  | WBE | 0.00  (-0.01,0.02) |
|  |  |  |  |  |  |  |  | RT |

### Appendix 15: Cumulative ranking plots to show comparative effectiveness of treatments from a BMD outcome at (A) LS, (B) FN, and (C) FN network meta-analysis in participants younger than 60 years old

A.

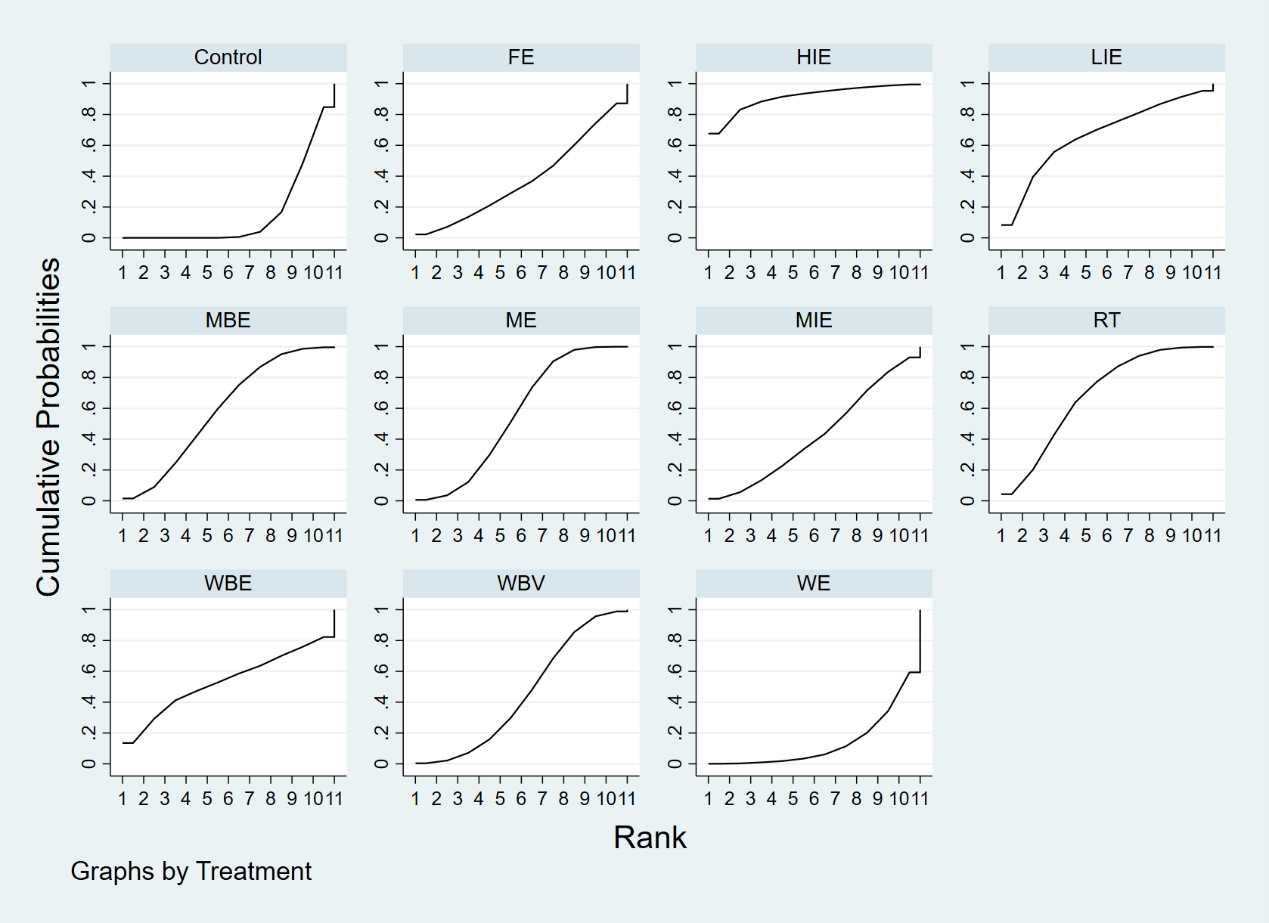

B.

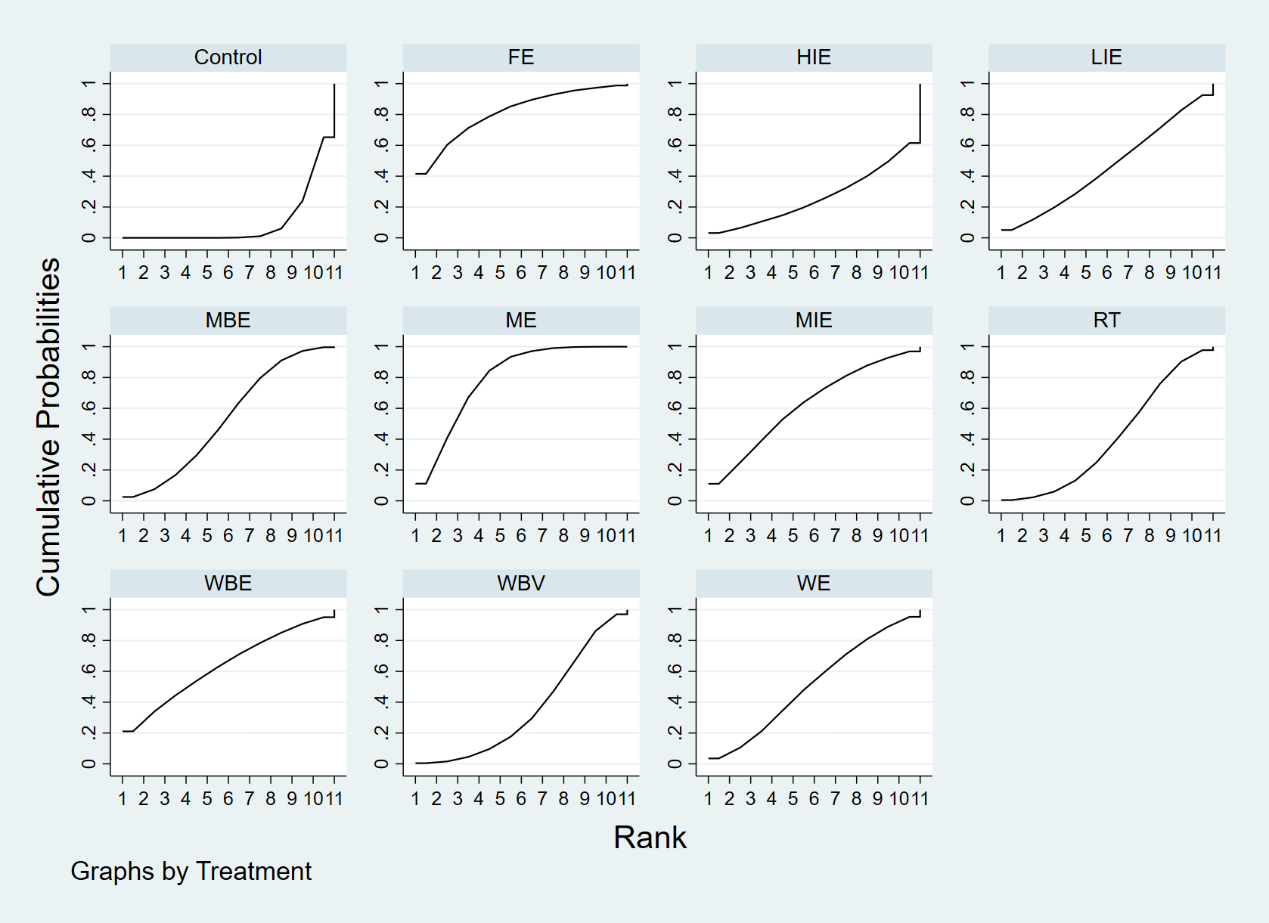

C.

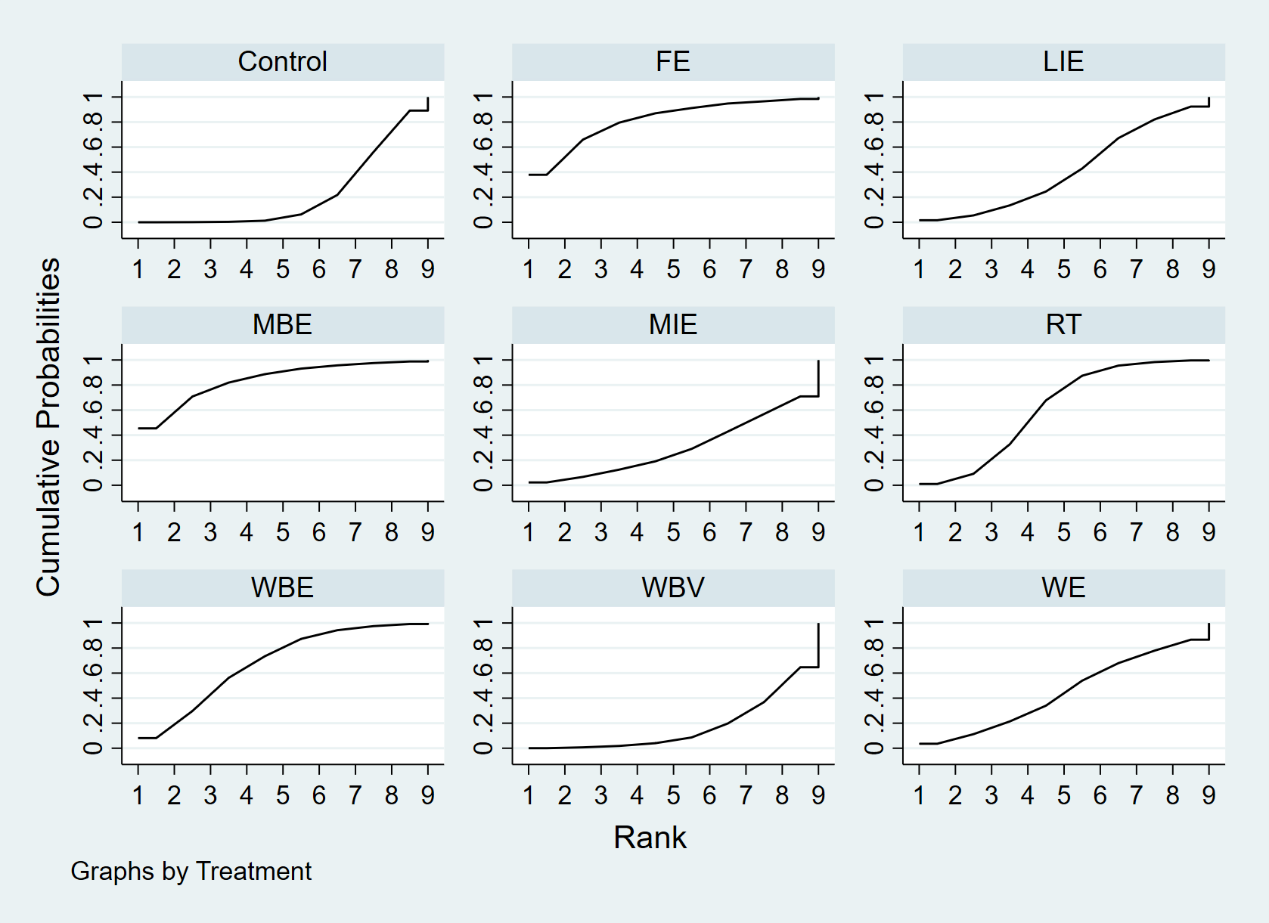

### Appendix 16.1: Relative effect estimates for the contrasts between the different interventions and control arms on BMD at LS when exercise intervention with duration no more than eight months

| Control |  |  |  |  |  |  |  |  |
| --- | --- | --- | --- | --- | --- | --- | --- | --- |
| **-0.02**  **(-0.03, -0.01)** | WBV |  |  |  |  |  |  |  |
| 0.02  (-0.02, 0.08) | 0.04  (-0.00, 0.10) | HIE |  |  |  |  |  |  |
| -0.02  (-0.04, 0.01) | 0.01  (-0.02, 0.03) | -0.04  (-0.10, 0.01) | MIE |  |  |  |  |  |
| -0.02  (-0.04, 0.00) | 0.00  (-0.02, 0.02) | -0.04  (-0.11, 0.00) | -0.00  (-0.04, 0.02) | LIE |  |  |  |  |
| **-0.02**  **(-0.04, -0.00)** | 0.00  (-0.02, 0.02) | -0.04  (-0.11, 0.00) | -0.01  (-0.03, 0.02) | -0.00  (-0.03, 0.03) | MBE |  |  |  |
| -0.01  (-0.05, 0.03) | 0.01  (-0.03, 0.05) | -0.03  (-0.11, 0.02) | 0.01  (-0.04, 0.05) | 0.01  (-0.04, 0.05) | 0.01  (-0.04, 0.05) | WBE |  |  |
| **-0.01**  **(-0.03, -0.00)** | 0.01  (-0.01, 0.02) | -0.03  (-0.10, 0.01) | 0.00  (-0.02, 0.02) | 0.01  (-0.02, 0.03) | 0.01  (-0.02, 0.03) | -0.00  (-0.04, 0.04) | RT |  |
| **-0.02**  **(-0.04, -0.01)** | -0.00  (-0.02, 0.02) | -0.04  (-0.11, 0.00) | -0.01  (-0.03, 0.02) | -0.00  (-0.03, 0.03) | -0.00  (-0.02, 0.02) | -0.01  (-0.05, 0.03) | -0.01  (-0.03, 0.01) | ME |

| Control |  |  |  |  |  |  |  |  |  |
| --- | --- | --- | --- | --- | --- | --- | --- | --- | --- |
| **-0.05**  **(-0.07, -0.03)** | ME |  |  |  |  |  |  |  |  |
| -0.01  (-0.04, 0.02) | **0.04**  **(0.01, 0.08)** | FE |  |  |  |  |  |  |  |
| **-0.03**  **(-0.05, -0.01)** | 0.02  (-0.00, 0.05) | -0.02  (-0.06, 0.02) | WBV |  |  |  |  |  |  |
| -0.01  (-0.05, 0.03) | 0.04  (-0.00, 0.09) | -0.00  (-0.05, 0.05) | 0.02  (-0.03, 0.06) | HIE |  |  |  |  |  |
| -0.01  (-0.04, 0.02) | **0.04**  **(0.01, 0.08)** | -0.00  (-0.04, 0.04) | 0.02  (-0.02, 0.05) | 0.00  (-0.05, 0.05) | MIE |  |  |  |  |
| -0.01  (-0.03, 0.01) | **0.05**  **(0.02, 0.07)** | 0.00  (-0.03, 0.03) | 0.02  (-0.01, 0.05) | 0.01  (-0.04, 0.05) | 0.01  (-0.03, 0.04) | LIE |  |  |  |
| -0.02  (-0.06, 0.02) | 0.03  (-0.01, 0.08) | -0.01  (-0.06, 0.04) | 0.01  (-0.03, 0.05) | -0.00  (-0.06, 0.05) | -0.01  (-0.05, 0.04) | -0.01  (-0.05, 0.03) | MBE |  |  |
| -0.02  (-0.06, 0.01) | 0.03  (-0.01, 0.07) | -0.02  (-0.06, 0.03) | 0.00  (-0.04, 0.05) | -0.01  (-0.07, 0.04) | -0.01  (-0.06, 0.03) | -0.02  (-0.06, 0.03) | -0.01  (-0.06, 0.05) | WBE |  |
| -0.01  (-0.02, 0.01) | **0.05**  **(0.02, 0.07)** | 0.00  (-0.03, 0.03) | 0.02  (-0.00, 0.05) | 0.01  (-0.04, 0.05) | 0.01  (-0.02, 0.03) | 0.00  (-0.02, 0.03) | 0.01  (-0.03, 0.05) | 0.02  (-0.02, 0.06) | RT |

### Appendix 16.2: Relative effect estimates for the contrasts between the different interventions and control arms on BMD at FN when exercise intervention with duration no more than eight months

### Appendix 16.3: Relative effect estimates for the contrasts between the different interventions and control arms on BMD at TH when exercise intervention with duration no more than eight months

| Control |  |  |  |  |  |  |  |
| --- | --- | --- | --- | --- | --- | --- | --- |
| -0.01  (-0.03, 0.02) | FE |  |  |  |  |  |  |
| 0.01  (-0.02, 0.04) | 0.02  (-0.02, 0.06) | WBV |  |  |  |  |  |
| -0.00  (-0.03, 0.02) | 0.01  (-0.03, 0.04) | -0.01  (-0.05, 0.03) | MIE |  |  |  |  |
| -0.00  (-0.02, 0.02) | 0.01  (-0.03, 0.04) | -0.01  (-0.05, 0.02) | 0.00  (-0.03, 0.03) | LIE |  |  |  |
| 0.00  (-0.03, 0.03) | 0.01  (-0.03, 0.05) | -0.01  (-0.04, 0.02) | 0.00  (-0.04, 0.04) | 0.00  (-0.03, 0.04) | MBE |  |  |
| -0.01  (-0.02, 0.01) | -0.00  (-0.03, 0.03) | -0.02  (-0.05, 0.01) | -0.01  (-0.03, 0.02) | -0.01  (-0.03, 0.02) | -0.01  (-0.04, 0.03) | RT |  |
| -0.02  (-0.04, 0.00) | -0.01  (-0.04, 0.02) | -0.03  (-0.06, 0.01) | -0.01  (-0.05, 0.02) | -0.02  (-0.04, 0.01) | -0.02  (-0.05, 0.02) | -0.01  (-0.04, 0.02) | ME |

### Appendix 17: Cumulative ranking plots to show comparative effectiveness of treatments from a BMD outcome at (A) LS, (B) FN, and (C) FN network meta-analysis when exercise intervention with duration no more than eight months

A.

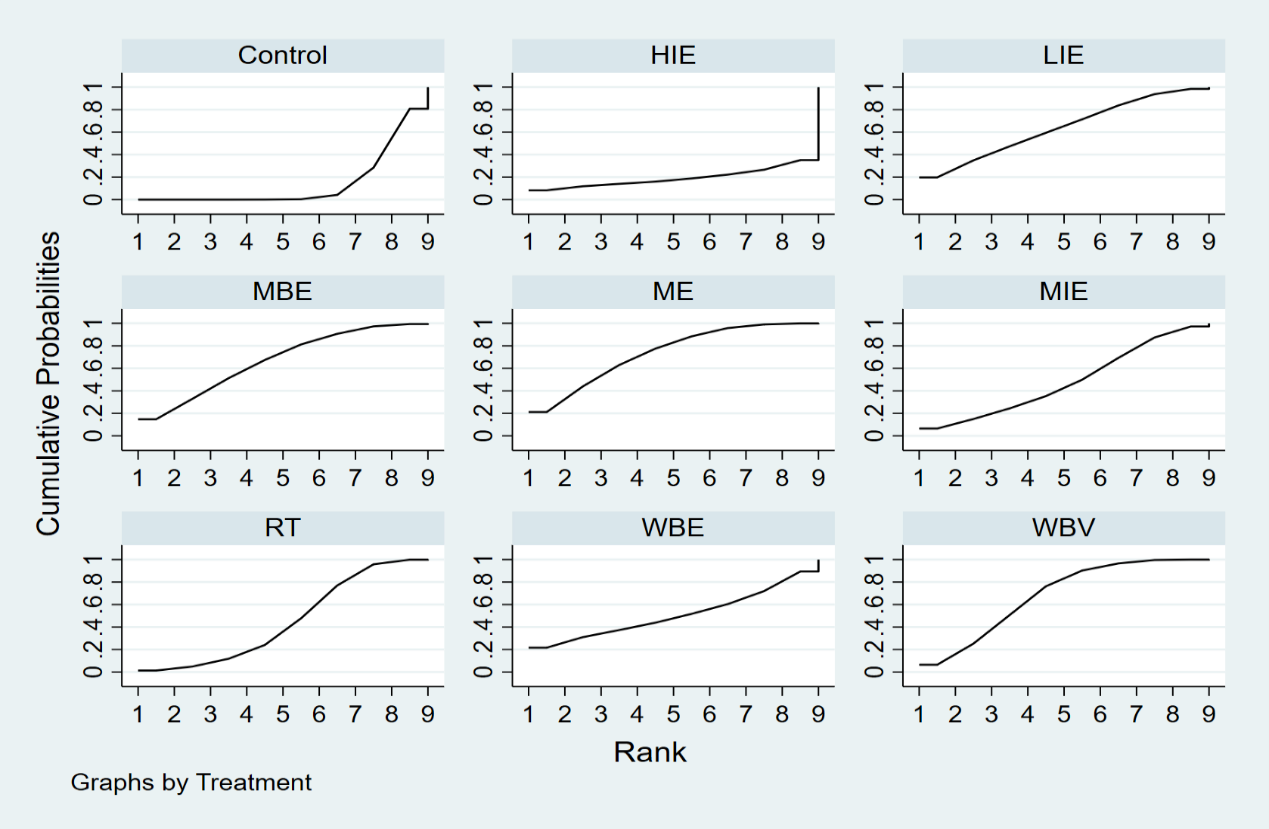

B.
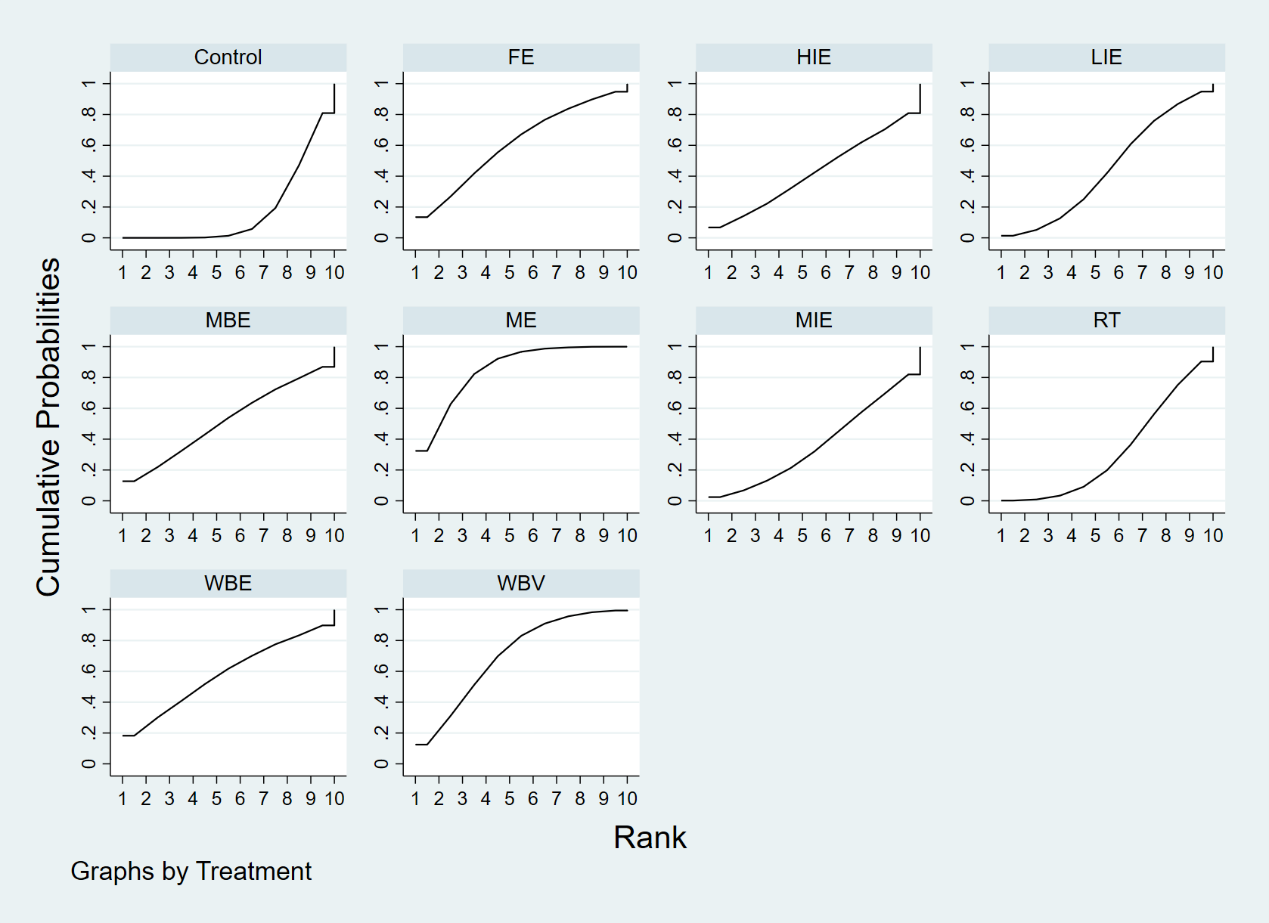

C.

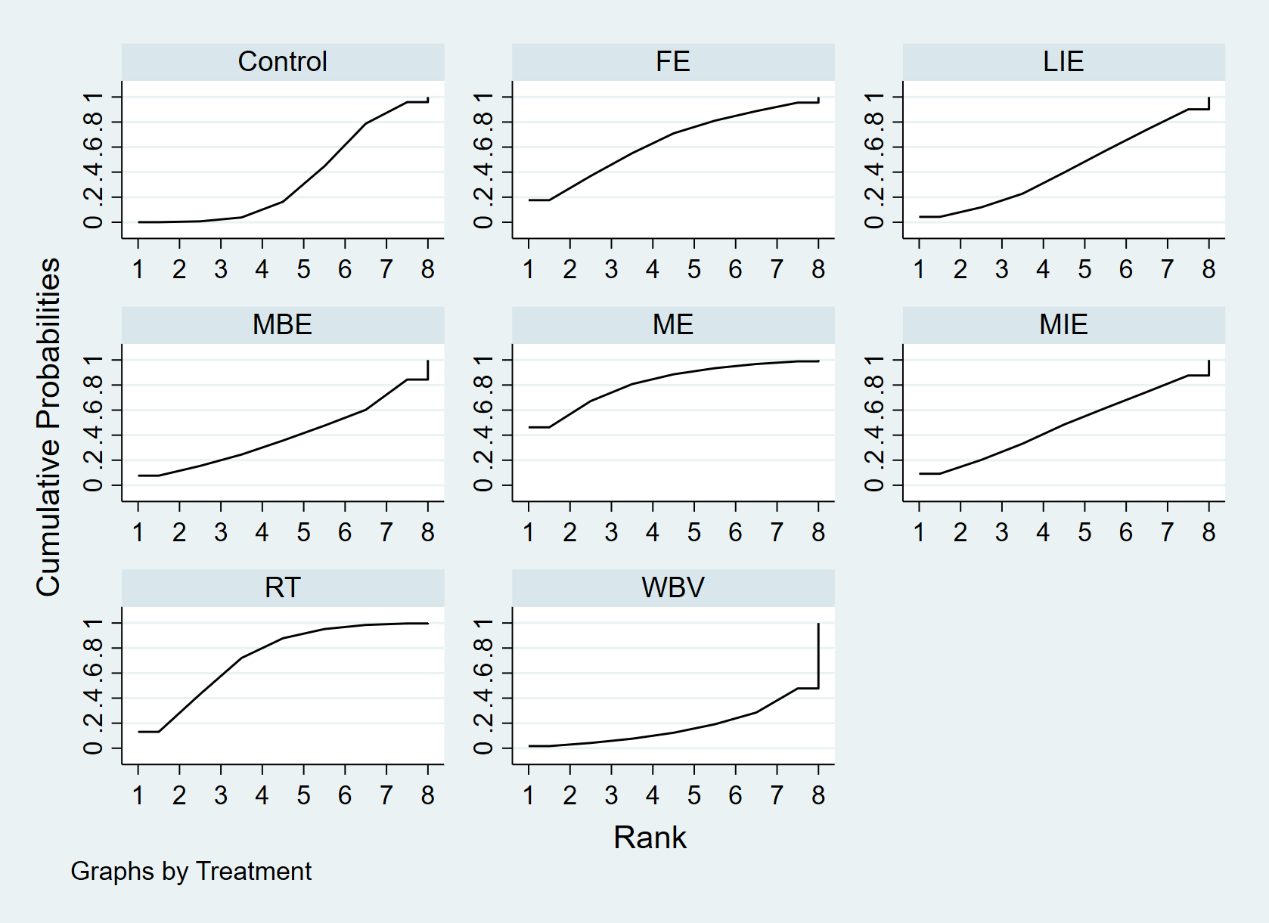

| Control | **-0.01**  **(-0.02, -0.00)** | **0.01**  **(0.00, 0.02)** | **-0.01**  **(-0.03, -0.00)** | -0.00  (-0.01, 0.01) | **-0.03**  **(-0.06, -0.01)** | -0.00  (-0.01, 0.01) | -0.01  (-0.02, 0.00) | -0.01  (-0.02, 0.00) | **-0.01**  **(-0.02, -0.00)** |
| --- | --- | --- | --- | --- | --- | --- | --- | --- | --- |
|  | ME | **0.02**  **(0.01, 0.02)** | -0.01  (-0.02, 0.01) | 0.01  (-0.00, 0.02) | -0.02  (-0.06, 0.00) | 0.01  (-0.01, 0.02) | -0.00  (-0.01, 0.01) | 0.00  (-0.01, 0.01) | -0.00  (-0.01, 0.01) |
|  |  | WE | **-0.02**  **(-0.04, -0.01)** | **-0.01**  **(-0.02, -0.00)** | **-0.04**  **(-0.07, -0.01)** | -0.01  (-0.02, 0.00) | **-0.02**  **(-0.03, -0.00)** | **-0.02**  **(-0.03, -0.00)** | **-0.02**  **(-0.03, -0.01)** |
|  |  |  | FE | 0.01  (-0.00, 0.03) | -0.02  (-0.05, 0.01) | 0.02  (-0.01, 0.03) | 0.01  (-0.01, 0.02) | 0.01  (-0.01, 0.02) | 0.00  (-0.01, 0.02) |
|  |  |  |  | WBV | **-0.03**  **(-0.07, -0.00)** | 0.00  (-0.01, 0.01) | -0.01  (-0.02, 0.00) | -0.01  (-0.02, 0.01) | -0.01  (-0.02, 0.00) |
|  |  |  |  |  | HIE | **0.03**  **(0.00, 0.06)** | 0.02  (-0.00, 0.06) | 0.03  (-0.00, 0.06) | 0.02  (-0.01, 0.05) |
|  |  |  |  |  |  | MIE | -0.01  (-0.02, 0.01) | -0.01  (-0.02, 0.01) | -0.01  (-0.03, 0.00) |
|  |  |  |  |  |  |  | LIE | 0.00  (-0.01, 0.01) | -0.00  (-0.02, 0.01) |
|  |  |  |  |  |  |  |  | MBE | -0.01  (-0.02, 0.01) |
|  |  |  |  |  |  |  |  |  | RT |

### Appendix 18.1: Relative effect estimates for the contrasts between the different interventions and control arms on BMD at LS when exercise intervention with duration between 9 to 18 months

### Appendix 18.2: Relative effect estimates for the contrasts between the different interventions and control arms on BMD at FN when exercise intervention with duration between 9 to 18 months

| Control | -0.01  (-0.02, 0.01) | 0.00  (-0.01, 0.02) | -0.01  (-0.03, 0.02) | -0.00  (-0.02, 0.01) | -0.00  (-0.04, 0.04) | -0.00  (-0.02, 0.02) | -0.00  (-0.02, 0.01) | **-0.02**  **(-0.03, -0.00)** | -0.01  (-0.02, 0.00) |
| --- | --- | --- | --- | --- | --- | --- | --- | --- | --- |
|  | ME | 0.01  (-0.01, 0.03) | 0.00  (-0.02, 0.03) | 0.01  (-0.01, 0.03) | 0.01  (-0.04, 0.05) | 0.01  (-0.01, 0.03) | 0.01  (-0.02, 0.03) | -0.01  (-0.03, 0.01) | -0.00  (-0.02, 0.02) |
|  |  | WE | -0.01  (-0.04, 0.02) | -0.01  (-0.02, 0.01) | -0.01  (-0.05, 0.04) | -0.00  (-0.02, 0.02) | -0.01  (-0.03, 0.02) | -0.02  (-0.05, 0.00) | -0.01  (-0.04, 0.01) |
|  |  |  | FE | 0.00  (-0.03, 0.03) | 0.00  (-0.04, 0.05) | 0.01  (-0.02, 0.04) | 0.00  (-0.03, 0.03) | -0.01  (-0.04, 0.02) | -0.00  (-0.03, 0.02) |
|  |  |  |  | WBV | -0.00  (-0.04, 0.04) | 0.00  (-0.02, 0.03) | -0.00  (-0.03, 0.02) | -0.02  (-0.04, 0.01) | -0.01  (-0.03, 0.01) |
|  |  |  |  |  | HIE | 0.00  (-0.04, 0.05) | 0.00  (-0.04, 0.04) | -0.02  (-0.06, 0.03) | -0.01  (-0.05, 0.03) |
|  |  |  |  |  |  | MIE | -0.00  (-0.03, 0.02) | -0.02  (-0.04, 0.00) | -0.01  (-0.03, 0.01) |
|  |  |  |  |  |  |  | LIE | -0.02  (-0.04, 0.01) | -0.01  (-0.03, 0.01) |
|  |  |  |  |  |  |  |  | MBE | 0.01  (-0.01, 0.03) |
|  |  |  |  |  |  |  |  |  | RT |

### Appendix 18.3: Relative effect estimates for the contrasts between the different interventions and control arms on BMD at TH when exercise intervention with duration between 9 to 18 months

| Control | -0.01  (-0.02, 0.01) | -0.01  (-0.02, 0.00) | -0.00  (-0.01, 0.01) | -0.00  (-0.01, 0.01) | -0.01  (-0.02, 0.00) | **-0.01**  **(-0.02, -0.00)** | -0.01  (-0.02, 0.00) |
| --- | --- | --- | --- | --- | --- | --- | --- |
|  | WE | -0.00  (-0.02, 0.01) | 0.00  (-0.01, 0.02) | 0.00  (-0.01, 0.02) | -0.00  (-0.02, 0.01) | -0.00  (-0.02, 0.01) | -0.00  (-0.01, 0.00) |
|  |  | FE | 0.01  (-0.01, 0.02) | 0.01  (-0.01, 0.02) | 0.00  (-0.01, 0.02) | 0.00  (-0.01, 0.02) | 0.00  (-0.01, 0.01) |
|  |  |  | WBV | -0.00  (-0.01, 0.01) | -0.01  (-0.02, 0.00) | -0.01  (-0.02, 0.00) | -0.01  (-0.02, 0.00) |
|  |  |  |  | LIE | -0.01  (-0.02, 0.01) | -0.01  (-0.02, 0.01) | -0.01  (-0.02, 0.01) |
|  |  |  |  |  | MBE | -0.00  (-0.01, 0.01) | -0.00  (-0.01, 0.01) |
|  |  |  |  |  |  | RT | 0.00  (-0.01, 0.01) |
|  |  |  |  |  |  |  | ME |

### Appendix 19: Cumulative ranking plots to show comparative effectiveness of treatments from a BMD outcome at (A) LS, (B) FN, and (C) FN network meta-analysis when exercise intervention with duration between 9 to 18 months

A.

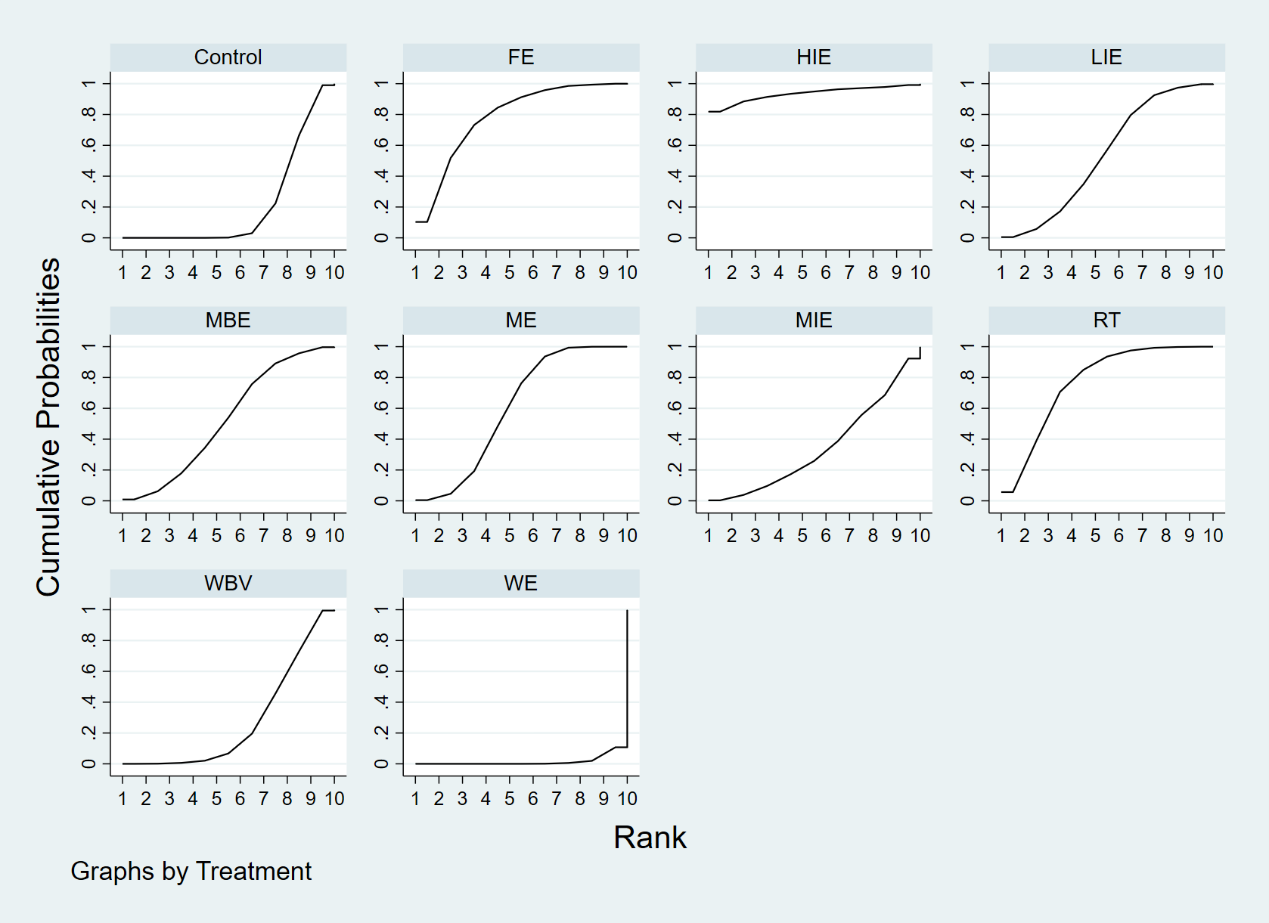

B.

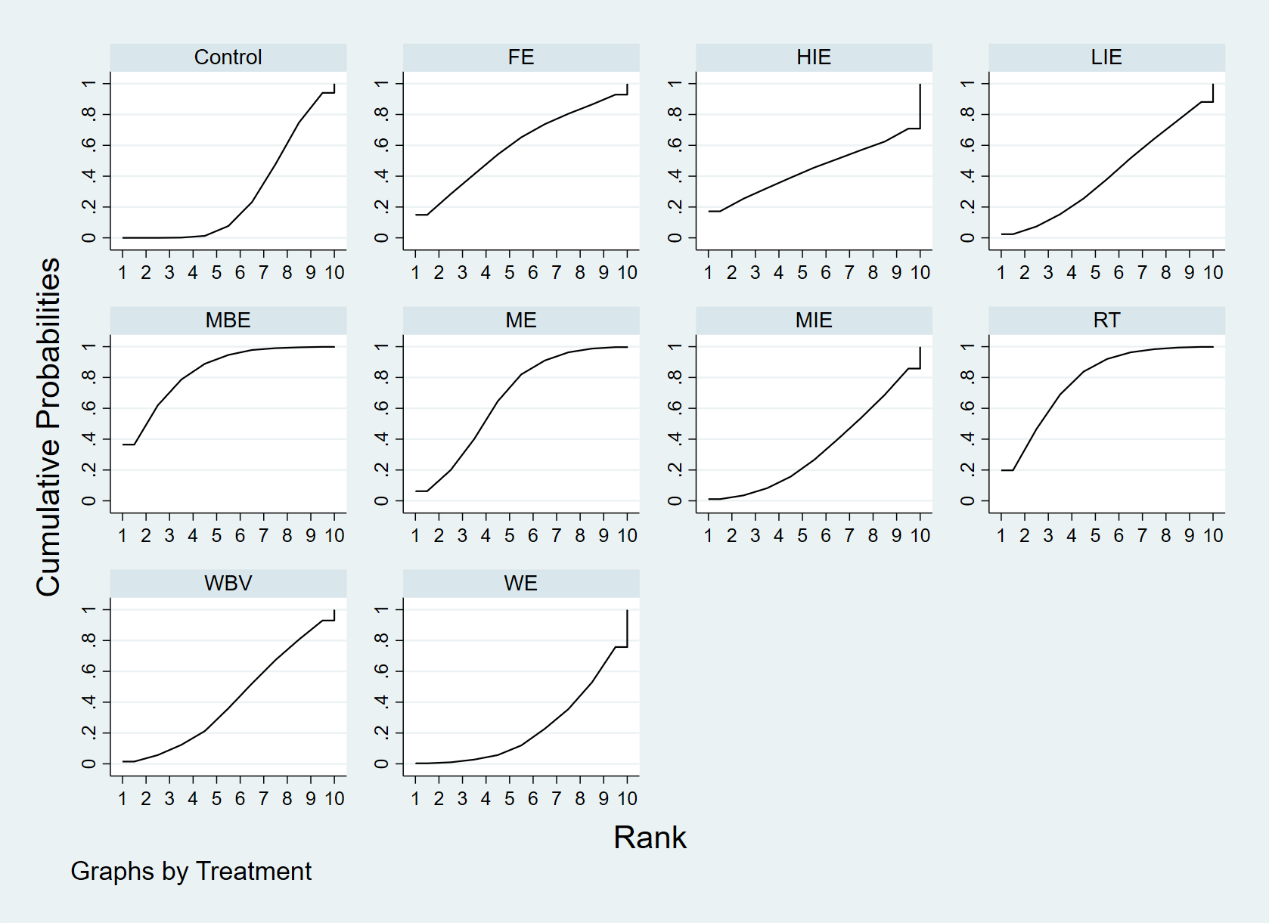

C.

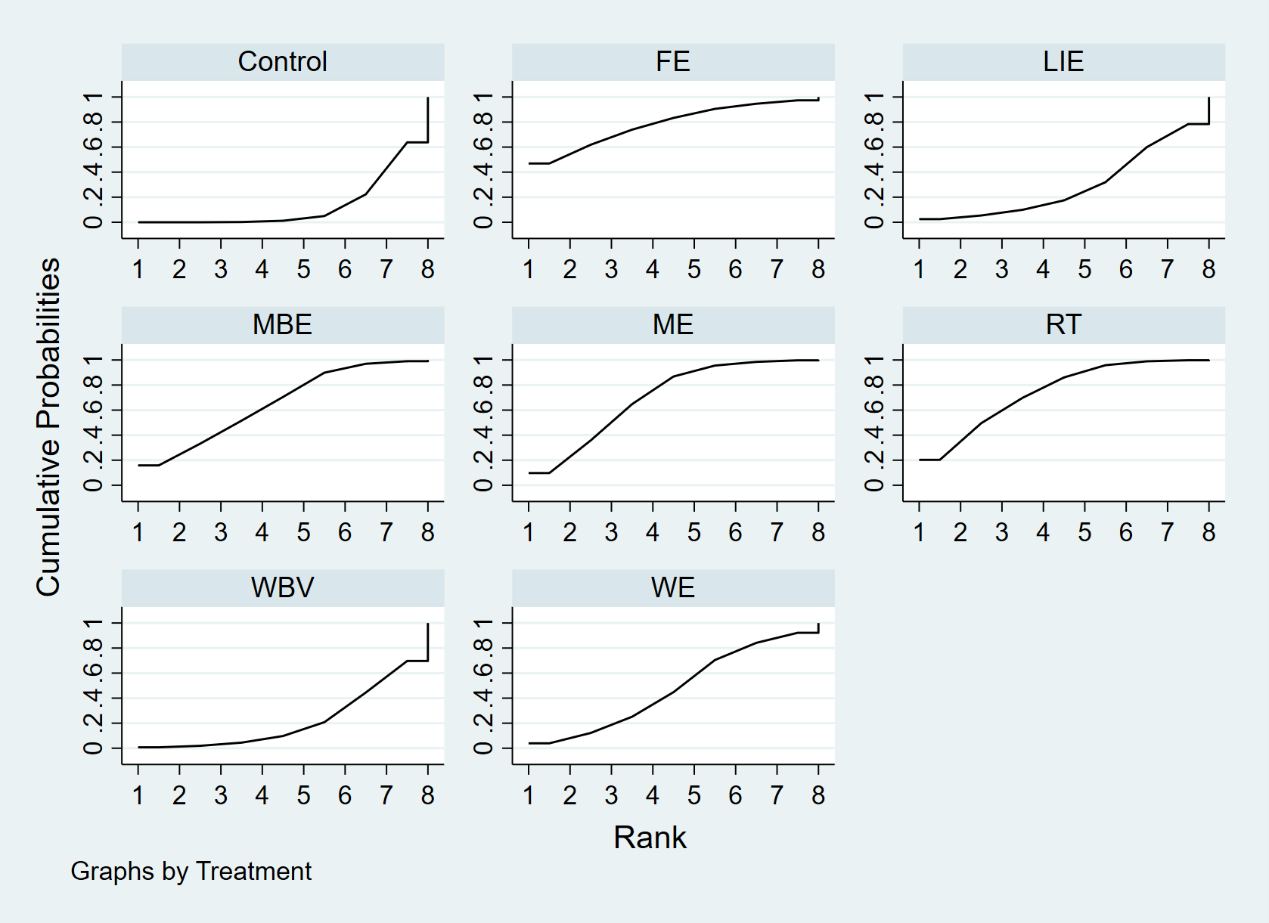

### Appendix 20: Summary of certainty of evidence for network meta-analysis in studies examining the effects of physical exercise in postmenopausal women for BMD at LS, FN, and TH.

| Outcome | Certainty of evidence | Reason for downgrade |
| --- | --- | --- |
| BMD at LS | Very low | Imprecision, incoherence |
| BMD at FN | Low | Imprecision |
| BMD at TH | Very low | Within-study bias, reporting bias, imprecision |

### Appendix 21.1: Details on the CINeMA assessment of each pairwise comparisons for BMD at LS

| Comparison | Number of studies | Within-study bias | Reporting bias | Indirectness | Imprecision | Heterogeneity | Incoherence | Confidence rating |
| --- | --- | --- | --- | --- | --- | --- | --- | --- |
| FE:ME | 1 | No concerns | Undetected | No concerns | Major concerns | No concerns | No concerns | Low |
| FE:PRT | 1 | Some concerns | Undetected | No concerns | Major concerns | No concerns | No concerns | Low |
| HIE:LIE | 1 | Some concerns | Undetected | No concerns | Major concerns | No concerns | No concerns | Low |
| control:HIE | 3 | Some concerns | Undetected | No concerns | No concerns | Major concerns | No concerns | Low |
| LIE:PRT | 1 | Major concerns | Undetected | No concerns | Major concerns | No concerns | No concerns | Very low |
| LIE:WBV | 1 | Some concerns | Undetected | No concerns | Major concerns | No concerns | No concerns | Low |
| control:LIE | 6 | Some concerns | Undetected | No concerns | No concerns | Major concerns | No concerns | Low |
| MBE:RT | 1 | Some concerns | Undetected | No concerns | Major concerns | No concerns | No concerns | Low |
| MBE:WBV | 1 | No concerns | Undetected | No concerns | Major concerns | No concerns | No concerns | Low |
| control:MBE | 7 | Some concerns | Undetected | No concerns | No concerns | Major concerns | No concerns | Low |
| ME:MIE | 2 | Some concerns | Undetected | No concerns | Major concerns | No concerns | No concerns | Low |
| ME:PRT | 1 | Some concerns | Undetected | No concerns | Major concerns | No concerns | No concerns | Low |
| ME:WBV | 1 | Some concerns | Undetected | No concerns | Major concerns | No concerns | No concerns | Low |
| ME:WE | 5 | Some concerns | Undetected | No concerns | No concerns | No concerns | No concerns | Moderate |
| control:ME | 14 | Some concerns | Undetected | No concerns | No concerns | No concerns | No concerns | Moderate |
| MIE:RT | 1 | Some concerns | Undetected | No concerns | Major concerns | No concerns | No concerns | Low |
| MIE:WE | 1 | Some concerns | Undetected | No concerns | Major concerns | No concerns | No concerns | Low |
| control:MIE | 5 | Some concerns | Undetected | No concerns | Major concerns | No concerns | No concerns | Low |
| PRT:WBV | 1 | Some concerns | Undetected | No concerns | Major concerns | No concerns | No concerns | Low |
| control:PRT | 13 | Major concerns | Undetected | No concerns | No concerns | Major concerns | No concerns | Very low |
| control:RT | 4 | Some concerns | Undetected | No concerns | No concerns | No concerns | No concerns | Moderate |
| control:WBE | 1 | Some concerns | Undetected | No concerns | Major concerns | No concerns | Major concerns | Very low |
| WBV:WE | 2 | No concerns | Undetected | No concerns | No concerns | Major concerns | No concerns | Low |
| control:WBV | 9 | No concerns | Undetected | No concerns | No concerns | Major concerns | No concerns | Low |
| FE:HIE | 0 | Some concerns | Undetected | No concerns | Major concerns | No concerns | Major concerns | Very low |
| FE:LIE | 0 | Some concerns | Undetected | No concerns | Major concerns | No concerns | Major concerns | Very low |
| FE:MBE | 0 | Some concerns | Undetected | No concerns | Major concerns | No concerns | Major concerns | Very low |
| FE:MIE | 0 | Some concerns | Undetected | No concerns | Major concerns | No concerns | Major concerns | Very low |
| FE:RT | 0 | Some concerns | Undetected | No concerns | Major concerns | No concerns | Major concerns | Very low |
| FE:WBE | 0 | Some concerns | Undetected | No concerns | Major concerns | No concerns | Major concerns | Very low |
| FE:WBV | 0 | No concerns | Undetected | No concerns | Major concerns | No concerns | Major concerns | Very low |
| FE:WE | 0 | No concerns | Undetected | No concerns | Major concerns | No concerns | Major concerns | Very low |
| control:FE | 0 | Some concerns | Undetected | No concerns | Major concerns | No concerns | Major concerns | Very low |
| HIE:MBE | 0 | Some concerns | Undetected | No concerns | Major concerns | No concerns | Major concerns | Very low |
| HIE:ME | 0 | Some concerns | Undetected | No concerns | Major concerns | No concerns | Major concerns | Very low |
| HIE:MIE | 0 | Some concerns | Undetected | No concerns | Major concerns | No concerns | Major concerns | Very low |
| HIE:PRT | 0 | Some concerns | Undetected | No concerns | Major concerns | No concerns | Major concerns | Very low |
| HIE:RT | 0 | Some concerns | Undetected | No concerns | Major concerns | No concerns | Major concerns | Very low |
| HIE:WBE | 0 | Some concerns | Undetected | No concerns | Major concerns | No concerns | Major concerns | Very low |
| HIE:WBV | 0 | Some concerns | Undetected | No concerns | Major concerns | No concerns | Major concerns | Very low |
| HIE:WE | 0 | Some concerns | Undetected | No concerns | No concerns | No concerns | Major concerns | Low |
| LIE:MBE | 0 | Some concerns | Undetected | No concerns | Major concerns | No concerns | Major concerns | Very low |
| LIE:ME | 0 | Some concerns | Undetected | No concerns | Major concerns | No concerns | Major concerns | Very low |
| LIE:MIE | 0 | Some concerns | Undetected | No concerns | Major concerns | No concerns | Major concerns | Very low |
| LIE:RT | 0 | Some concerns | Undetected | No concerns | Major concerns | No concerns | Major concerns | Very low |
| LIE:WBE | 0 | Some concerns | Undetected | No concerns | Major concerns | No concerns | Major concerns | Very low |
| LIE:WE | 0 | Some concerns | Undetected | No concerns | No concerns | Major concerns | Major concerns | Very low |
| MBE:ME | 0 | Some concerns | Undetected | No concerns | Major concerns | No concerns | Major concerns | Very low |
| MBE:MIE | 0 | Some concerns | Undetected | No concerns | Major concerns | No concerns | Major concerns | Very low |
| MBE:PRT | 0 | Some concerns | Undetected | No concerns | Major concerns | No concerns | Major concerns | Very low |
| MBE:WBE | 0 | Some concerns | Undetected | No concerns | Major concerns | No concerns | Major concerns | Very low |
| MBE:WE | 0 | Some concerns | Undetected | No concerns | No concerns | Major concerns | Major concerns | Very low |
| ME:RT | 0 | Some concerns | Undetected | No concerns | Major concerns | No concerns | Major concerns | Very low |
| ME:WBE | 0 | Some concerns | Undetected | No concerns | Major concerns | No concerns | Major concerns | Very low |
| MIE:PRT | 0 | Some concerns | Undetected | No concerns | Major concerns | No concerns | Major concerns | Very low |
| MIE:WBE | 0 | Some concerns | Undetected | No concerns | Major concerns | No concerns | Major concerns | Very low |
| MIE:WBV | 0 | Some concerns | Undetected | No concerns | Major concerns | No concerns | Major concerns | Very low |
| PRT:RT | 0 | Some concerns | Undetected | No concerns | Major concerns | No concerns | Major concerns | Very low |
| PRT:WBE | 0 | Some concerns | Undetected | No concerns | Major concerns | No concerns | Major concerns | Very low |
| PRT:WE | 0 | Some concerns | Undetected | No concerns | No concerns | Major concerns | Major concerns | Very low |
| RT:WBE | 0 | Some concerns | Undetected | No concerns | Major concerns | No concerns | Major concerns | Very low |
| RT:WBV | 0 | Some concerns | Undetected | No concerns | Major concerns | No concerns | Major concerns | Very low |
| RT:WE | 0 | Some concerns | Undetected | No concerns | No concerns | No concerns | Major concerns | Very low |
| WBE:WBV | 0 | Some concerns | Undetected | No concerns | Major concerns | No concerns | Major concerns | Very low |
| WBE:WE | 0 | Some concerns | Undetected | No concerns | Major concerns | No concerns | Major concerns | Very low |
| control:WE | 0 | Some concerns | Undetected | No concerns | Major concerns | No concerns | Major concerns | Very low |

### Appendix 21.2: Details on the CINeMA assessment of each pairwise comparisons for BMD at FN

| Comparison | Number of studies | Within-study bias | Reporting bias | Indirectness | Imprecision | Heterogeneity | Incoherence | Confidence rating |
| --- | --- | --- | --- | --- | --- | --- | --- | --- |
| 1:03 | 1 | Some concerns | Undetected | No concerns | Major concerns | No concerns | No concerns | Low |
| 1:04 | 6 | No concerns | Undetected | No concerns | No concerns | Major concerns | No concerns | Low |
| 1:05 | 2 | Some concerns | Undetected | No concerns | Major concerns | No concerns | No concerns | Low |
| 1:06 | 5 | Some concerns | Undetected | No concerns | Major concerns | No concerns | No concerns | Low |
| 1:07 | 8 | Some concerns | Undetected | No concerns | Major concerns | No concerns | No concerns | Low |
| 1:08 | 6 | Some concerns | Undetected | No concerns | No concerns | Major concerns | No concerns | Low |
| 1:09 | 1 | Some concerns | Undetected | No concerns | Major concerns | No concerns | No concerns | Low |
| 1:10 | 2 | Some concerns | Undetected | No concerns | Major concerns | No concerns | No concerns | Low |
| 1:11 | 15 | Some concerns | Undetected | No concerns | Major concerns | No concerns | No concerns | Low |
| 1:12 | 9 | Some concerns | Undetected | No concerns | No concerns | Major concerns | Major concerns | Very low |
| 2:04 | 2 | No concerns | Undetected | No concerns | Major concerns | No concerns | No concerns | Low |
| 2:06 | 1 | Some concerns | Undetected | No concerns | Major concerns | No concerns | No concerns | Low |
| 4:07 | 1 | Some concerns | Undetected | No concerns | Major concerns | No concerns | Major concerns | Very low |
| 4:08 | 1 | No concerns | Undetected | No concerns | Major concerns | No concerns | No concerns | Low |
| 10:06 | 1 | Some concerns | Undetected | No concerns | Major concerns | No concerns | No concerns | Low |
| 11:03 | 2 | Some concerns | Undetected | No concerns | Major concerns | No concerns | No concerns | Low |
| 11:06 | 1 | Some concerns | Undetected | No concerns | Major concerns | No concerns | No concerns | Low |
| 11:07 | 1 | Some concerns | Undetected | No concerns | Major concerns | No concerns | No concerns | Low |
| 12:02 | 2 | Some concerns | Undetected | No concerns | No concerns | Major concerns | No concerns | Low |
| 12:03 | 1 | Some concerns | Undetected | No concerns | Major concerns | No concerns | Major concerns | Very low |
| 1:02 | 0 | Some concerns | Undetected | No concerns | Major concerns | No concerns | No concerns | Low |
| 2:03 | 0 | Some concerns | Undetected | No concerns | Major concerns | No concerns | No concerns | Low |
| 2:05 | 0 | Some concerns | Undetected | No concerns | Major concerns | No concerns | No concerns | Low |
| 2:07 | 0 | Some concerns | Undetected | No concerns | Major concerns | No concerns | No concerns | Low |
| 2:08 | 0 | Some concerns | Undetected | No concerns | Major concerns | No concerns | No concerns | Low |
| 2:09 | 0 | Some concerns | Undetected | No concerns | Major concerns | No concerns | No concerns | Low |
| 10:02 | 0 | Some concerns | Undetected | No concerns | Major concerns | No concerns | No concerns | Low |
| 11:02 | 0 | Some concerns | Undetected | No concerns | Major concerns | No concerns | No concerns | Low |
| 3:04 | 0 | Some concerns | Undetected | No concerns | Major concerns | No concerns | No concerns | Low |
| 3:05 | 0 | Some concerns | Undetected | No concerns | Major concerns | No concerns | No concerns | Low |
| 3:06 | 0 | Some concerns | Undetected | No concerns | Major concerns | No concerns | No concerns | Low |
| 3:07 | 0 | Some concerns | Undetected | No concerns | Major concerns | No concerns | No concerns | Low |
| 3:08 | 0 | Some concerns | Undetected | No concerns | Major concerns | No concerns | No concerns | Low |
| 3:09 | 0 | Some concerns | Undetected | No concerns | Major concerns | No concerns | No concerns | Low |
| 10:03 | 0 | Some concerns | Undetected | No concerns | Major concerns | No concerns | No concerns | Low |
| 4:05 | 0 | Some concerns | Undetected | No concerns | Major concerns | No concerns | No concerns | Low |
| 4:06 | 0 | Some concerns | Undetected | No concerns | Major concerns | No concerns | No concerns | Low |
| 4:09 | 0 | Some concerns | Undetected | No concerns | Major concerns | No concerns | No concerns | Low |
| 10:04 | 0 | Some concerns | Undetected | No concerns | Major concerns | No concerns | No concerns | Low |
| 11:04 | 0 | Some concerns | Undetected | No concerns | Major concerns | No concerns | No concerns | Low |
| 12:04 | 0 | Some concerns | Undetected | No concerns | Major concerns | No concerns | No concerns | Low |
| 5:06 | 0 | Some concerns | Undetected | No concerns | Major concerns | No concerns | No concerns | Low |
| 5:07 | 0 | Some concerns | Undetected | No concerns | Major concerns | No concerns | No concerns | Low |
| 5:08 | 0 | Some concerns | Undetected | No concerns | Major concerns | No concerns | No concerns | Low |
| 5:09 | 0 | Some concerns | Undetected | No concerns | Major concerns | No concerns | No concerns | Low |
| 10:05 | 0 | Some concerns | Undetected | No concerns | Major concerns | No concerns | No concerns | Low |
| 11:05 | 0 | Some concerns | Undetected | No concerns | Major concerns | No concerns | No concerns | Low |
| 12:05 | 0 | Some concerns | Undetected | No concerns | Major concerns | No concerns | No concerns | Low |
| 6:07 | 0 | Some concerns | Undetected | No concerns | Major concerns | No concerns | No concerns | Low |
| 6:08 | 0 | Some concerns | Undetected | No concerns | Major concerns | No concerns | No concerns | Low |
| 6:09 | 0 | Some concerns | Undetected | No concerns | Major concerns | No concerns | No concerns | Low |
| 12:06 | 0 | Some concerns | Undetected | No concerns | No concerns | Major concerns | No concerns | Low |
| 7:08 | 0 | Some concerns | Undetected | No concerns | Major concerns | No concerns | No concerns | Low |
| 7:09 | 0 | Some concerns | Undetected | No concerns | Major concerns | No concerns | No concerns | Low |
| 10:07 | 0 | Some concerns | Undetected | No concerns | Major concerns | No concerns | No concerns | Low |
| 12:07 | 0 | Some concerns | Undetected | No concerns | No concerns | Major concerns | No concerns | Low |
| 8:09 | 0 | Some concerns | Undetected | No concerns | Major concerns | No concerns | No concerns | Low |
| 10:08 | 0 | Some concerns | Undetected | No concerns | Major concerns | No concerns | No concerns | Low |
| 11:08 | 0 | Some concerns | Undetected | No concerns | Major concerns | No concerns | No concerns | Low |
| 12:08 | 0 | Some concerns | Undetected | No concerns | Major concerns | No concerns | No concerns | Low |
| 10:09 | 0 | Some concerns | Undetected | No concerns | Major concerns | No concerns | No concerns | Low |
| 11:09 | 0 | Some concerns | Undetected | No concerns | Major concerns | No concerns | No concerns | Low |
| 12:09 | 0 | Some concerns | Undetected | No concerns | Major concerns | No concerns | No concerns | Low |
| 10:11 | 0 | Some concerns | Undetected | No concerns | Major concerns | No concerns | No concerns | Low |
| 10:12 | 0 | Some concerns | Undetected | No concerns | Major concerns | No concerns | No concerns | Low |
| 11:12 | 0 | Some concerns | Undetected | No concerns | No concerns | Major concerns | No concerns | Low |

### Appendix 21.3: Details on the CINeMA assessment of each pairwise comparisons for BMD at TH

| Comparison | Number of studies | Within-study bias | Reporting bias | Indirectness | Imprecision | Heterogeneity | Incoherence | Confidence rating |
| --- | --- | --- | --- | --- | --- | --- | --- | --- |
| 1:03 | 1 | Some concerns | Suspected | No concerns | No concerns | Major concerns | No concerns | Very low |
| 1:04 | 3 | No concerns | Suspected | No concerns | Major concerns | No concerns | No concerns | Very low |
| 1:06 | 1 | No concerns | Suspected | No concerns | Major concerns | No concerns | No concerns | Very low |
| 1:07 | 3 | Some concerns | Suspected | No concerns | Major concerns | No concerns | No concerns | Very low |
| 1:08 | 3 | No concerns | Suspected | No concerns | Major concerns | No concerns | No concerns | Very low |
| 1:09 | 1 | Some concerns | Suspected | No concerns | Major concerns | No concerns | No concerns | Very low |
| 1:10 | 2 | Some concerns | Suspected | No concerns | No concerns | No concerns | No concerns | Very low |
| 1:11 | 8 | Some concerns | Suspected | No concerns | No concerns | Major concerns | No concerns | Very low |
| 1:12 | 5 | Some concerns | Suspected | No concerns | No concerns | Major concerns | No concerns | Very low |
| 4:08 | 1 | No concerns | Suspected | No concerns | Major concerns | No concerns | No concerns | Very low |
| 11:03 | 2 | Some concerns | Suspected | No concerns | Major concerns | No concerns | No concerns | Very low |
| 11:06 | 1 | No concerns | Suspected | No concerns | Major concerns | No concerns | No concerns | Very low |
| 12:02 | 3 | No concerns | Suspected | No concerns | Major concerns | No concerns | No concerns | Very low |
| 12:03 | 1 | Some concerns | Suspected | No concerns | Major concerns | No concerns | No concerns | Very low |
| 1:02 | 0 | Some concerns | Suspected | No concerns | Major concerns | No concerns | No concerns | Very low |
| 2:03 | 0 | Some concerns | Suspected | No concerns | Major concerns | No concerns | No concerns | Very low |
| 2:04 | 0 | No concerns | Suspected | No concerns | Major concerns | No concerns | No concerns | Very low |
| 2:06 | 0 | No concerns | Suspected | No concerns | Major concerns | No concerns | No concerns | Very low |
| 2:07 | 0 | Some concerns | Suspected | No concerns | Major concerns | No concerns | No concerns | Very low |
| 2:08 | 0 | No concerns | Suspected | No concerns | Major concerns | No concerns | No concerns | Very low |
| 2:09 | 0 | Some concerns | Suspected | No concerns | Major concerns | No concerns | No concerns | Very low |
| 10:02 | 0 | Some concerns | Suspected | No concerns | Major concerns | No concerns | No concerns | Very low |
| 11:02 | 0 | Some concerns | Suspected | No concerns | Major concerns | No concerns | No concerns | Very low |
| 3:04 | 0 | No concerns | Suspected | No concerns | No concerns | Major concerns | No concerns | Very low |
| 3:06 | 0 | No concerns | Suspected | No concerns | Major concerns | No concerns | No concerns | Very low |
| 3:07 | 0 | Some concerns | Suspected | No concerns | Major concerns | No concerns | No concerns | Very low |
| 3:08 | 0 | Some concerns | Suspected | No concerns | Major concerns | No concerns | No concerns | Very low |
| 3:09 | 0 | Some concerns | Suspected | No concerns | Major concerns | No concerns | No concerns | Very low |
| 10:03 | 0 | Some concerns | Suspected | No concerns | Major concerns | No concerns | No concerns | Very low |
| 4:06 | 0 | No concerns | Suspected | No concerns | Major concerns | No concerns | No concerns | Very low |
| 4:07 | 0 | No concerns | Suspected | No concerns | Major concerns | No concerns | No concerns | Very low |
| 4:09 | 0 | No concerns | Suspected | No concerns | Major concerns | No concerns | No concerns | Very low |
| 10:04 | 0 | No concerns | Suspected | No concerns | No concerns | No concerns | No concerns | Low |
| 11:04 | 0 | Some concerns | Suspected | No concerns | No concerns | Major concerns | No concerns | Very low |
| 12:04 | 0 | No concerns | Suspected | No concerns | No concerns | Major concerns | No concerns | Very low |
| 6:07 | 0 | Some concerns | Suspected | No concerns | Major concerns | No concerns | No concerns | Very low |
| 6:08 | 0 | No concerns | Suspected | No concerns | Major concerns | No concerns | No concerns | Very low |
| 6:09 | 0 | Some concerns | Suspected | No concerns | Major concerns | No concerns | No concerns | Very low |
| 10:06 | 0 | Some concerns | Suspected | No concerns | Major concerns | No concerns | No concerns | Very low |
| 12:06 | 0 | No concerns | Suspected | No concerns | Major concerns | No concerns | No concerns | Very low |
| 7:08 | 0 | Some concerns | Suspected | No concerns | Major concerns | No concerns | No concerns | Very low |
| 7:09 | 0 | Some concerns | Suspected | No concerns | Major concerns | No concerns | No concerns | Very low |
| 10:07 | 0 | Some concerns | Suspected | No concerns | Major concerns | No concerns | No concerns | Very low |
| 11:07 | 0 | Some concerns | Suspected | No concerns | Major concerns | No concerns | No concerns | Very low |
| 12:07 | 0 | Some concerns | Suspected | No concerns | Major concerns | No concerns | No concerns | Very low |
| 8:09 | 0 | Some concerns | Suspected | No concerns | Major concerns | No concerns | No concerns | Very low |
| 10:08 | 0 | Some concerns | Suspected | No concerns | Major concerns | No concerns | No concerns | Very low |
| 11:08 | 0 | Some concerns | Suspected | No concerns | Major concerns | No concerns | No concerns | Very low |
| 12:08 | 0 | Some concerns | Suspected | No concerns | Major concerns | No concerns | No concerns | Very low |
| 10:09 | 0 | Some concerns | Suspected | No concerns | Major concerns | No concerns | No concerns | Very low |
| 11:09 | 0 | Some concerns | Suspected | No concerns | Major concerns | No concerns | No concerns | Very low |
| 12:09 | 0 | Some concerns | Suspected | No concerns | Major concerns | No concerns | No concerns | Very low |
| 10:11 | 0 | Some concerns | Suspected | No concerns | Major concerns | No concerns | No concerns | Very low |
| 10:12 | 0 | Some concerns | Suspected | No concerns | Major concerns | No concerns | No concerns | Very low |
| 11:12 | 0 | Some concerns | Suspected | No concerns | Major concerns | No concerns | No concerns | Very low |

### Appendix 22: Funnel plot for the BMT at (A) LS, (B) FN, and (C) FN network

A.

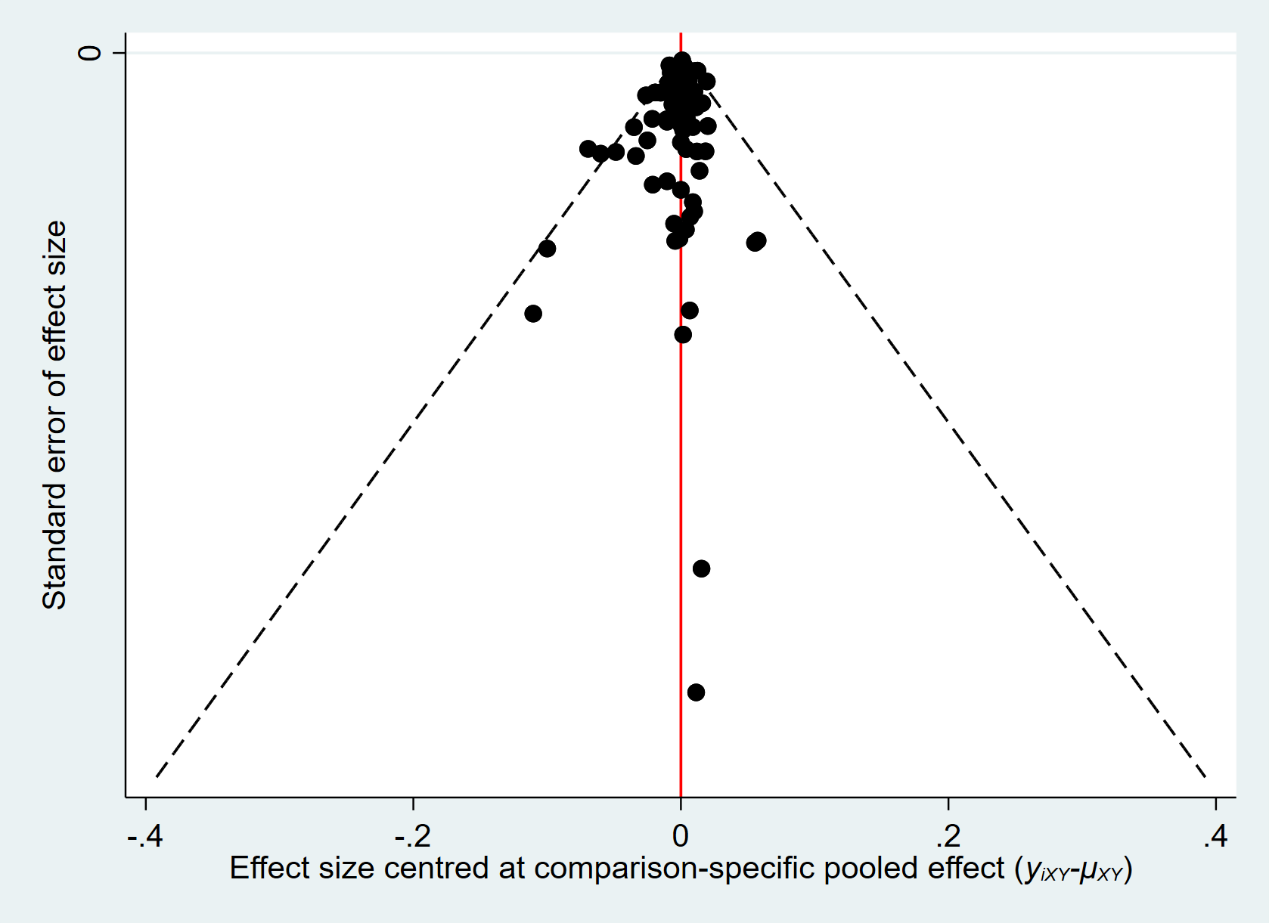
B.

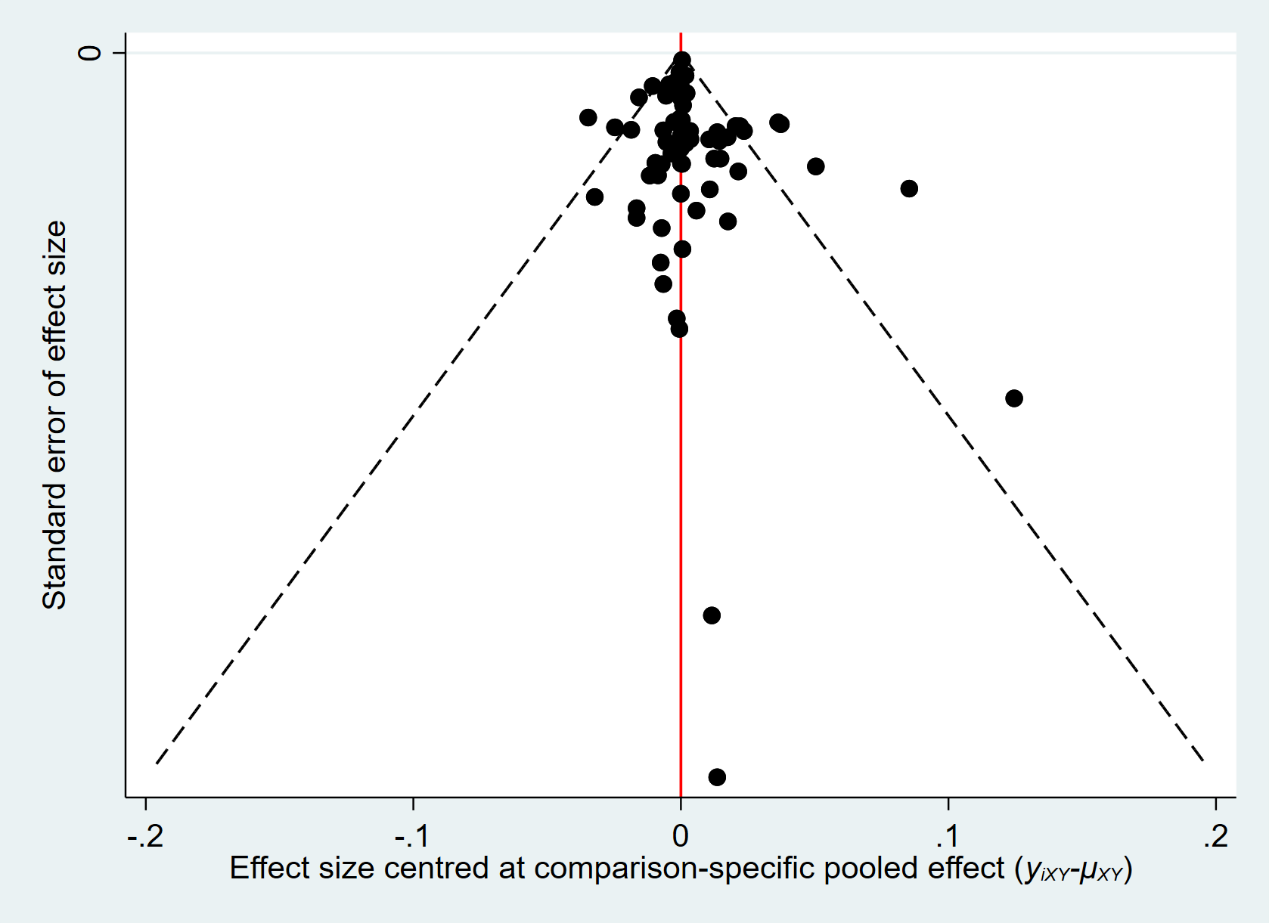

C.

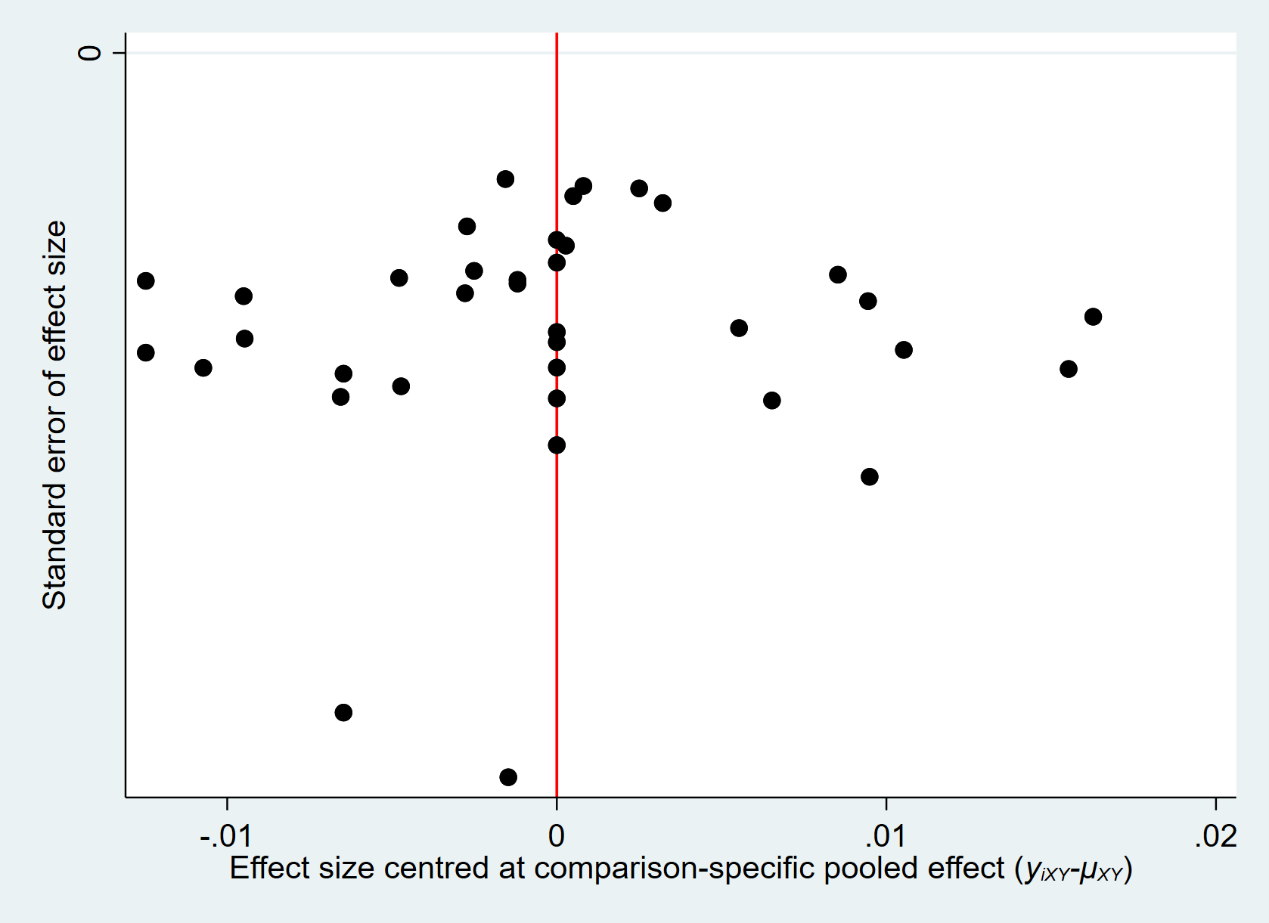
